# Causal and Candidate Gene Variants in a Large Cohort of Women with Primary Ovarian Insufficiency

**DOI:** 10.1101/2021.06.02.21258048

**Authors:** B Gorsi, EJ Hernandez, B Moore, M Moriwaki, CY Chow, E Coelho, E Taylor, C Lu, A Walker, P Touraine, LM Nelson, AR Cooper, ER Mardis, A Rajkovic, M Yandell, CK Welt

## Abstract

A genetic etiology accounts for unexplained primary ovarian insufficiency (POI; amenorrhea with an elevated FSH level). Subjects with POI (n=291) and controls recruited for health in old age or 1000 Genomes (n=233) underwent whole exome or whole genome sequencing. Data were analyzed using a rare variant scoring method and a Bayes factor-based framework for identifying genes harboring pathogenic variants. Candidate heterozygous variants were identified in known genes and genes with functional evidence. Gene sets with increased burden of deleterious alleles included the categories transcription and translation, DNA damage and repair, meiosis and cell division. Variants were found in novel genes from the enhanced categories. Functional evidence supported 7 new risk genes for POI (*USP36, VCP, WDR33, PIWIL3*, *NPM2*, *LLGL1* and *BOD1L1*). Aggregating clinical data and genetic risk with a categorical approach may expand the genetic architecture of heterozygous rare gene variants causing risk for POI.

## Introduction

Primary ovarian insufficiency (POI) encompasses a continuum from infertility in women with ovarian dysfunction to early menopause^1^. The cause of POI remains unknown in the majority of women, making intervention impossible to initiate until it is too late^1^.

Data overwhelmingly support a genetic cause in women with POI^2–4^. Twin studies estimate heritability from 53-71%^2–4^. There is a strong relationship between age at menopause in mothers and daughters, with an odds ratio of 6 (95% confidence intervals 3.4, 10.7) for early menopause in daughters whose mothers had early menopause^5^. In small studies it has been estimated that up to 30% of POI cases are familial^6^. The most common known genetic causes include X chromosome defects, *FMR1* premutations and autoimmune causes^1^. Nevertheless, the additive effect of these and known iatrogenic causes explain less than 30% of POI. A remarkable number of new POI-associated genes have been discovered, facilitated by whole exome sequencing (WES) in consanguineous and large families^7–15^. The women in these families typically develop POI before puberty, also termed ovarian dysgenesis. Mutations in the DNA of these women have been identified in genes important for mitochondrial function, meiosis, homologous recombination and DNA damage repair^7,8,10,12,16^.

The inheritance pattern for POI is not recessive in all cases. Heterozygous mutations in genes such as *eIF4ENIF1* cause POI in women in the mid reproductive years^9^. Recessive gene mutations found to cause POI and primary amenorrhea also cause POI or earlier menopause in heterozygous mothers, demonstrating that dominant and semi-dominant mutations may be causal^17,18^, with heterozygous damaging variants in known genes or in two or more candidate genes causing POI^19,20^. One study of POI suggested an additive effect from common variants contributing to age at menopause, with a recent study suggesting that common variation may explain a portion of earlier age at menopause, as low as age 34 years^21,22^.

Most previous WES analyses in large numbers of women with sporadic POI used a candidate gene approach to identify gene mutations most likely to cause POI. However, a variant-centric approach has identified novel POI candidate genes^19,23^. We used an unbiased approach and a new prioritization algorithm (GEM)^24–26^ to identify damaging gene variants in known POI genes. We then used a category-wide association approach to test the hypothesis that additional candidate mutations could be found in clustered gene sets created using known genes and gene candidates from model organisms^27,28^. We demonstrated a significant enhancement in identified gene sets in women with POI compared to controls. These gene sets revealed additional candidate genes in POI, with seven genes confirmed by functional studies to play a role in oocyte or ovary development. These findings improve our understanding of the genetic architecture of POI, an extremely heterogeneous disorder.

## Subjects

All subjects were diagnosed with POI as defined by at least 4 months of amenorrhea and an FSH level in the menopausal range. All women were 18 years or older, and had a 46XX karyotype, and normal *FMR1* repeat number. Subjects (n=35) were recruited in Boston. Additional subjects were recruited from the Partners Biobank (n=63). A second cohort (n=98) was recruited at the National Institute of Health (NIH) for a study of non-syndromic POI^29^. These subjects were re-consented to have their DNA undergo WES at Washington University (LMN, ARC and ERM). A third cohort was recruited from Pittsburgh (n=20), Italy (n=43) and France (n=32)(AR and PT). All Boston subjects underwent a medical history and physical exam and family history. Subjects from the NIH, Pittsburgh, Italy and France had limited phenotypic data.

Control subjects for category-wide association using GEM^24^ included 96 unrelated, unaffected subjects recruited for health in old age and 137 CEU, FIN and GBR samples from the 1000 Genomes Project (total n=233 controls)^30,31^. The majority of CEU samples are from Utah families recruited for large family size (n=47 of 61)^32^. The control subjects underwent whole genome sequencing, as previously described^33^. All subjects provided written, informed consent from the University of Utah, Washington University, University of Pittsburgh or the Sorbonne Universite IRB.

## Methods

DNA samples were extracted (Qiagen) and subjected to WES. The Boston cohort was sequenced using the Illumina HiSeq 2000 (Illumina). All candidate susceptibility variants in the Boston cohort were Sanger sequenced for verification. Sequencing of the NIH/Washington University cohort was performed using the Roche NimbleGen VCRome 2.1 (HGSC design) exome capture and the Illumina HiSeq 2500 for sequencing at the McDonnell Genome Institute at Washington University. The dataset was accessed through dbGAP (NIH approved request #47895-1, Project #11971). The Pittsburgh, French and Italian cohorts were sequenced at the Pittsburgh Clinical Genomics Laboratory using the Haloplex Exome Target Enrichment System or the Agilent SureSelect V5 Capture Kit (Agilent Technologies, Santa Clara, CA), and 2× 100 bp paired-end WES was performed on an Illumina HiSeq 2500 (San Diego, CA, USA).

The control subjects’ DNA underwent whole genome sequencing (WGS) using the Illumina X Ten sequencing platform (Nantomics, Culver City, CA). The comparison of variants in cases using WES versus controls using WGS would result in a conservative estimate of variants in cases based on the higher coverage expected from WGS.

### Alignment and Variant Calling

Alignment and variant calling were performed by the Utah Center for Genetic Discovery (UCGD) core services. Fastq files were downloaded from the Pittsburgh Clinical Genomics Laboratory and dbGAP. Variants were called through the UCGD pipeline using the Sentieon software package (https://www.sentieon.com) ^34^. Reads were aligned to the human reference build GRCh37 using BWA-MEM (Burrows-Wheeler Aligner). SAMBLASTER was used to mark duplicate reads and de-duplicate aligned BAM files. Aligned BAM files underwent INDEL realignment and base recalibration using Realigner and QualCal algorithms from the Sentieon software package3 to produce polished BAM files. Each polished BAM file was processed using the Sentieon’s Haplotyper algorithm to produce gVCF files^35^. Sample gVCF files were combined and jointly genotyped with 728 samples comprised of the 1000 genomes project (CEU) samples and samples unrelated to reproduction or cancer phenotypes to produce a multi-sample VCF file. To produce the final VCF variant quality scores, VCF files were recalibrated using Sentieon’s VarCal algorithm to estimate the accuracy of variant calls and reduce potential false positive calls.

### Quality Control

Quality control algorithms were applied to sequence reads (Fastq files), aligned reads (BAM files) and variants (VCF files)^36^. Fastp was used to evaluate read quality, read duplication rate, presence of adapter and overrepresented sequences in Fastq files^37^. Indexcov was used to estimate depth and coverage of aligned sequence data using BAM indexes. Further alignment quality metrics were calculated on BAM files with samtools stats. The 291 cases were sequenced using different exome capture kits, we therefore standardized QC analysis regions with a bed file made up of exonic regions from coding gene models from RefSeq and Ensemble gene sets. These regions were used to obtain the total number of reads, percentage aligned reads and mean and median coverage for all samples.

Variant quality metrics were calculated by running bcftools stats^38–40^. The overall quality of VCF callsets were evaluated using Peddy to confirm sex, relatedness, heterozygosity and ancestry of each individual and identifying potential sample-level data quality issues^41^.

### Identification of Damaging Gene Variants

For each case, the uploaded VCF file was scored with VAAST Variant Prioritizer (VVP) and Variant Annotation Analysis and Search Tool (VAAST) to prioritize potentially deleterious variants and damaged genes^25,42^. VVP and VAAST use a likelihood ratio test (LRT) to score each variant and the aggregate burden of variants for each gene in affected individuals relative to a set of 2,492 control genomes of healthy individuals from the 1000 Genomes Project^43^. The LRT incorporates three components of each variant; the severity of amino acid substitution, phylogenetic conservation of the variant, and the frequency of the variant relative to the control population. The sum of the top scoring variant(s) based (one variant for dominant inheritance and two variants for recessive inheritance) represents the cumulative likelihood ratio (CLR) for a given gene. The significance of each gene’s VAAST CLRT score is evaluated by a permutation test that randomizes the case/control status of individuals in each of 1e^6^ permutations and generates a permutation *p* value for the gene. The output from VAAST is an ordered gene list ranked for the probability of being damaged relative to the control genomes.

Variants identified in the VAAST analysis above were further refined by selecting only variants found at a minor allele frequency (MAF) <0.001 and with no homozygotes found in gnomAD^44,45^. The choice of a MAF <0.001 cutoff was based on the frequency of the fragile X premutation. The prevalence of the premutation in the population is 0.004 and it is the most common single gene cause of POI identified to date. A fragile X premutation accounts for only 6% of sporadic POI cases^46^, making 0.001 a conservative upper bound for the risk attributable to any one gene. We also removed variants in genes known to tolerate a large burden of genetic variation such as olfactory receptors, snoRNAs, mucins and T cell receptors^47^. Finally, we required an Omicia score of >0.7; a meta-classifier that combines scores from SIFT, PolyPhen, MutationTaster and PhyloP to predict pathogenicity^48–53^. A range of 4-25 damaging gene variants were found per person.

### GEM Analysis

We also used GEM to identify gene variants in each subject that were most likely to be pathogenic^24^. GEM is an Electronic Clinical Decision Support System (eCDSS) framework that aggregates and adjudicates data from multiple algorithms and clinical datasets to provide rapid and accurate diagnosis of individual genomes^24^. GEM generates a Bayes Factor-based score that calculates the degree of support for and against a given model (a gene allele is pathogenic vs. benign) considering multiple lines of evidence from the following variant analysis tools and data sources: VVP, VAAST, Phevor, mode of inheritance for disease genes from Online Mendelian Inheritance in Man (OMIM), pathogenicity of variants (ClinVar), population specific allele frequencies (gnomAD), quality of variants and the overall genome (data) and quality of the genomic location (gnomAD)^25,26,42,44, 54–56^. Using this data, GEM identifies potentially pathogenic genotypes and evaluates support for their association with disease. Gene variants for each subject were considered candidates if they had a GEM score ≥ 1 (strong support for the model of pathogenicity)^57^, together with genes having a GEM score ≥ 0.69 (substantial support for the model of pathogenicity), and a Phevor Bayes factor ≥ 0.9 (genes with a strong association with POI)^54^. One to twenty-four gene variants were identified for each subject.

### Creating Categories for Enrichment Analysis

We used the Database for Annotation, Visualization and Integrated Discovery (DAVID) to functionally annotate known POI genes and candidate genes identified in model organisms ^14,58^. The analysis yielded 47 clusters with one removed for too few genes (<20; Cluster 37)(Supplementary Table 1). Cluster 37 contained mismatch repair genes, but the genes were also found in other clusters and was therefore redundant.

### Calculating Gene Burden on Resampled Gene Lists and Housekeeping Genes

We randomly selected 146 genes from a list of housekeeping genes that are constitutively expressed over many developmental time points in 16 tissue types (Supplementary Table 2). We ensured that none of the housekeeping genes were found in our gene sets identified in the GEM analysis. In addition, we created a burden-matched set of genes. For this, we created a burden ratio for every gene by summing the number of rare variants (MAF ≤ 0.005) in the longest coding transcript of each gene and dividing by transcript length. For each decile in the distribution of this burden ratio, we determined the mean and standard deviation of the burden ratio. We then used these mean and standard deviations to generate randomly sampled gene sets for each decile that had matched mean and standard deviation for burden ratio. These burden matched gene sets were used to test the significance of gene set enrichment in the subjects.

### Permutation Tests and Case/Control Comparison

For the permutation tests and analysis, we used the GEM results with a GEM score ≥ 1 generated from the POI cases and control individuals to test for enrichment in individuals with POI compared to controls. Both sets of data generated GEM results using two different HPO terms: POI (HP:0008209) and phenotypic abnormality (HP:0000118) to create 4 sets of data: Cases POI, Cases Phenotypic Abnormality (root), Controls POI, Controls Phenotypic Abnormality (root). The reason we ran GEM using the root of the HPO ontology (Phenotypic Abnormality) was to control for overly connected genes that might have inflated Phevor scores, thus reducing biases due to the nature of the ontology. We then determined the number of damaged genes found in GEM results (number of successes) from genes listed in the individual pathways, a burden-matched gene list and a housekeeping gene list.

Permutation analyses were performed using a random sampling strategy to evaluate enrichment of the POI dataset against gene lists related to functional aspects of the disease. A gene list containing 18,876 RefSeq genes was first created, excluding mucins and olfactory receptor genes. For each functional gene list of size N, random samples of equal size were drawn from the 18,876 genes. This process was repeated 100,000 times, each time with an independently generated random gene list, to create an empirical distribution of the number of damaged genes (GEM score ≥ 1) for each functional gene list. To test whether the probands show enrichment in the functional gene lists, the actual number of damaged genes found for each list was compared to the distribution of damaged genes found using burden-matched gene lists and the housekeeping genes list. To test for statistical significance, we used Fisher’s exact test to calculate a *p* value using the 2×2 contingency table testing the hypothesis that the number of damaging genes that matched the functional list was significantly larger in the POI GEM runs than in the root Phenotypic Abnormality GEM runs. All *p* values were adjusted for multiple testing (False Discovery Rate, FDR). To generate a final score depicting the most significantly enriched pathways, adjusted POI *p* values were divided by Phenotypic Abnormality *p* values to generate a normalized score that represents enrichment. The higher the ratio, the more enriched the pathway. Pathways with a corrected *p* value <0.05 and a log_2_ ratio of greater than 2 for the POI *p* value/Phenotypic Abnormality *p* value were considered significant pathways.

### Oocyte Expression

To determine whether candidate genes are expressed in mammalian oocytes, 35 day-old female mice were treated with an intraperitoneal injection of 5 IU PMSG to initiate follicular development and 5 IU hCG 48 hours later to induce ovulation^60^. Eighteen hours later, mice were sacrificed, oviducts dissected to remove oocytes and cumulus cells manually removed. RNA was isolated from oocytes using RNeasy (Qiagen, Valencia, CA)^9^. Reverse transcription was performed with SuperScript Master Mix (Life Technologies, Carlsbad, CA) using SuperScript III RT and random primers. Quantitative real-time polymerase chain reaction was performed for the expression of candidate genes and glyceraldehyde-3-phosphate dehydrogenase (*Gapdh*) as an endogenous control using PowerUp SYBR Green Master Mix (Applied Biosystems, Foster City, CA). Primers were designed to span two exons to avoid amplifying genomic DNA. Primer sequences are provided (Supplementary Table 3). Samples were examined in triplicate and at three dilutions. mRNA levels were determined using the 2^-ΔΔCT^ method to calculate relative quantification and to correct for expression of endogenous controls.

### Functional Analysis

Flies were raised at 25° C on standard diet based on the Bloomington *Drosophila* Stock Center standard medium with malt. We obtained 20 RNAi lines from the Bloomington *Drosophila* Stock Center. Ovary/germline specific RNAi knockdowns were performed using Gal4 DNA-binding protein and Upstream Activator Sequence (*GAL4/UAS*) technology, as previously described^61^. We crossed flies carrying the *Maternal Triple Driver-GAL4* (*MTD-GAL4*; BDSC 31777) transgene to flies carrying each respective UAS-RNAi transgene to generate female flies with ovary specific knockdown of each gene. Control flies were generated by crossing flies carrying the *MTD-GAL4* transgene to the appropriate *AttP* RNAi background strain (does not carry UAS-RNAi transgene). Virgin female RNAi knockdown (and control) flies were collected on CO_2_ anesthesia and aged 3-5 days on standard media supplemented with dry yeast. Female knockdown flies were singly mated with a 3-5 day old Canton S male. Individual mating pairs were observed to ensure successful mating. Males were removed after mating. We measured four female reproductive phenotypes: 1) egg number: number of eggs laid in first 8 hrs post mating; 2) hatchability: number of adults that hatched from those eggs; 3) total fertility over 10 days post mating; 4) overall ovary appearance and morphology^62^. For egg number, newly mated females were place in vials for 8 hrs and egg number was counted. For hatchability, all the progeny that eclosed from the egg number vial were counted (progeny #/egg #). To measure total fertility, mated females were transferred to new vials every two days for ten days and all the progeny were summed over the entire period. For ovary images, adult females were collected under CO_2_ anesthesia, dissected and immediately imaged. Ovaries were imaged at 3X magnification using a Leica EC3 camera. We assayed at 8-10 females per RNAi knockdown. Statistical analysis was performed using R software. P-values were determined using ANOVA. A *p* value < 0.05 was used for significance.

## Results

Whole exome sequencing produced a mean of 96 million reads per individual (range 31-186 million) and an average of 99.6% mapped/aligned reads to the GRCh37 human reference genome with an average duplication rate of 8.4% (Supplementary Figure 1A-C) for the 280 samples that passed QC metric cutoffs. Fastp identified read quality and insert sizes within the normal ranges. Variant calling produced an average of 21,923 SNVs and 576 indels per sample, with an average depth of 55x per sample (Supplementary Figure 1G-I). Peddy was used to infer the sex, heterozygosity, ancestry, and relatedness of subjects, and compared to known metadata about samples (Supplementary Figure 2A-D)^41^. From these quality control metrics we identified and removed four samples (Supplementary Table 4) that had very low heterozygosity and low coverage. Three samples were removed due to high duplication rates. One sample was removed due to excess heterozygosity. We also discovered a previously unidentified deletion of the long arm of the X chromosome (93.7% homozygous X:130678467-X:155171537) in sample IPOF32. In total, we removed 9 samples from analysis for the quality issues described above leaving 282 samples for the analysis. Peddy confirmed the sex of the POI subjects (Supplementary Figure 2A), and the known relatedness of a few individuals (Supplementary Table 5, Supplementary Figure 2D-E). An additional four sib pairs were identified in the cohort and one family with dominant inheritance was included (Supplementary Figure 2E). For these related individuals, only one subject was included in the joint analyses. PCA projection of the samples together with data from the 1000 genomes identified individuals of European descent (n=235), admixed American (n=18), African (n=10), South Asian (n=4), East Asian (n=3), and unknown (n=12) ancestry (Supplementary Table 6, Supplementary Figure 2C).

In 19 subjects, we identified variants in genes previously determined to cause POI, including confirmation of previously identified variants in 12 subjects with primary amenorrhea (6.7%; Table 1)^19,63–66^. Five of these variants were found as heterozygous genotypes in the genes *NR5A1, PTPN22* and *eIF4ENIF1*^9,17,67^.

**Table 1.**
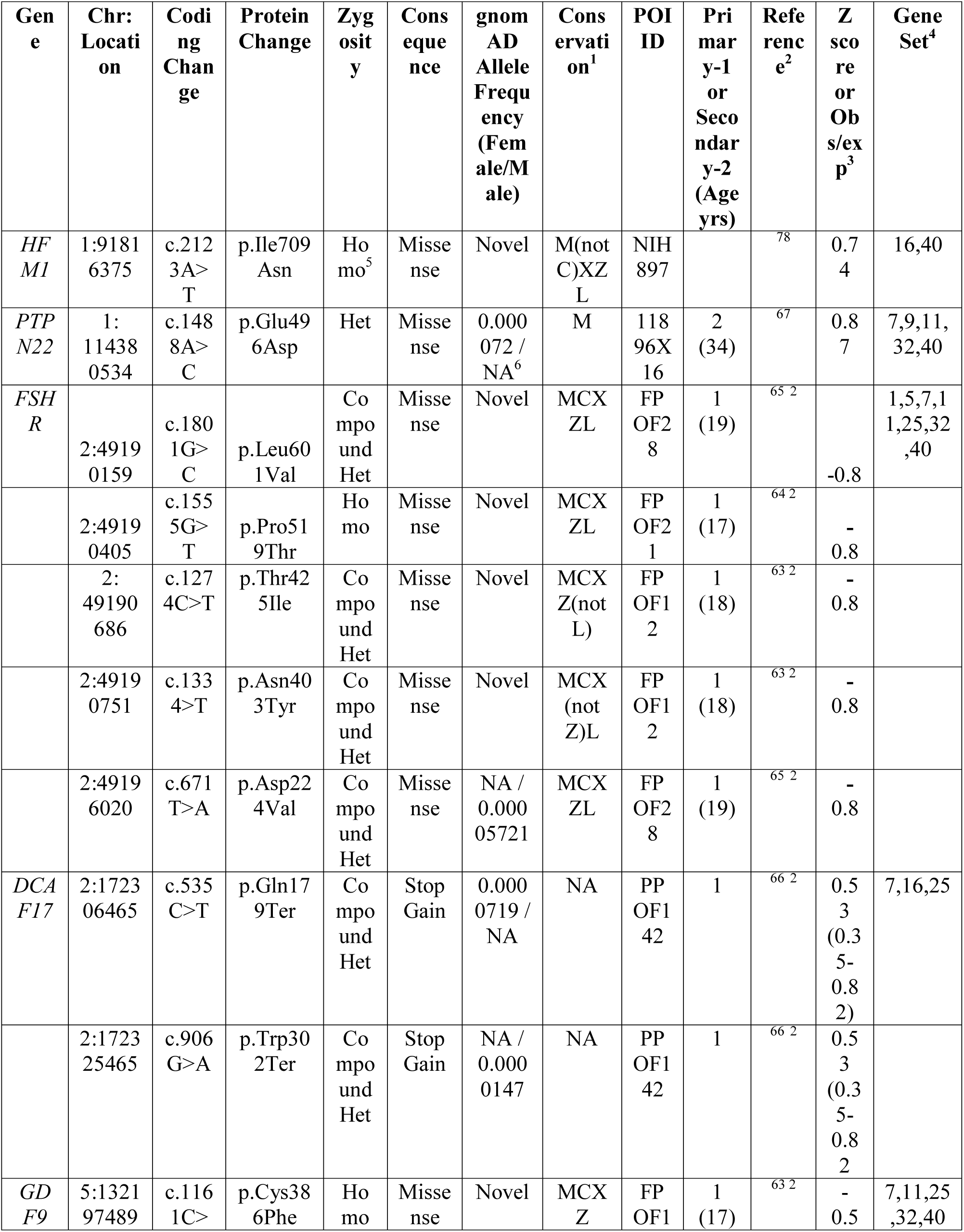
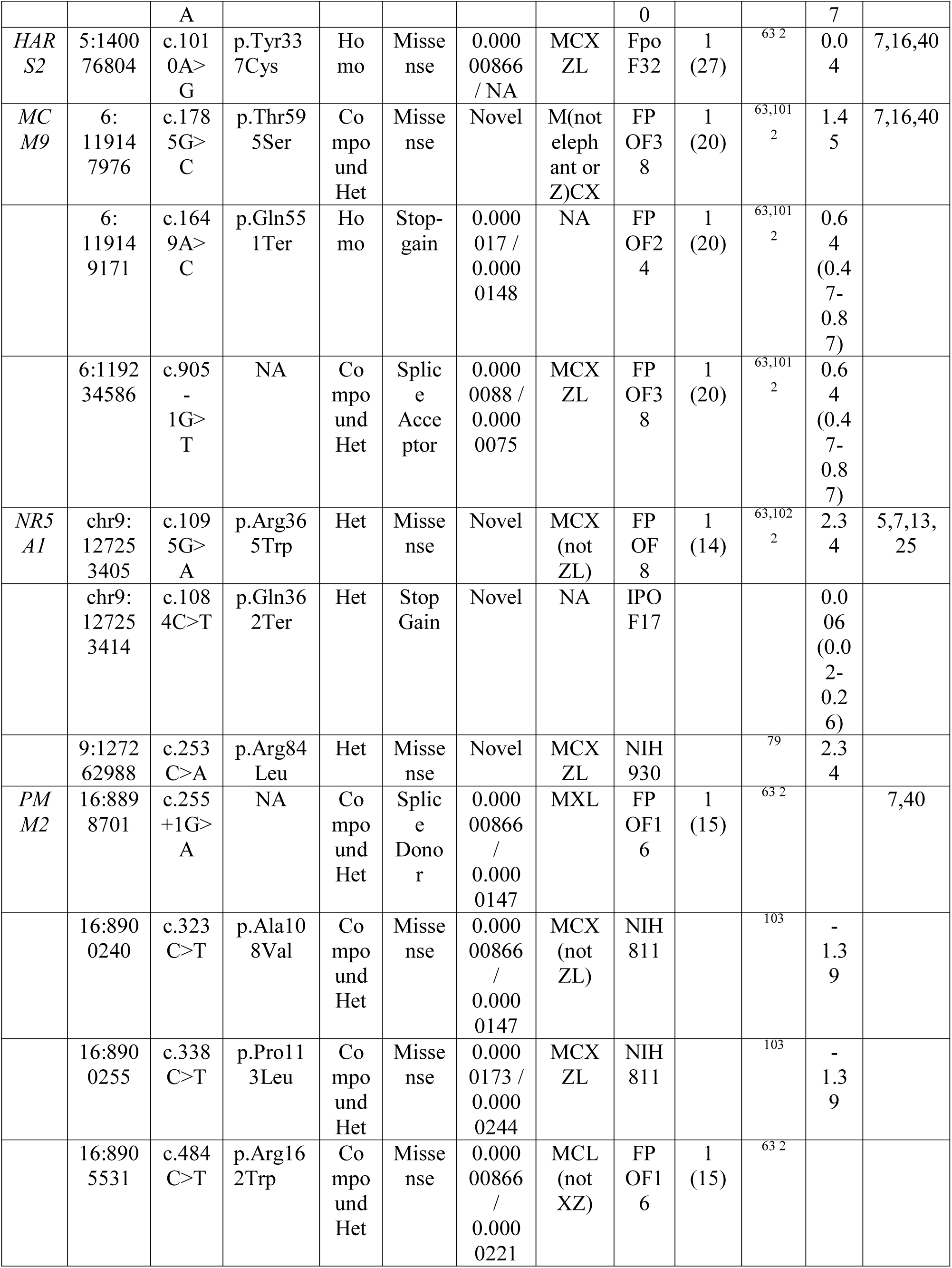
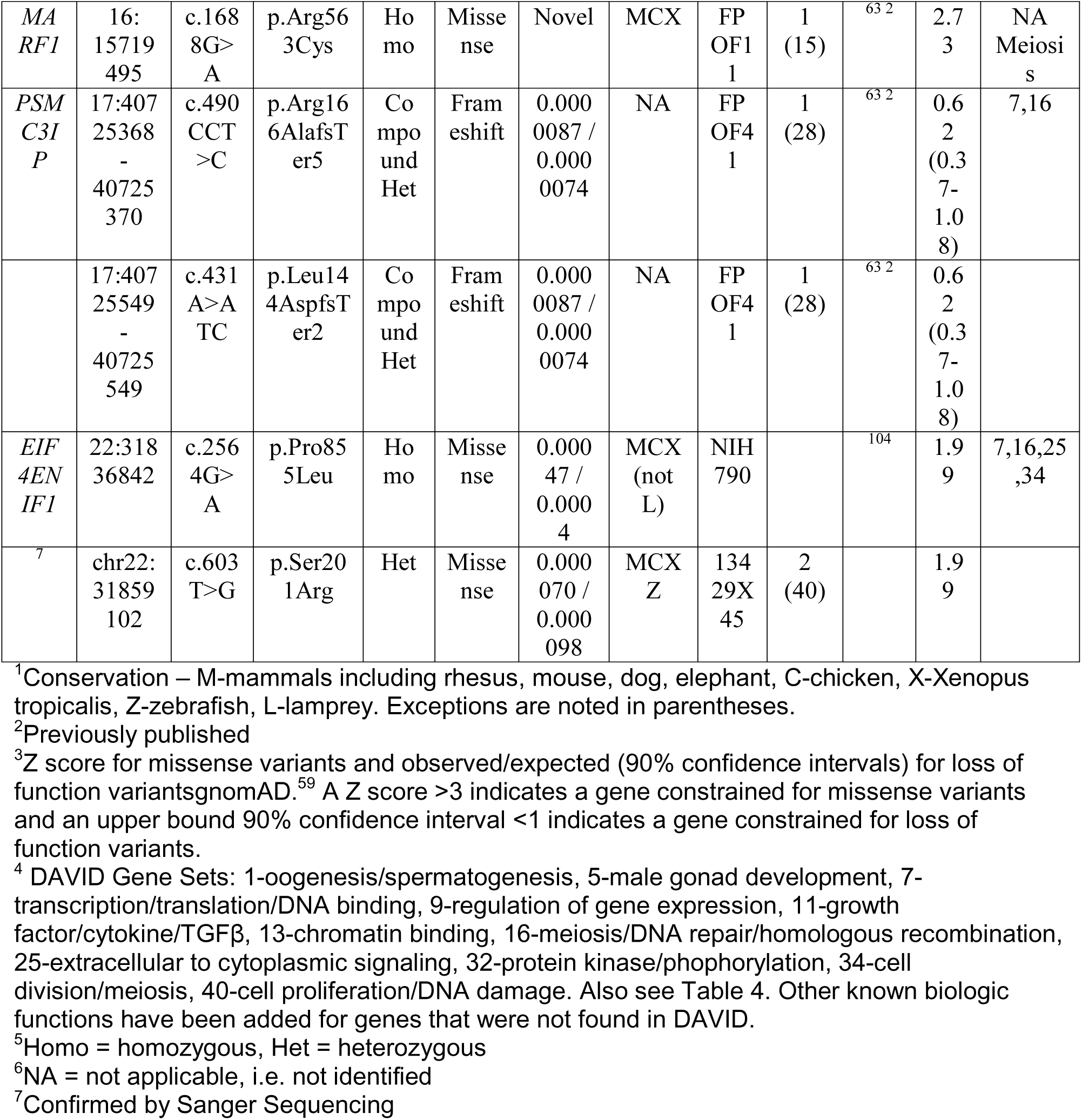
Candidate variants in previously identified genes causing POI.

Sixty-four subjects (23%) carried at least one variant in a previously identified POI gene that was determined to cause ovarian dysgenesis or primary amenorrhea with autosomal recessive inheritance (Table 2). Twenty-seven subjects (10%) carried a heterozygous variant in a gene for which there was a previously identified functional model (Table 3)^68,69^. Variants at genomic loci that were not conserved across species were not considered for analysis, although variants impacting conserved amino acids that were found only in mammalian species were included. One subject carried two variants in *FANCM* and one subject carried two variants in *RECQL4,* however, it was not possible to confirm whether these variants were in *cis* or *trans*. Fourteen subjects carried 2, one subject carried 3 and one subject carried 4 candidate POI risk variants in different genes.

**Table 2.**
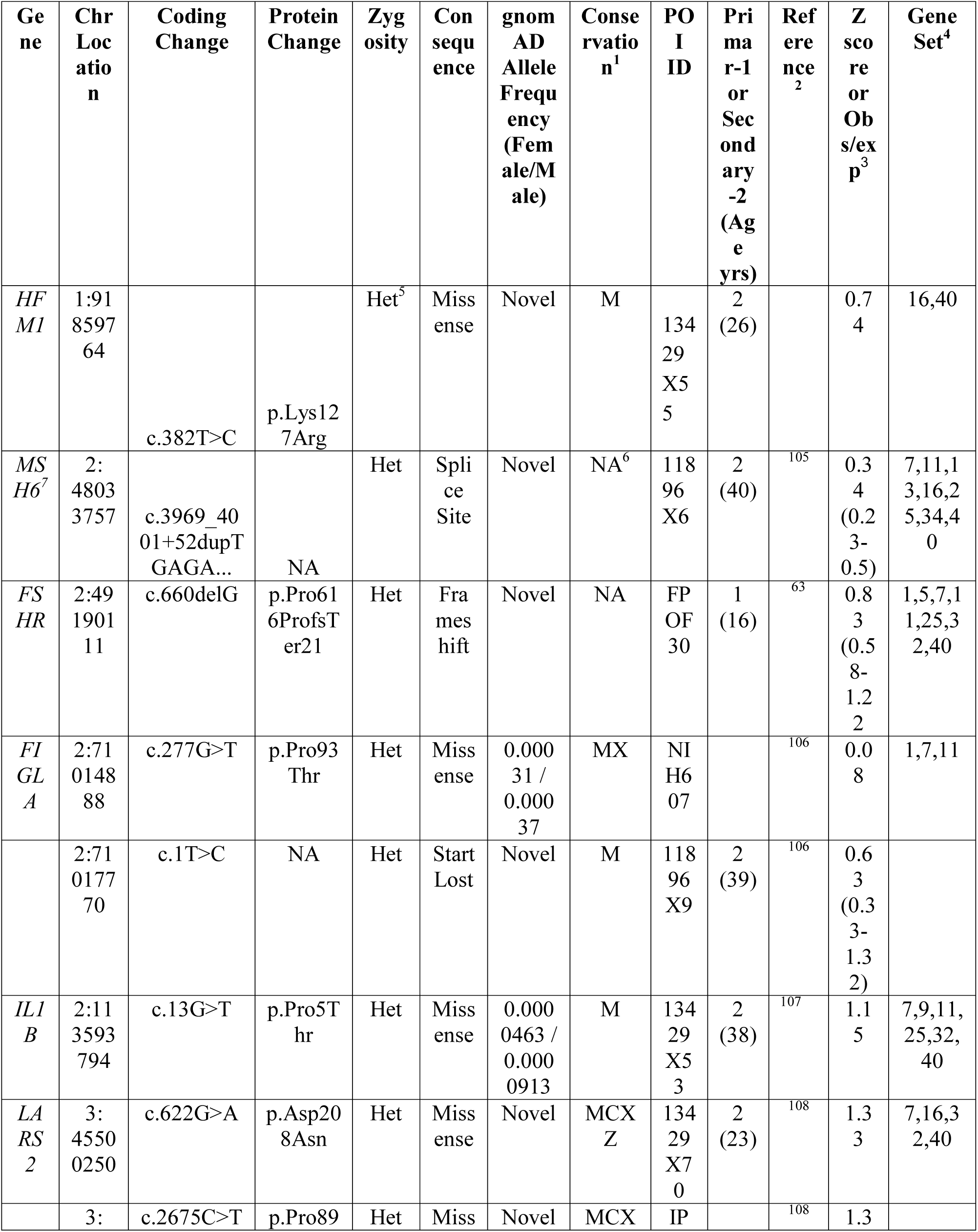
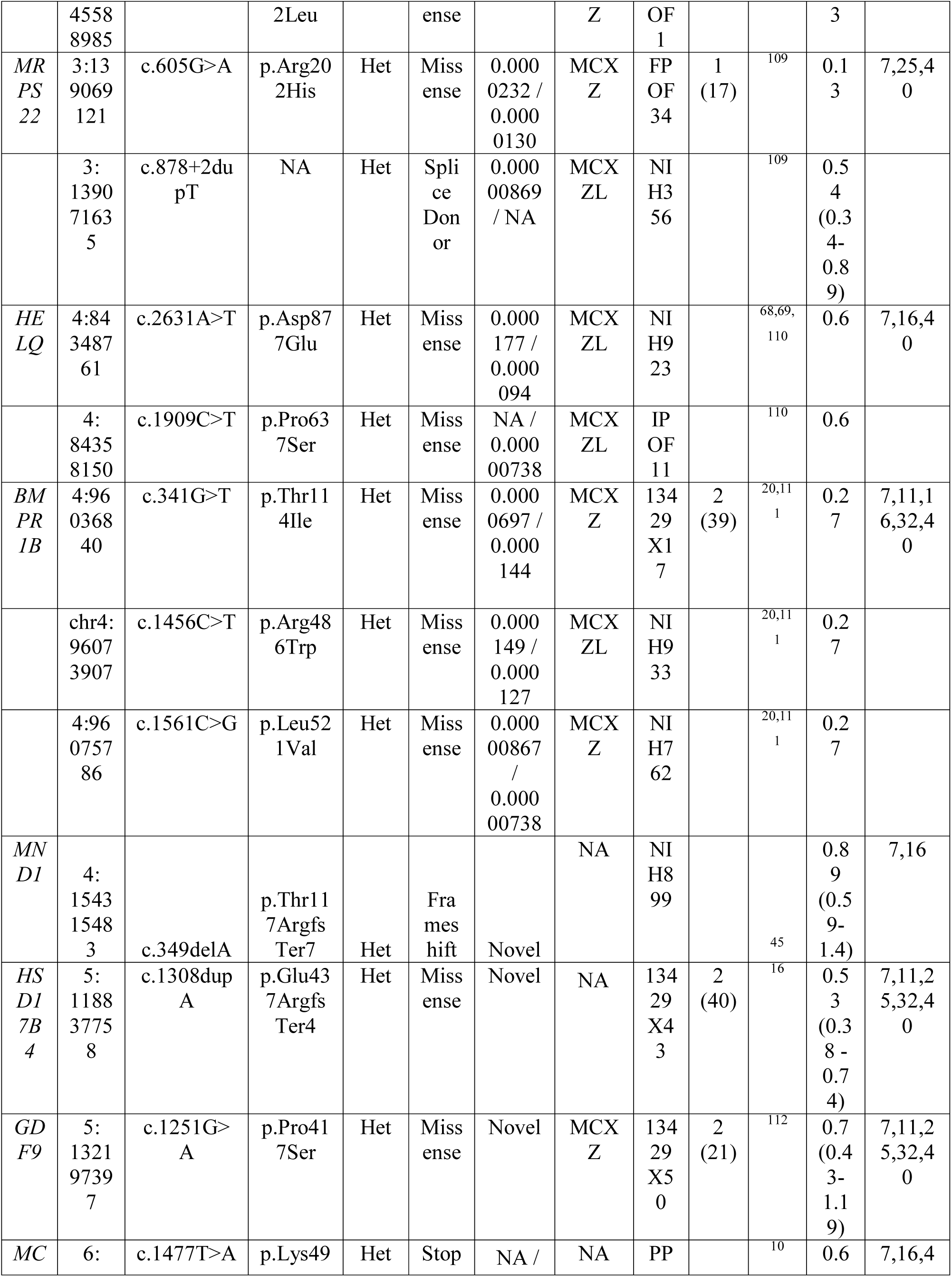
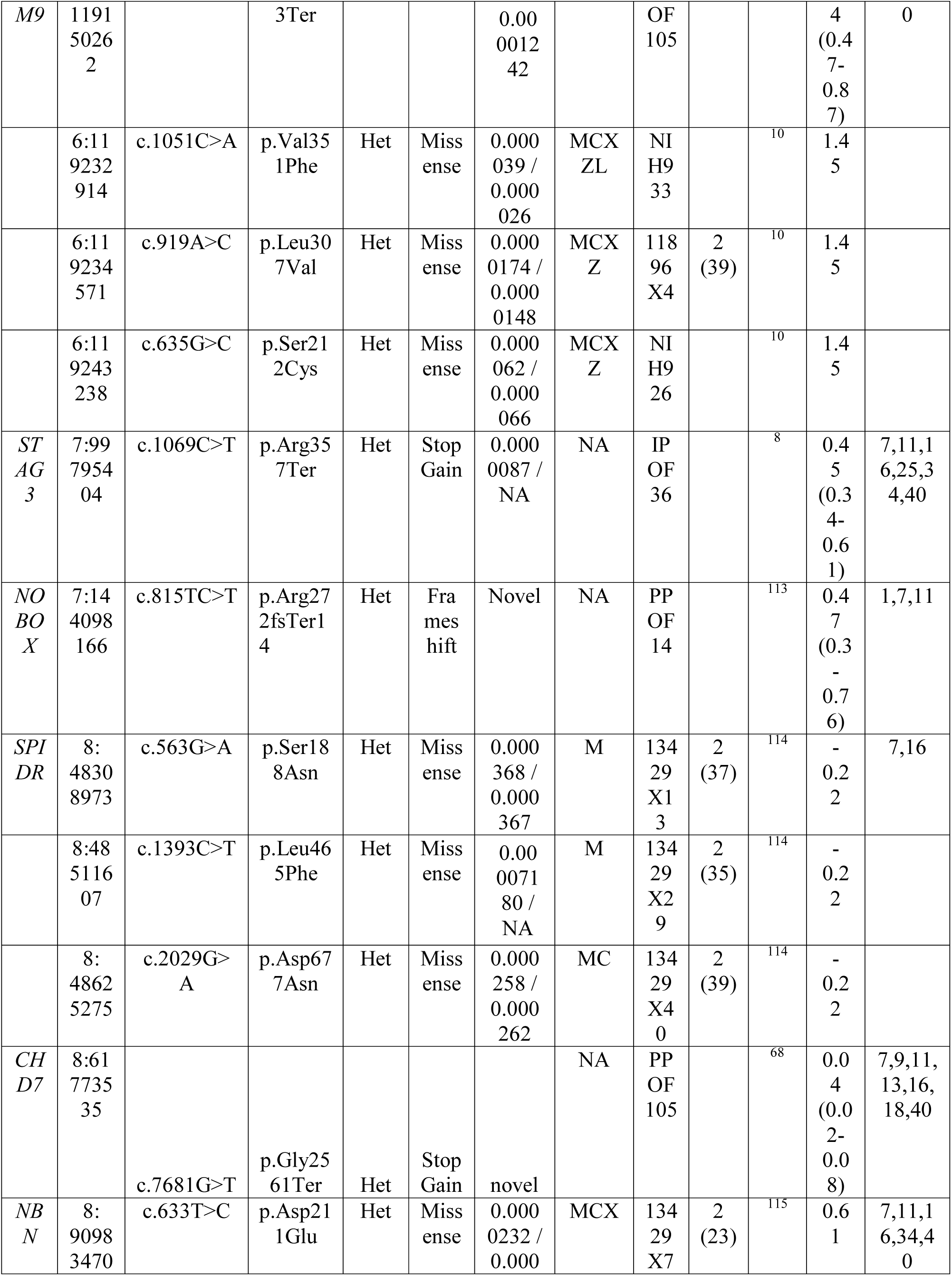
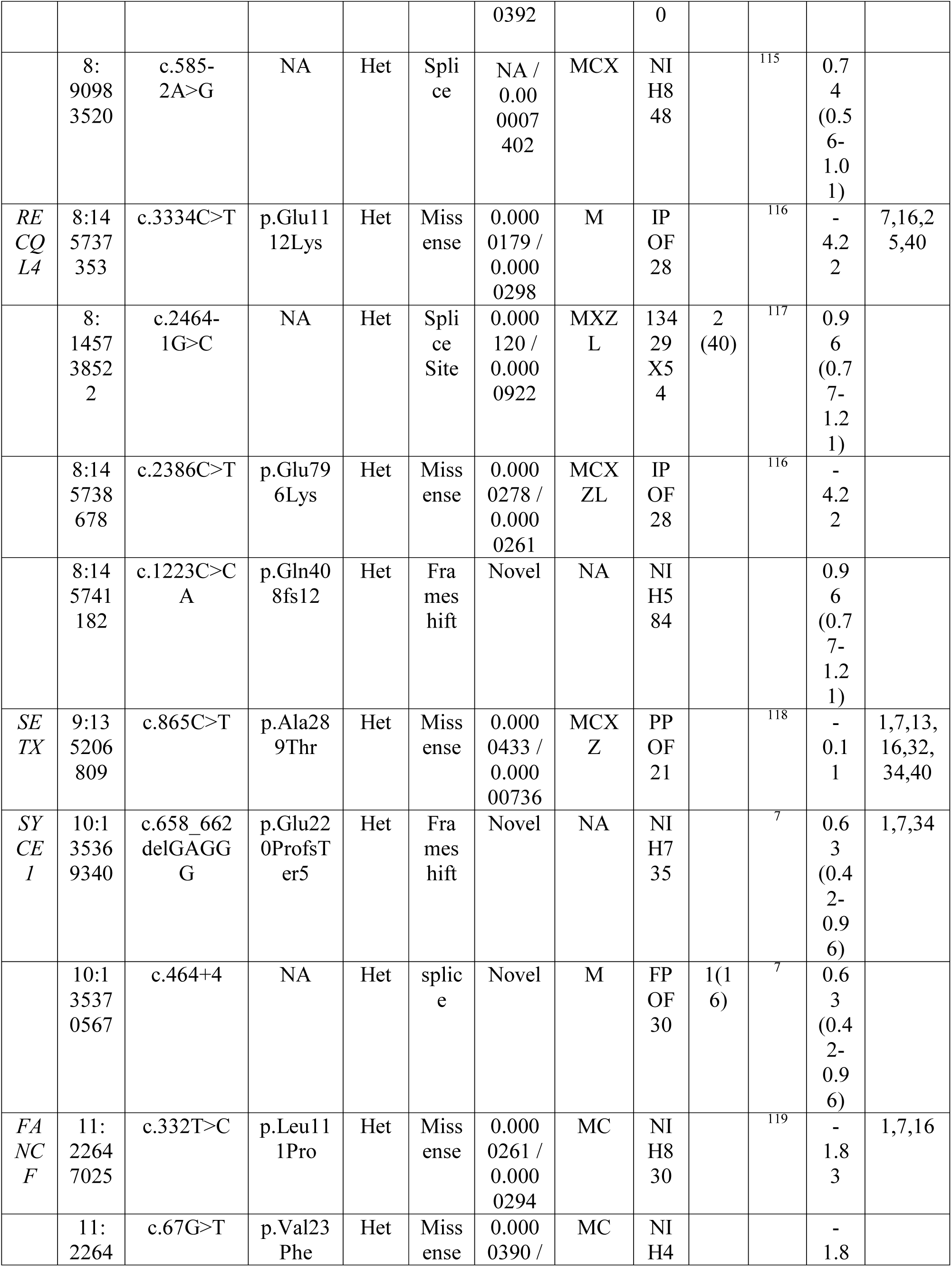
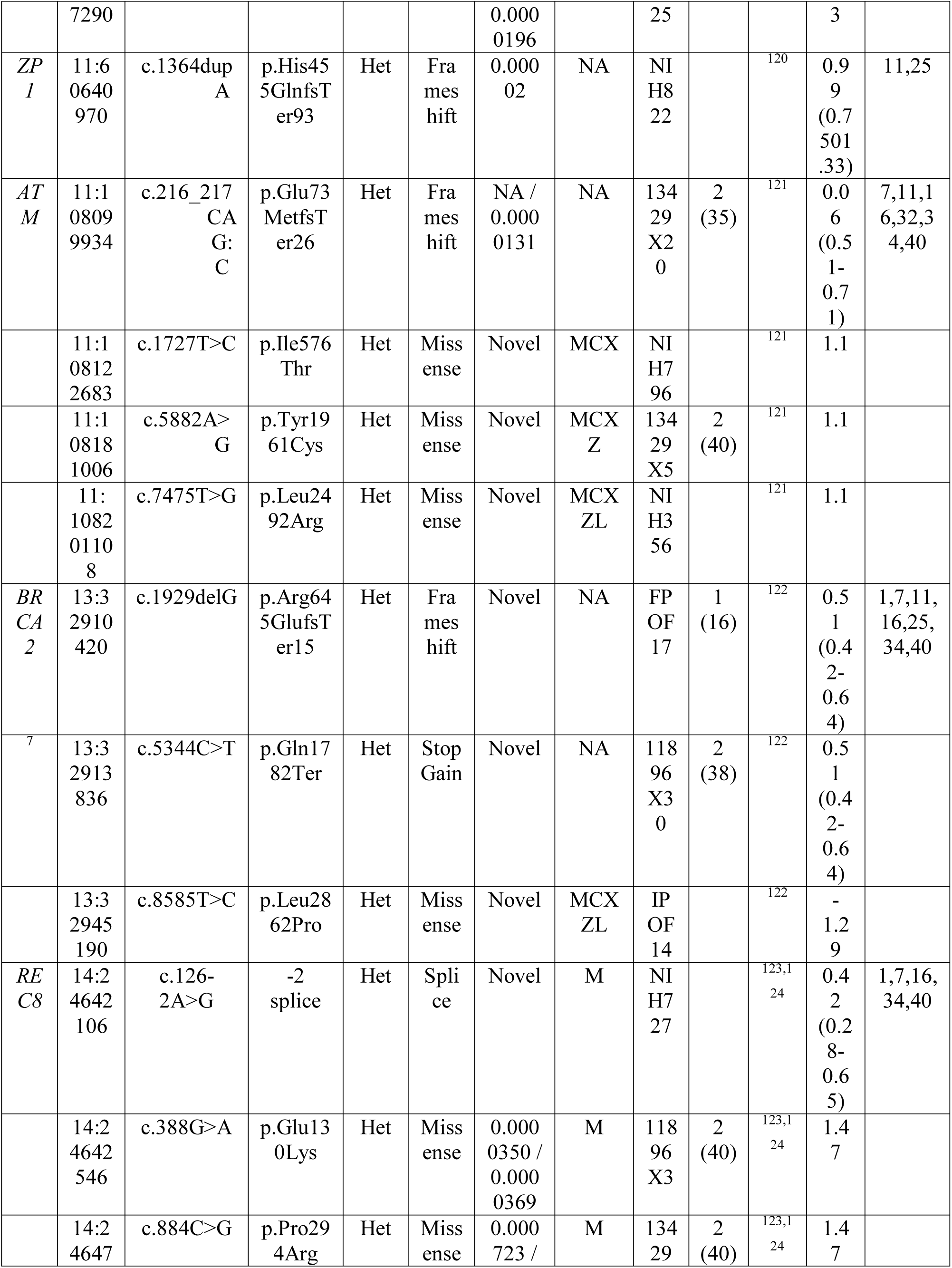
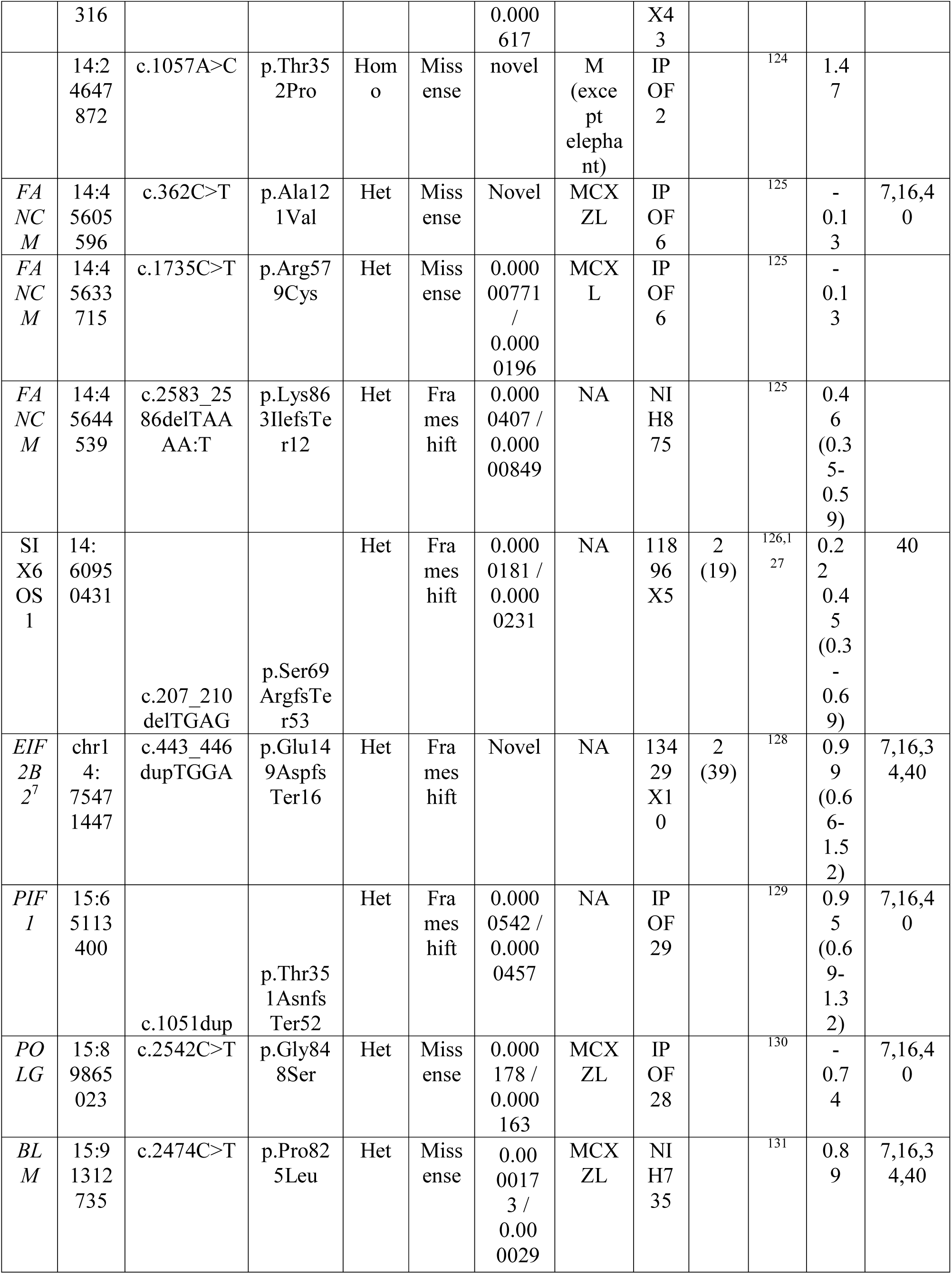
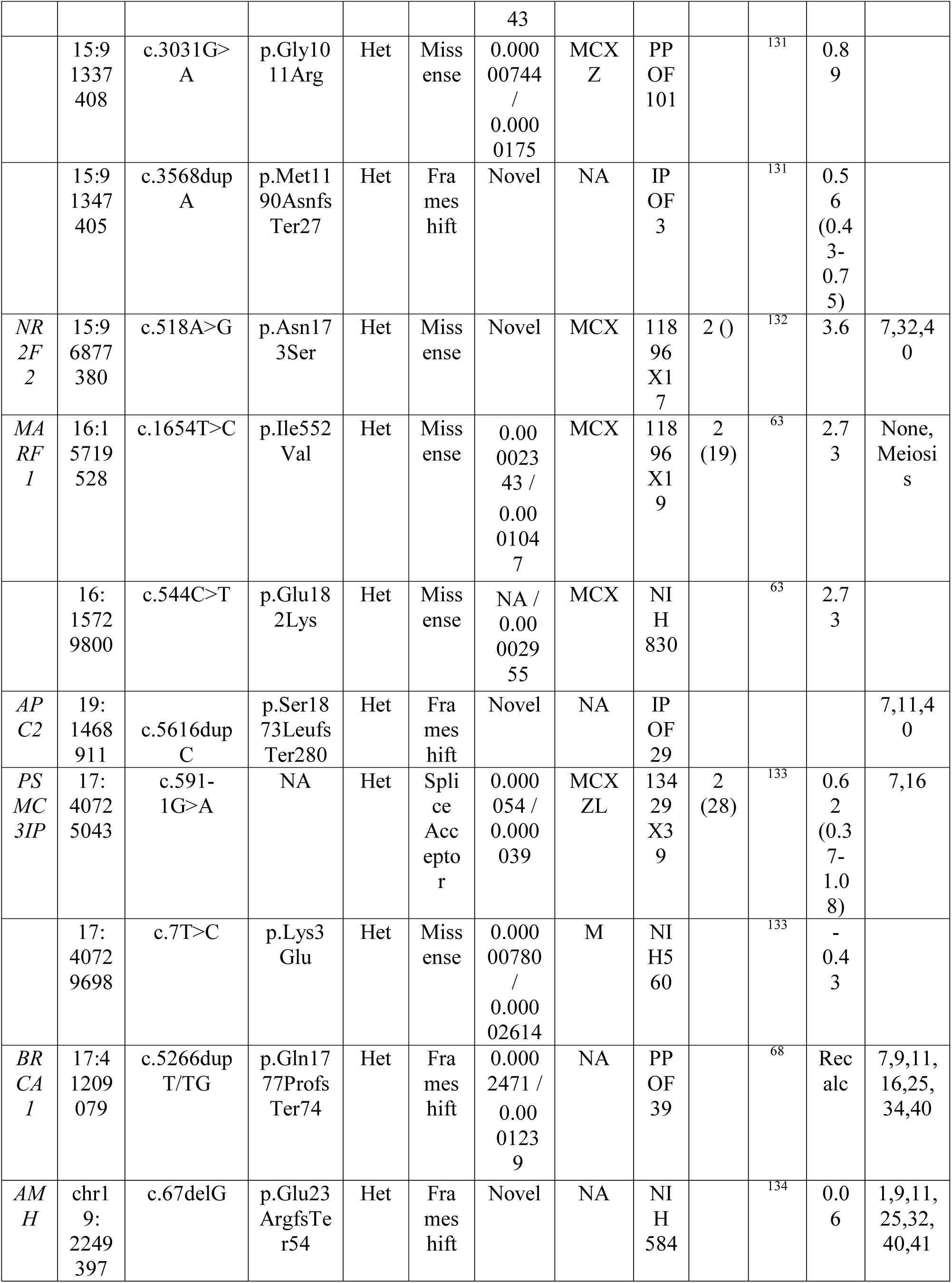
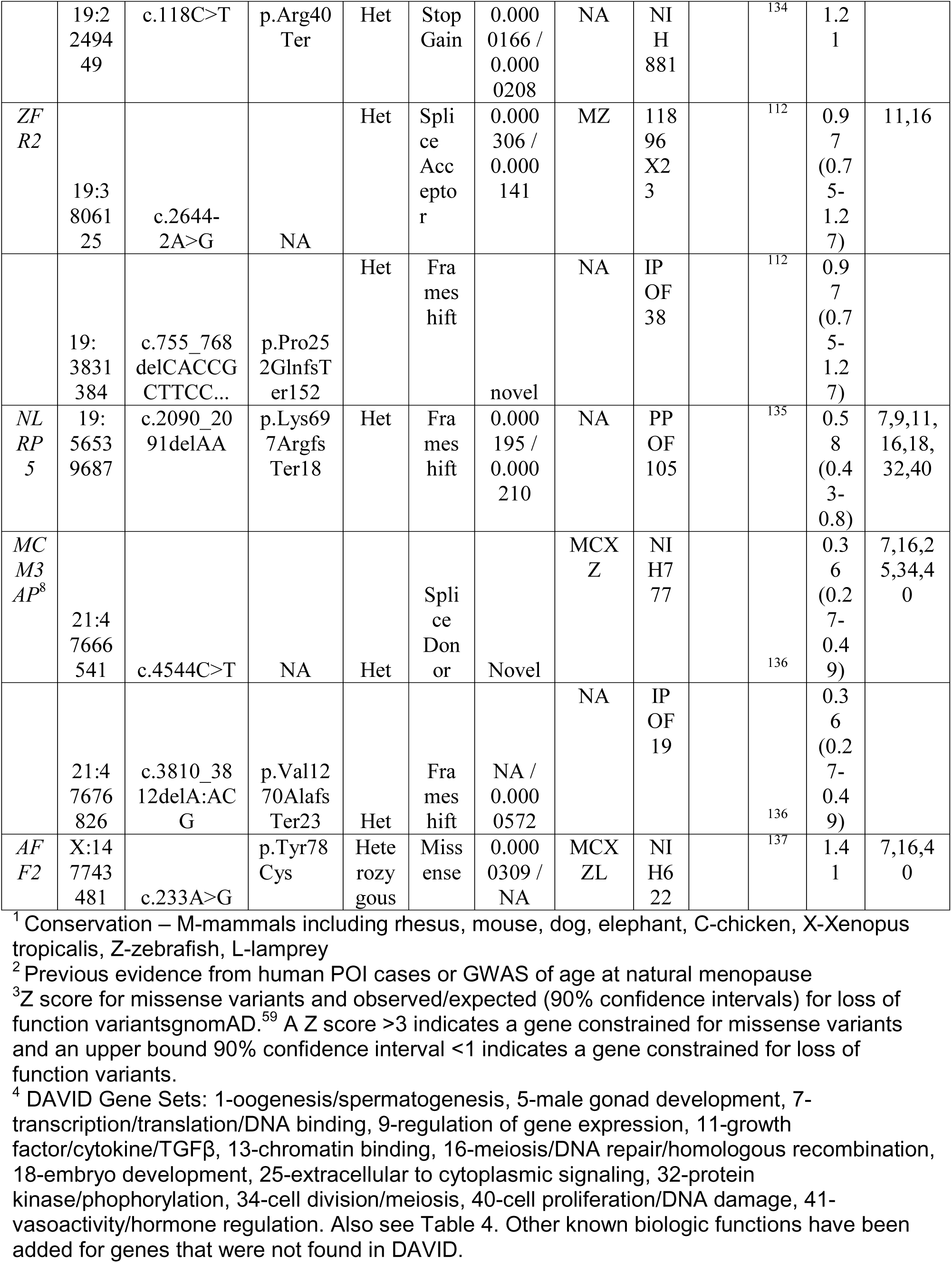
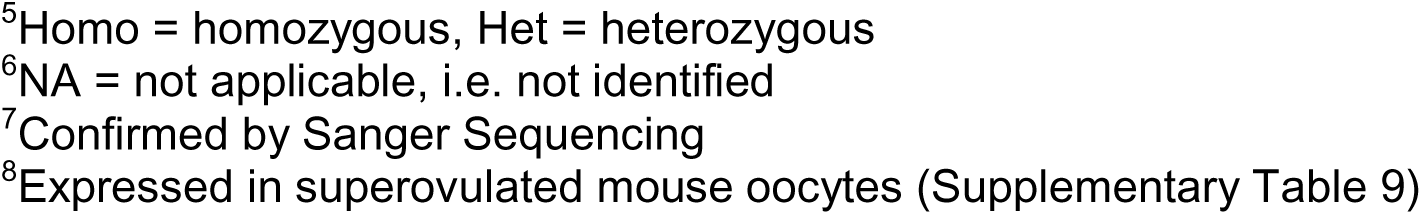
Candidate variants found as heterozygotes in previously identified genes causing POI or associated with age at natural menopause.

**Table 3.**
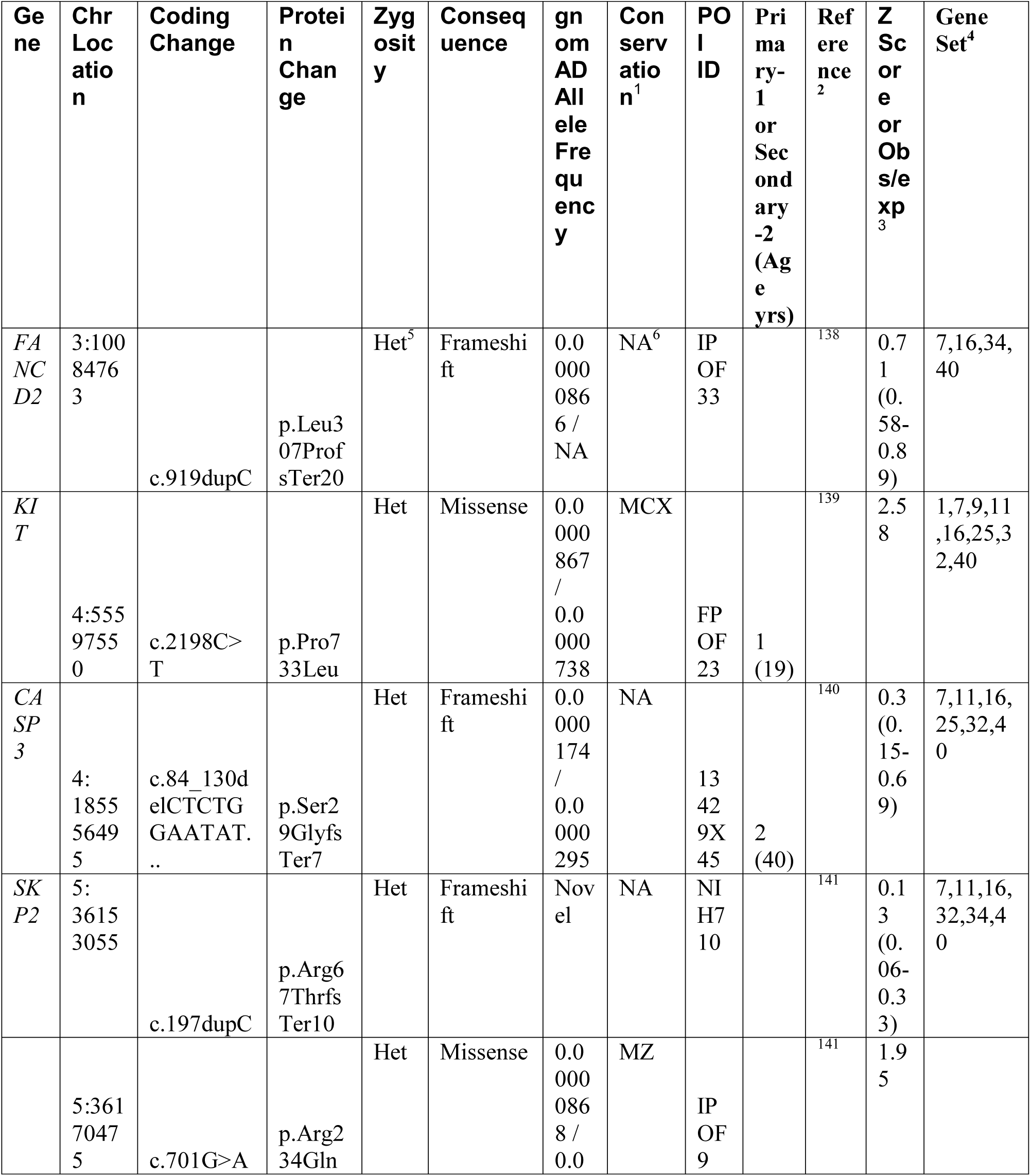
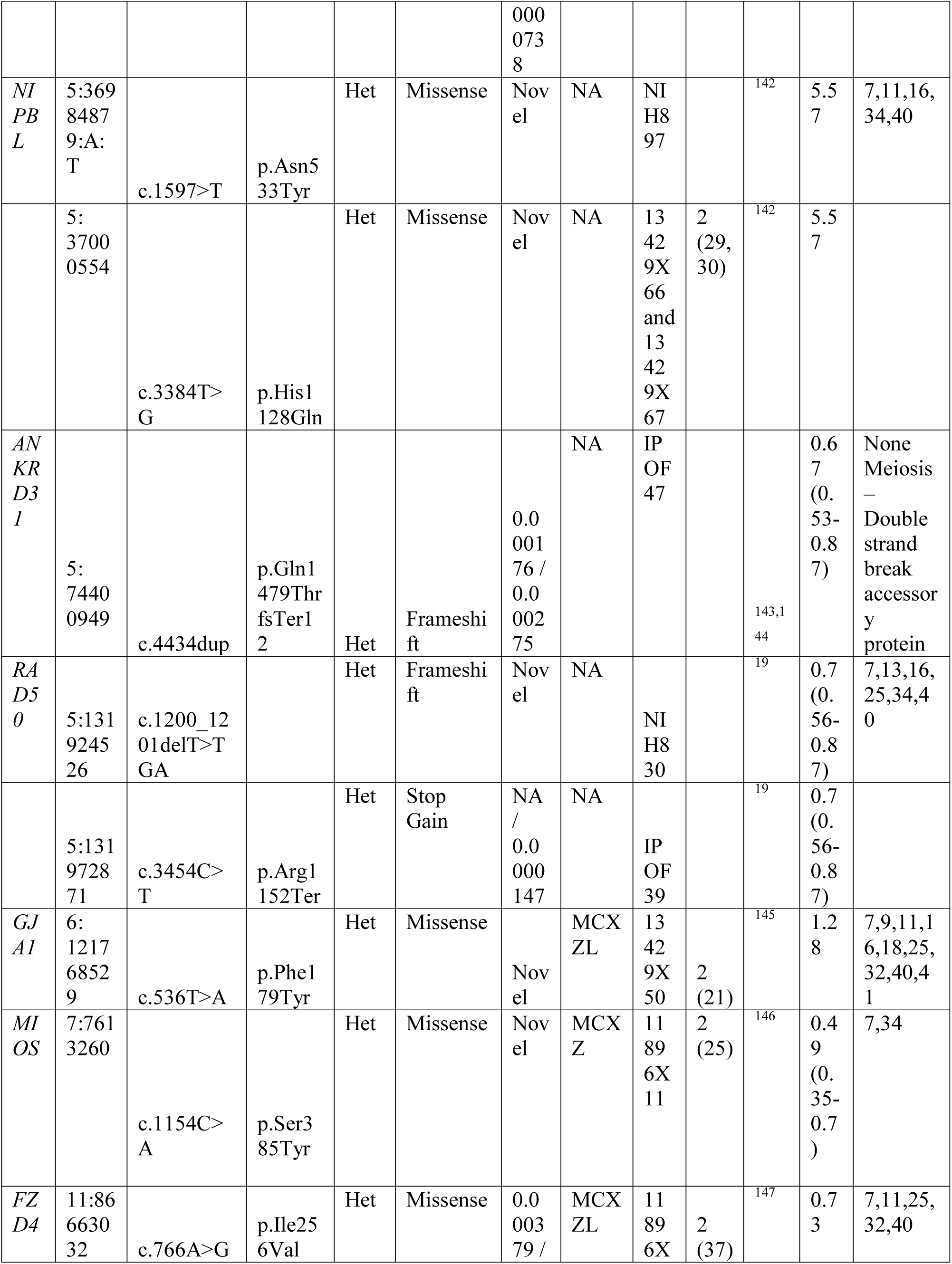
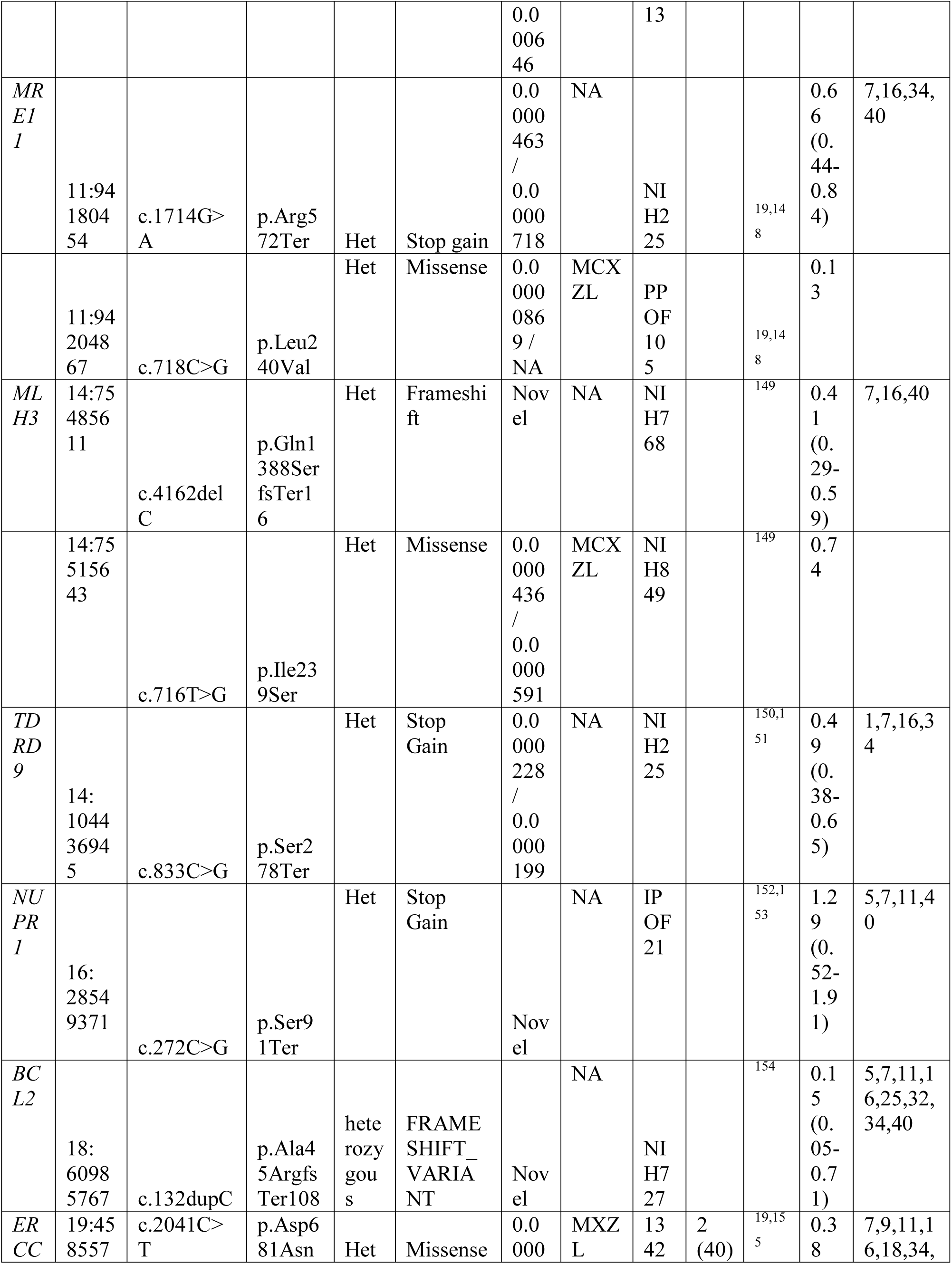
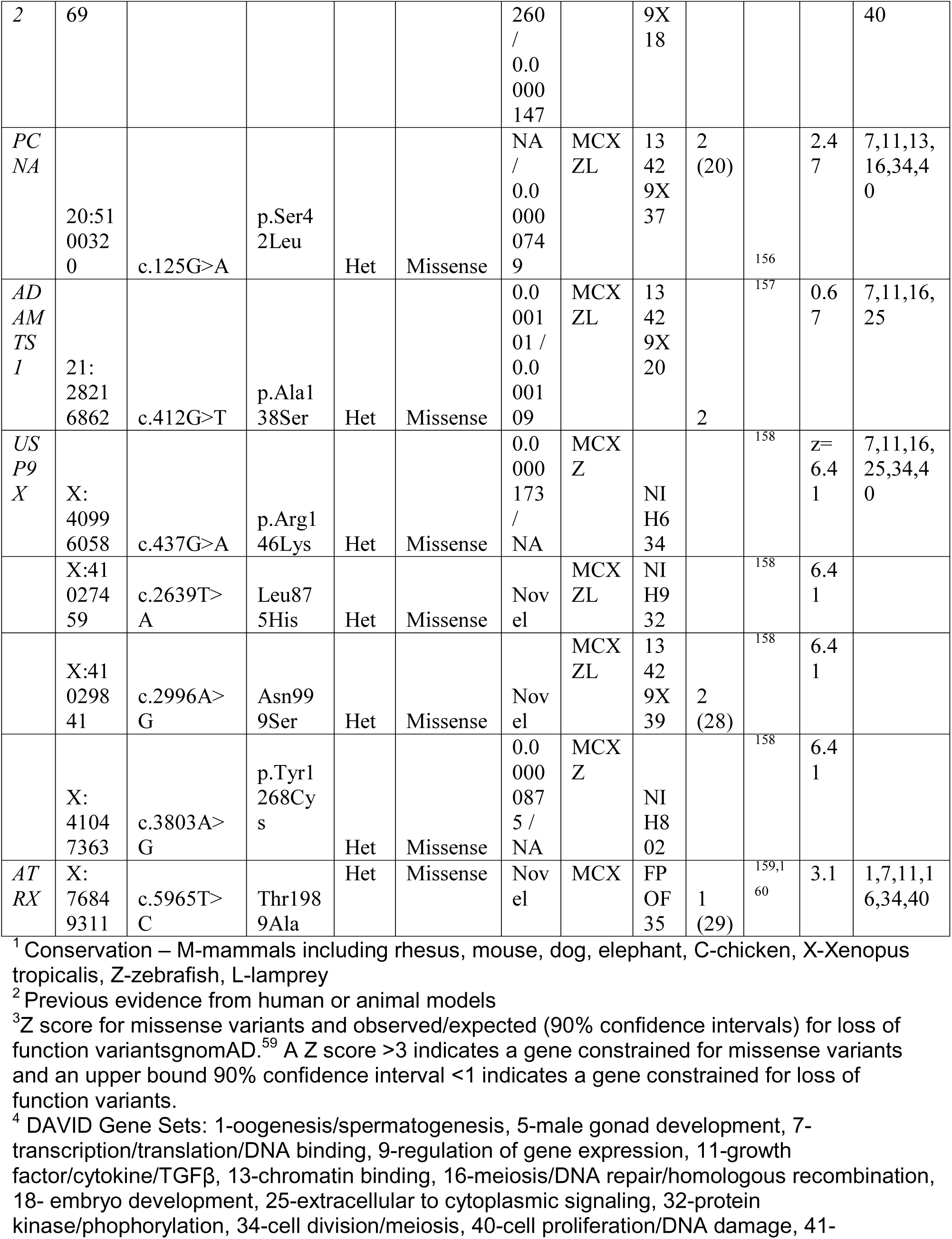
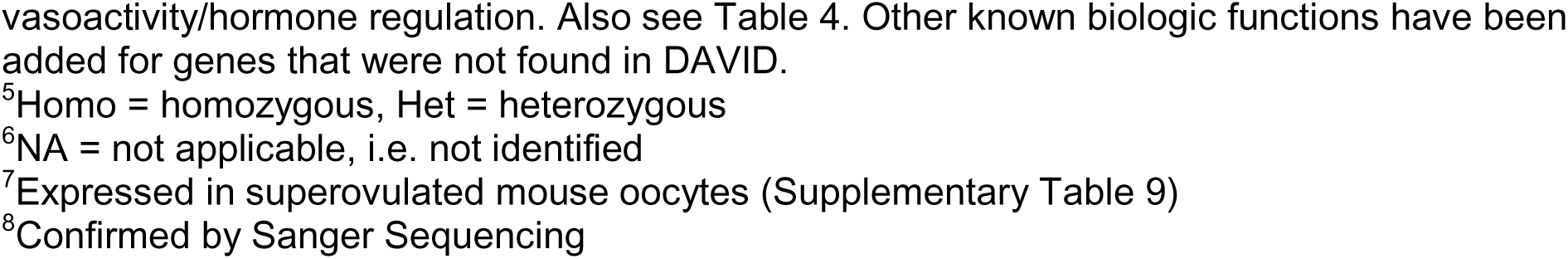
Candidate variants in genes from pathways with functional models

We next determined gene clusters for known genes for POI in women and candidates from animal models. DAVID analysis identified 47 gene list clusters with enrichment >2 (Supplementary Table 1). We then examined enrichment of these 47 gene sets in women with POI compared to controls and found 13 significant gene sets. These gene sets encompassed GO term biological processes including transcription/translation, DNA damage and repair, oogenesis, cell proliferation, hormone regulation, growth factors, regulation of gene expression, embryogenesis, cytoplasmic signaling, male gonad development, chromatin binding, cell division and protein phosphorylation (Table 4, Figure 1). Further, there was significant enhancement compared to housekeeping genes and burden-matched gene lists in POI cases compared to controls (Figure 1A-C and Supplementary Table 2). The majority of the causal or candidate genes were found in the enriched gene sets (Tables 1-3). The two genes that were not found in the gene lists are important for meiosis (*MARF1* and *ANKRD31*).

**Table 4.**
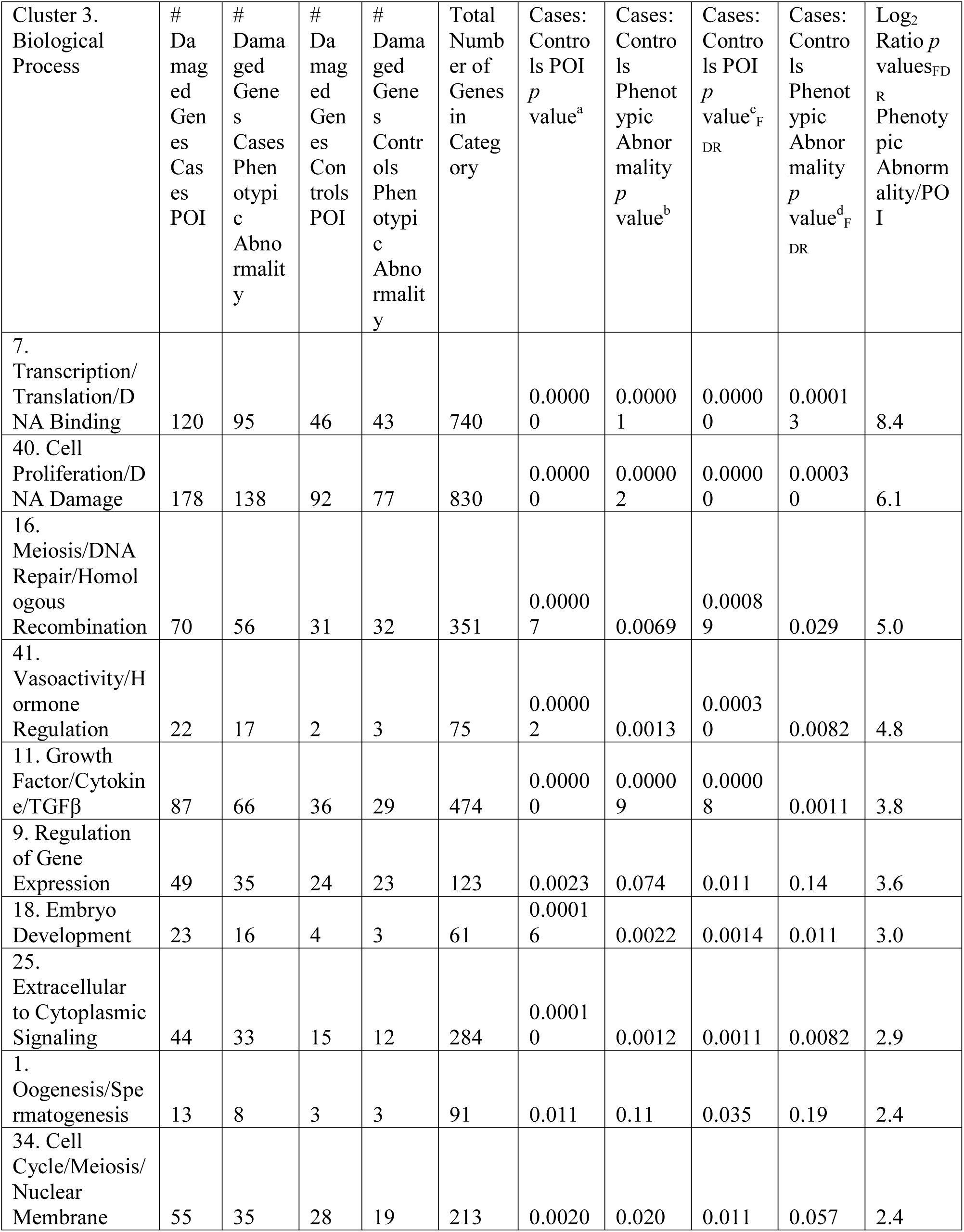
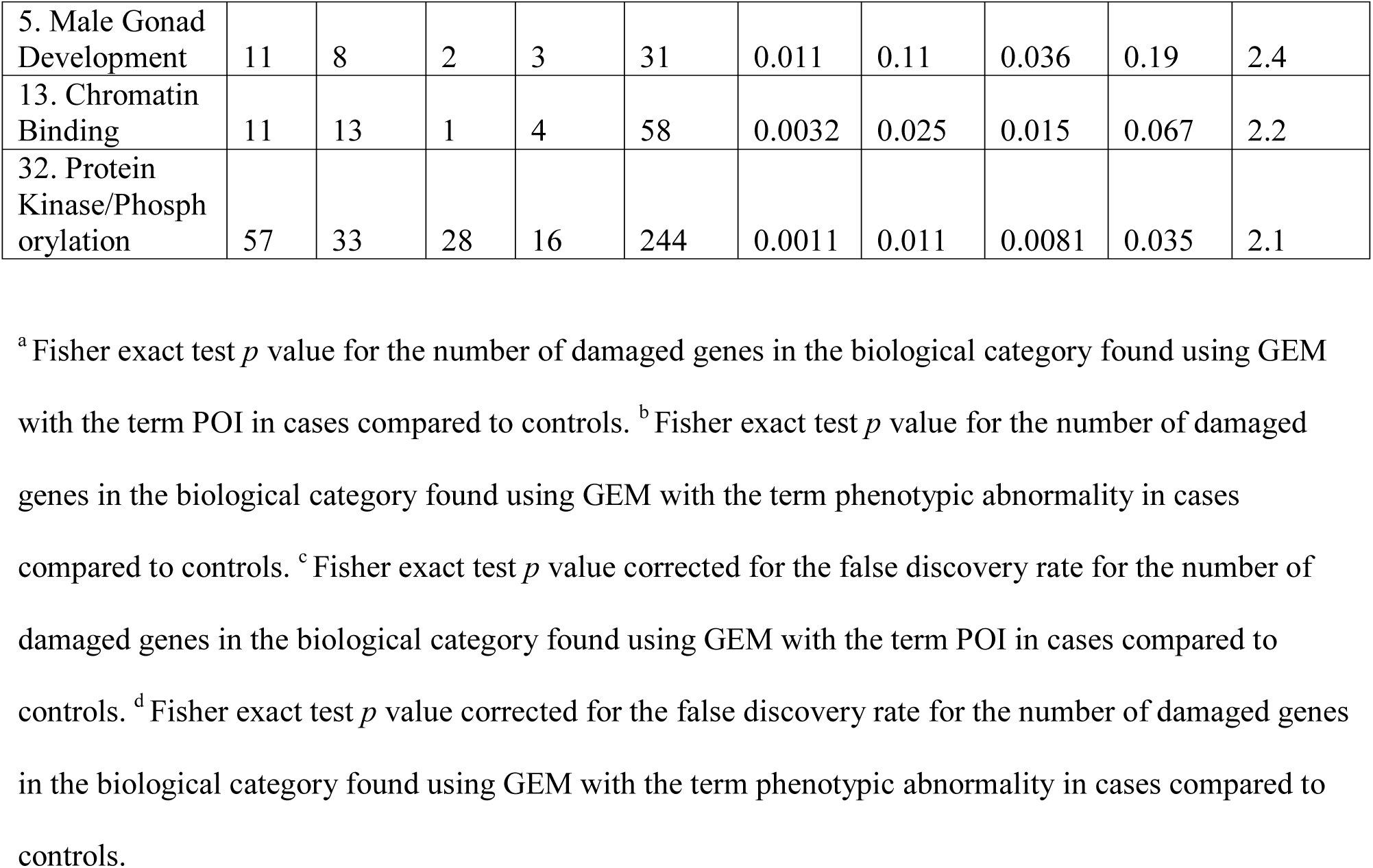
Enhanced biological pathways or clusters in women with POI compared to controls.

**Figure 1.**
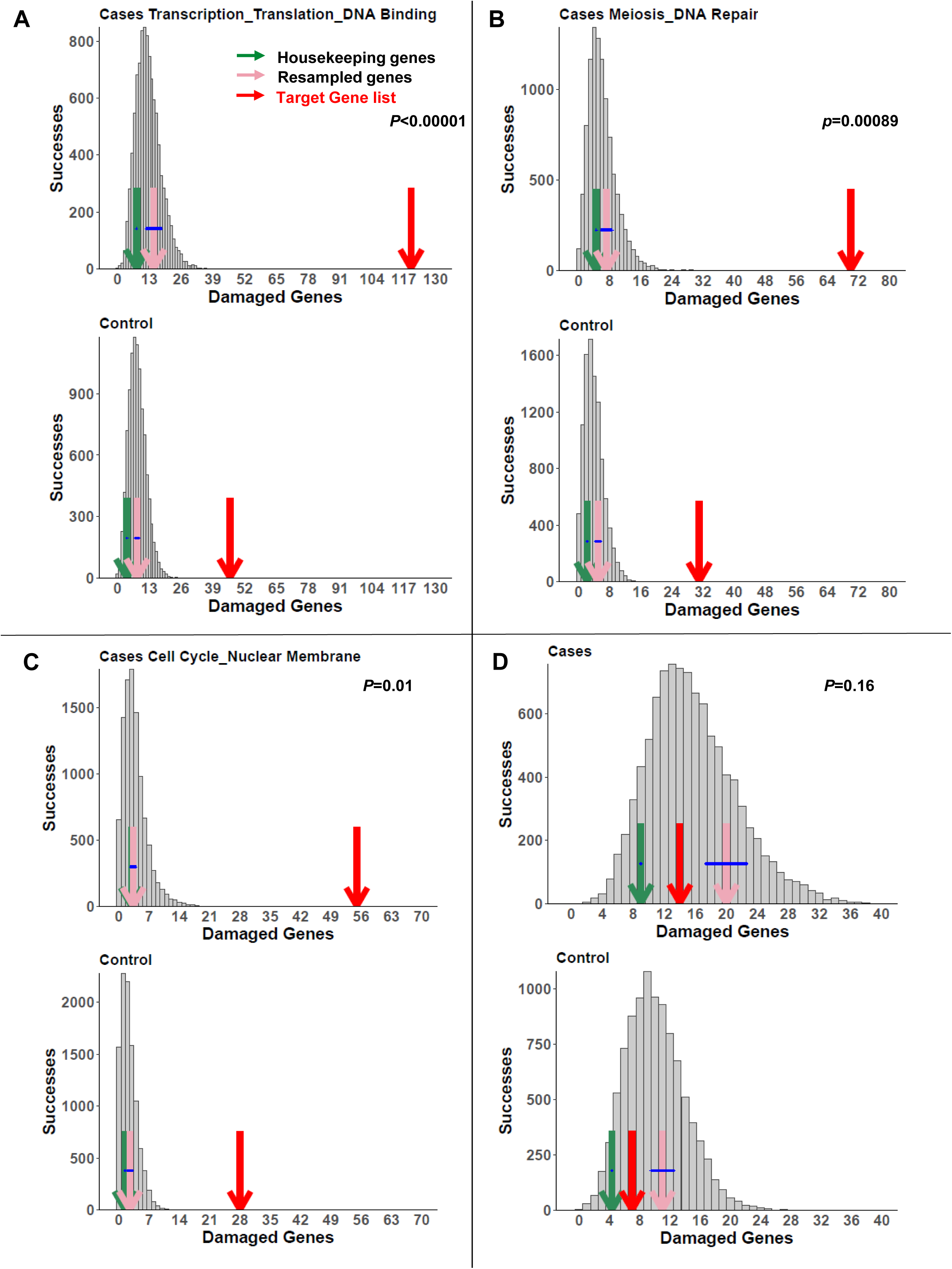
Three examples of enriched pathways in the POI data set as determined by the permutation tests in cases (upper panels) and controls (lower panels). Enriched pathways that encompassed novel POI genes (Table 5) included: A) Transcription/Translation/DNA binding, B) Meiosis/DNA Repair/Homologous Recombination and C) Cell Division/Meiosis, compared to D) Housekeeping Genes. The number of damaged genes from the target gene list in the pathways of interest (red arrow) is compared to the distribution of damaged genes in random gene lists of equal number to the lists of interest (gray bars), burden-matched control genes (pink arrows) and housekeeping genes (green arrows). The burden-matched genes and housekeeping genes are not significantly enriched for any gene set. *p* values are controlled for the false discovery rate.

We examined the remaining candidate genes that had not yet been implicated in a woman with POI or in an animal model. We identified several deleterious variants in genes found in the implicated gene sets (Table 5)^70–77^. None of these gene variants was identified in the control groups that we assessed. Additional candidates were identified in these and other gene sets, although their pathogenicity was not as strong based on conservation, allele frequency or gene constraint (Supplementary Table 7).

**Table 5.**
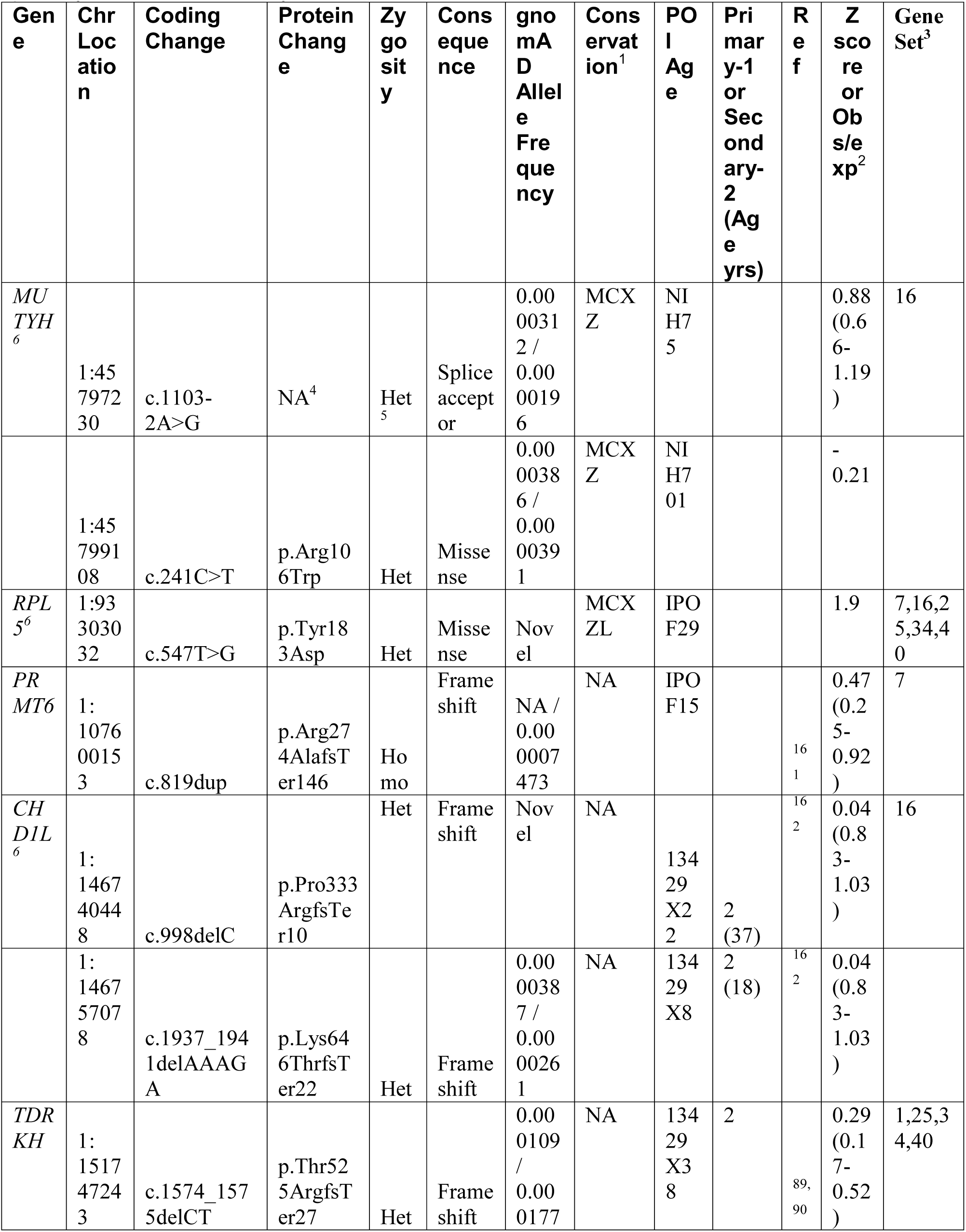
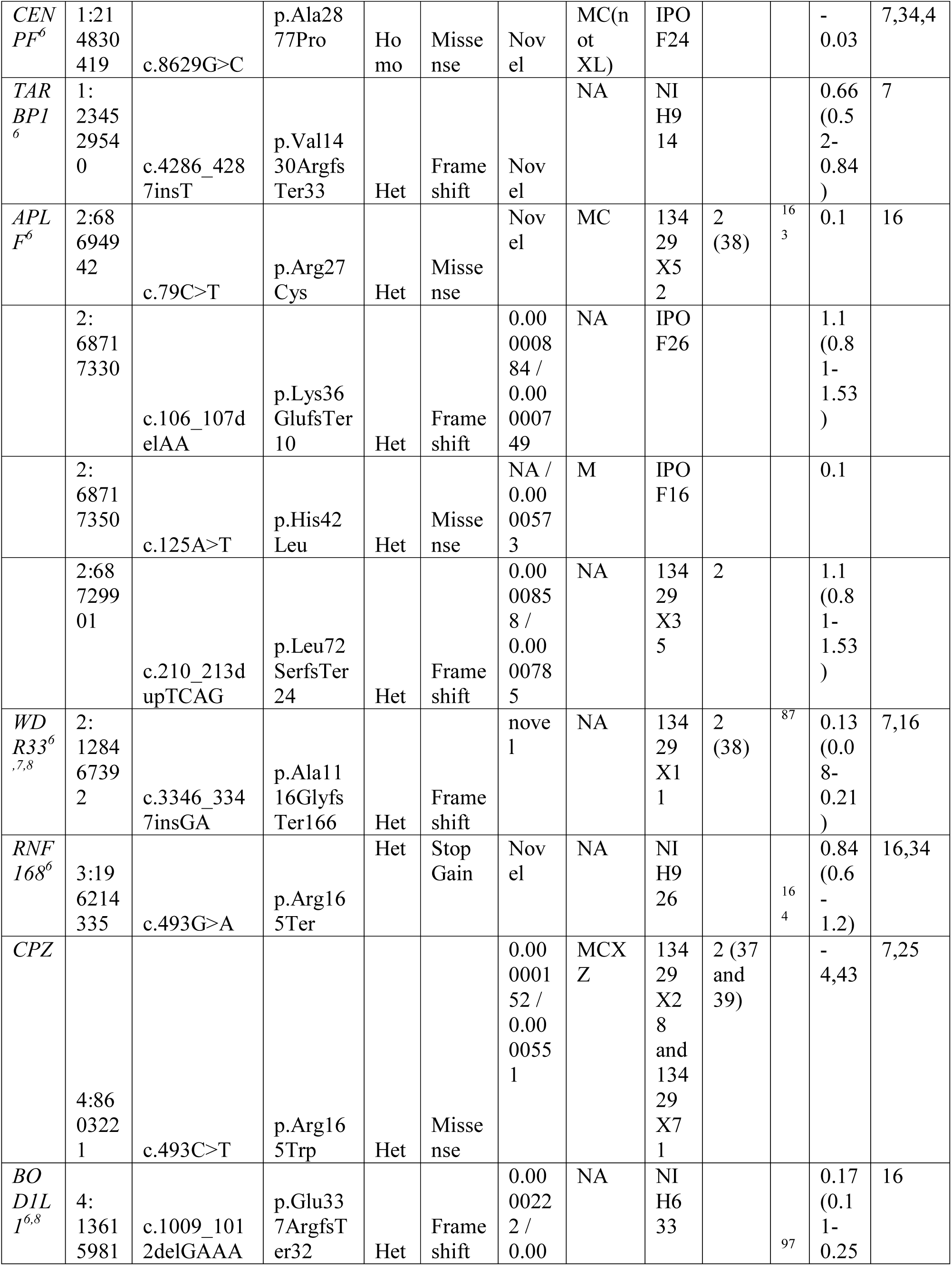
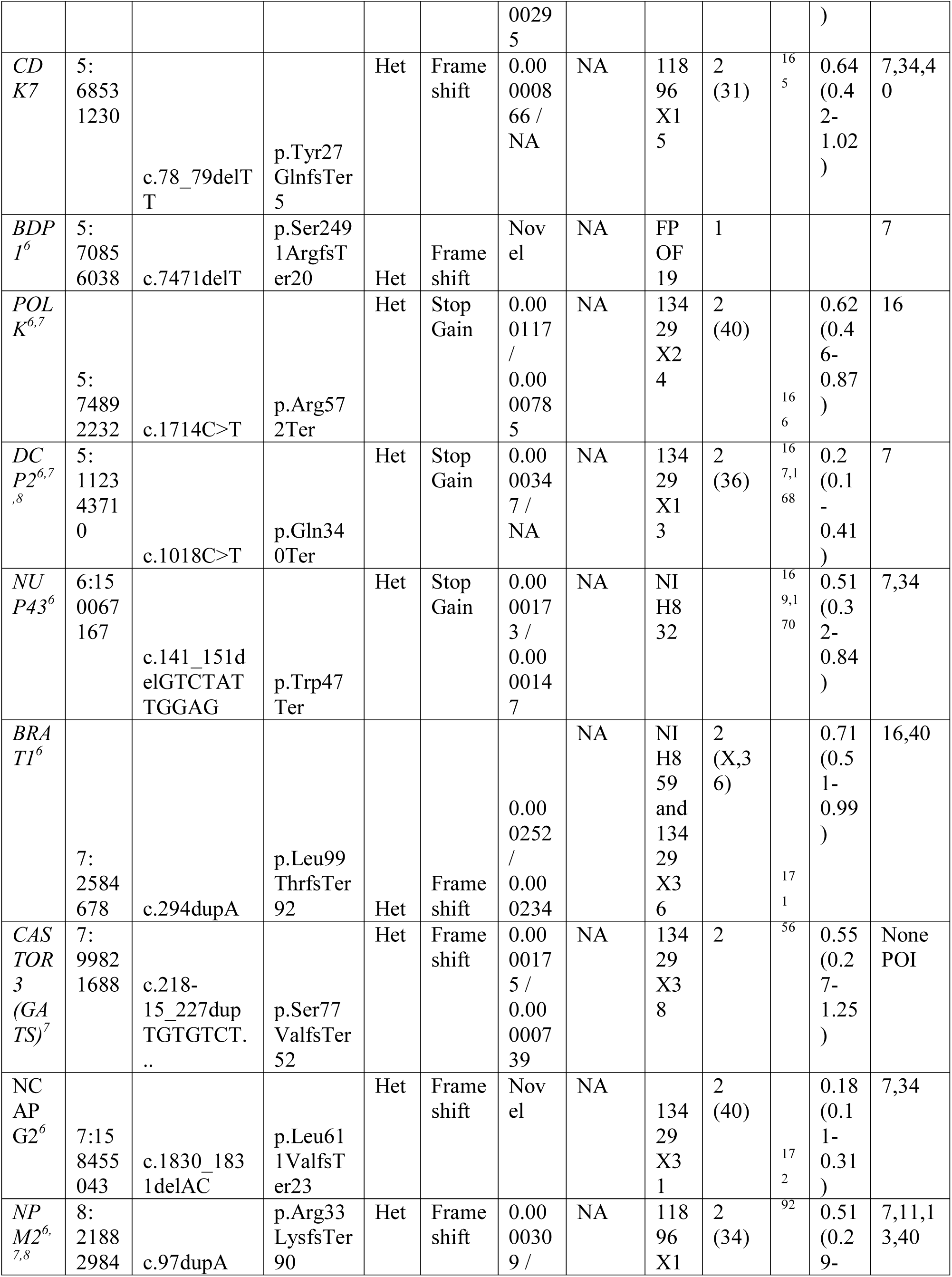
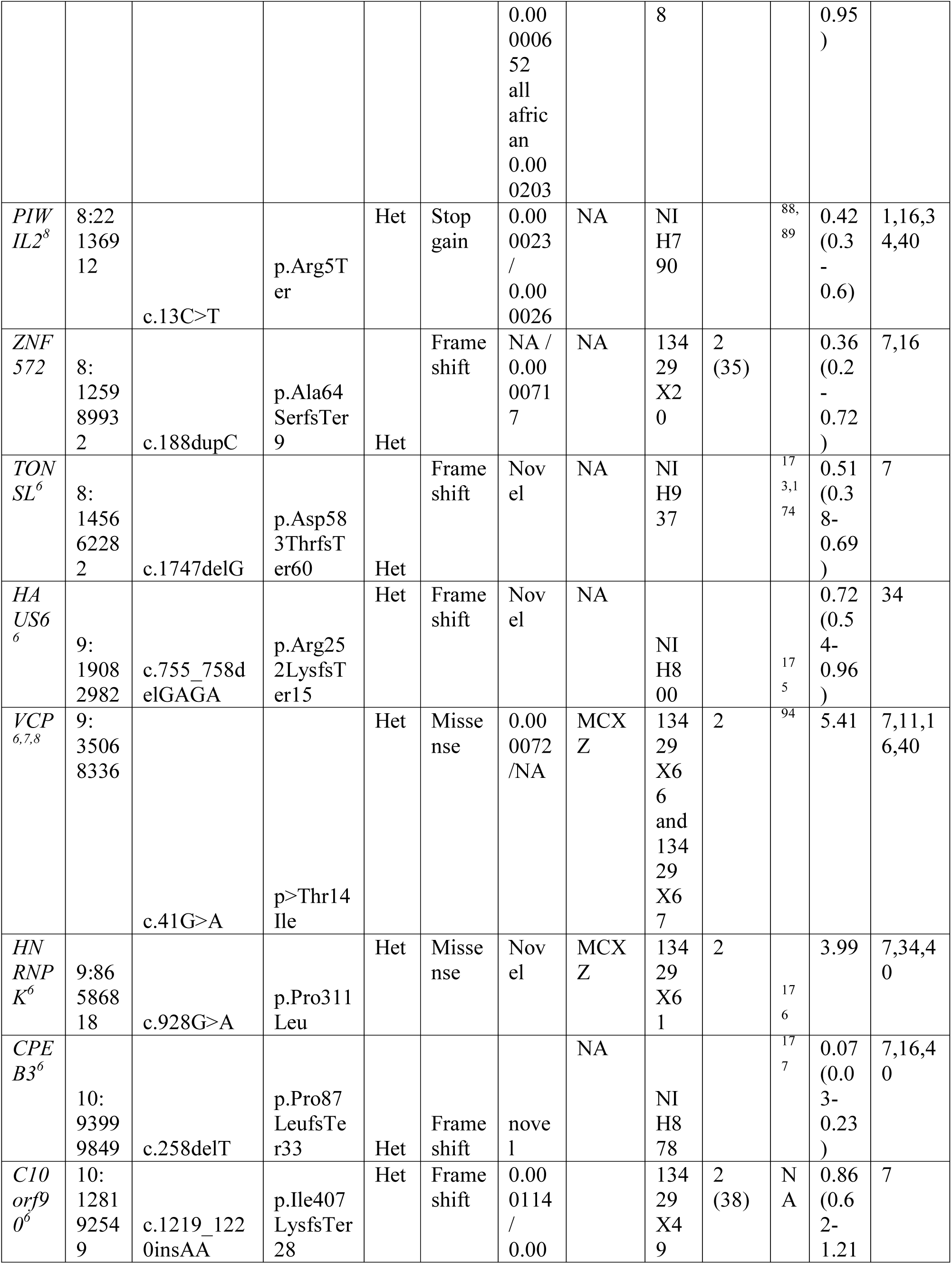
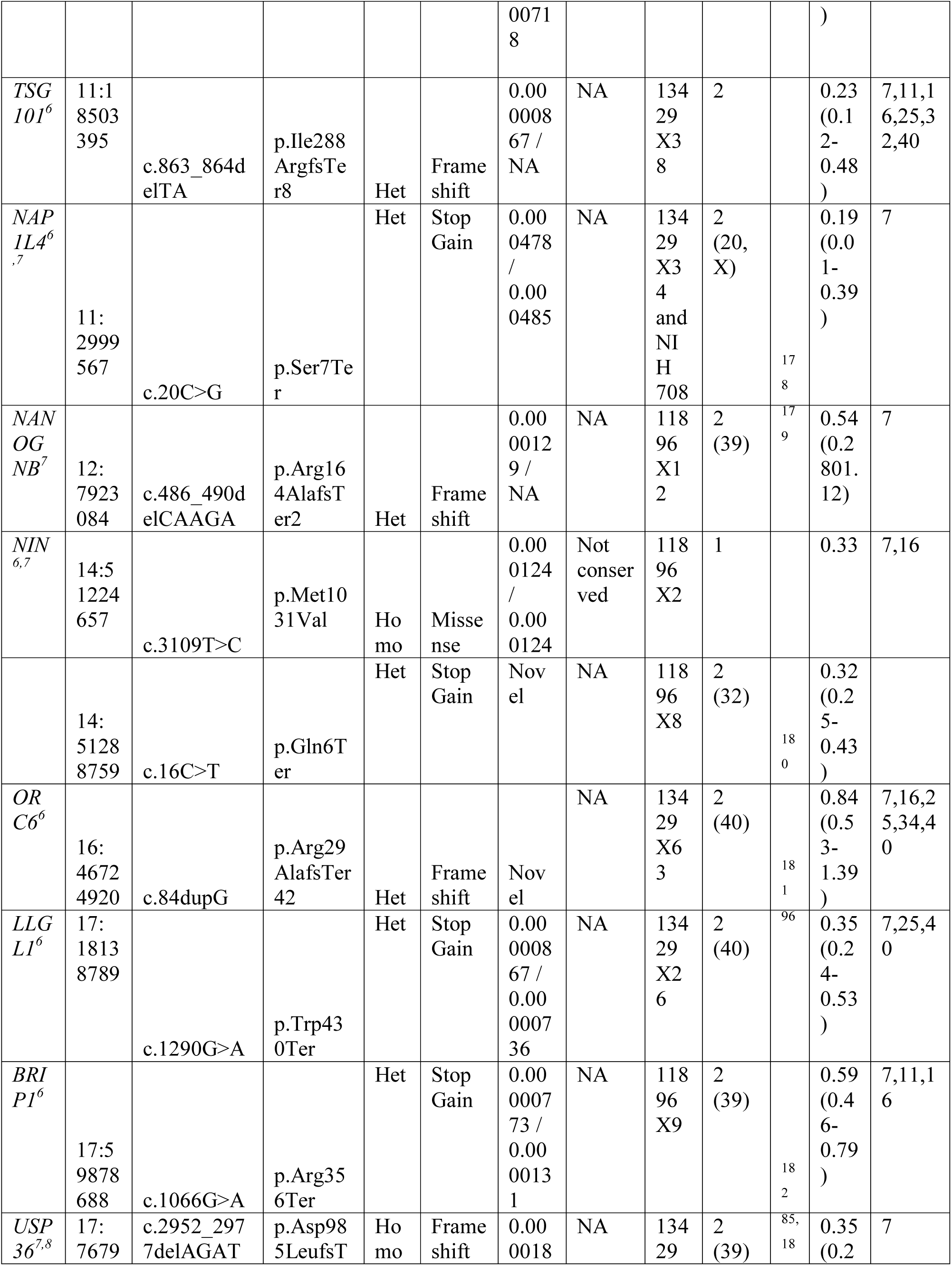
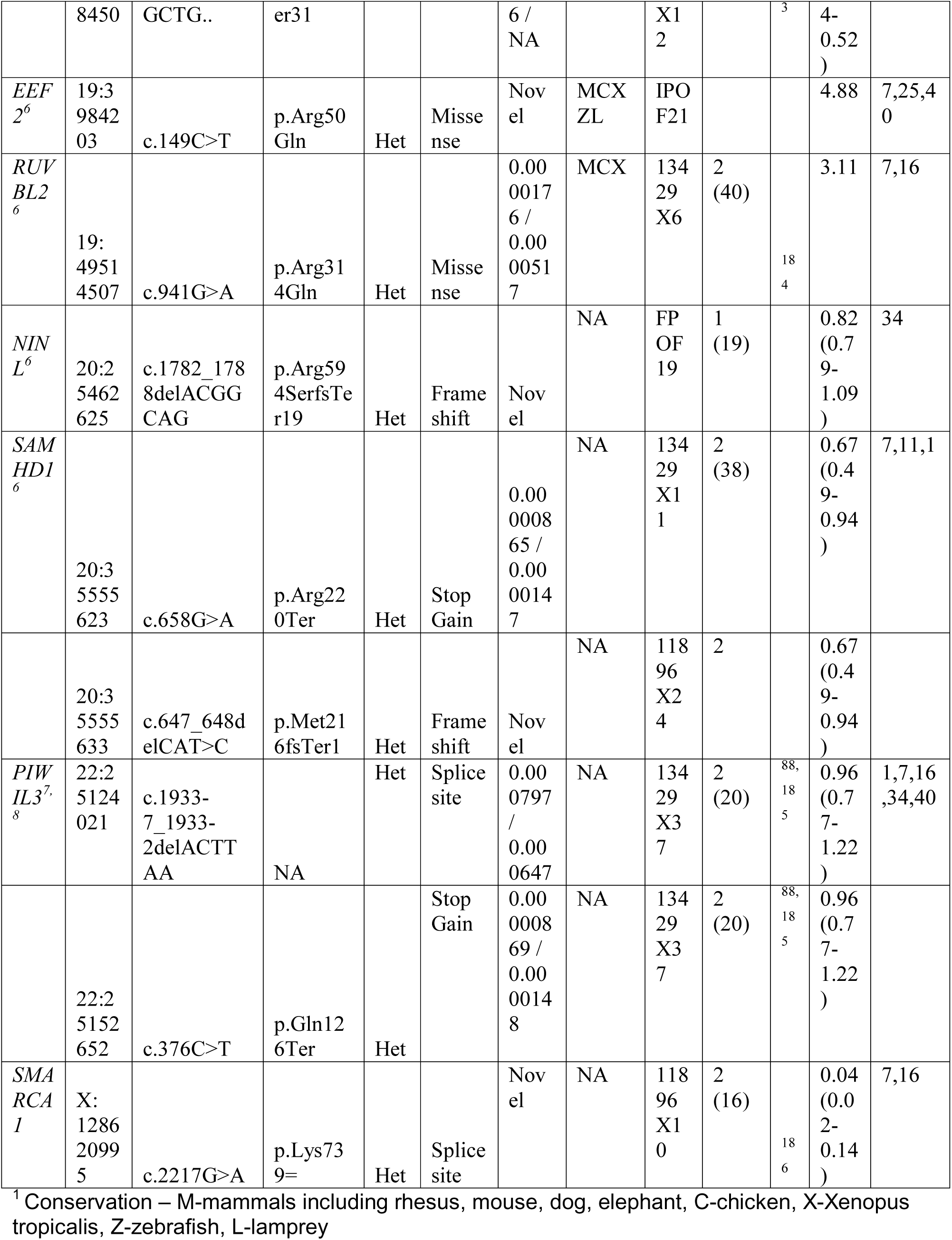
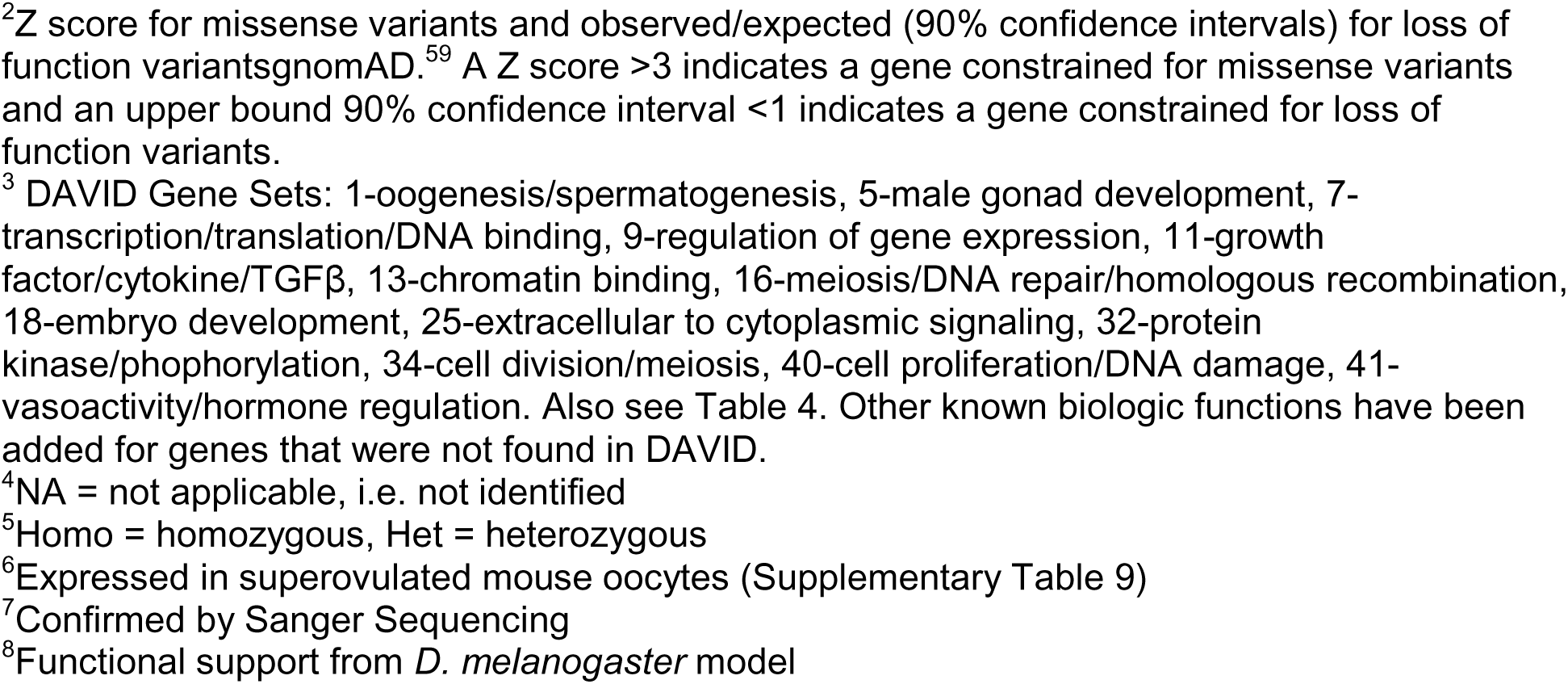
Variants in candidate genes and candidate pathways with no previous model for primary ovarian insufficiency.

### Functional Studies

The potential pathogenicity of the variants not previously identified is outlined in the Supplementary Data (Supplementary Information). For genes with variants not previously identified in POI or in animal models of POI or not previously examined in oocytes (Table 5), RTPCR in mouse oocytes that have resumed and/or completed meiosis I was performed to ensure that the candidate gene was expressed. Of the 24 genes tested, four were present but not highly expressed in the oocyte (Table 5 and Supplementary Tables 3 and 8).

*D. melanogaster* orthologues were identified for 20 of 35 candidate genes queried (Supplemental Table 9). Thirteen candidates could not be obtained based on availability or could not be tested based on lack of orthology or absence of ovarian expression. Two of the candidates had multiple weak orthologues and were not pursued (*POLK* and *ANKRD31*).

Five knockdowns (*USP36, VCP, WDR33, PIWIL3* and *NPM2*) were completely infertile with atrophic ovaries (Table 6 and Supplementary Figures 18 and 19). Two gene knockdowns demonstrated decreased hatchability and fertility, with abnormal (*LLGL1*) or normal ovaries (*BOD1L1*). Two gene knockdowns had variable or mild ovarian defects that were not statistically significant (*CDK7* and *BRIP1*). One gene knockdown was lethal (*RUVBL2*).

**Table 6.**
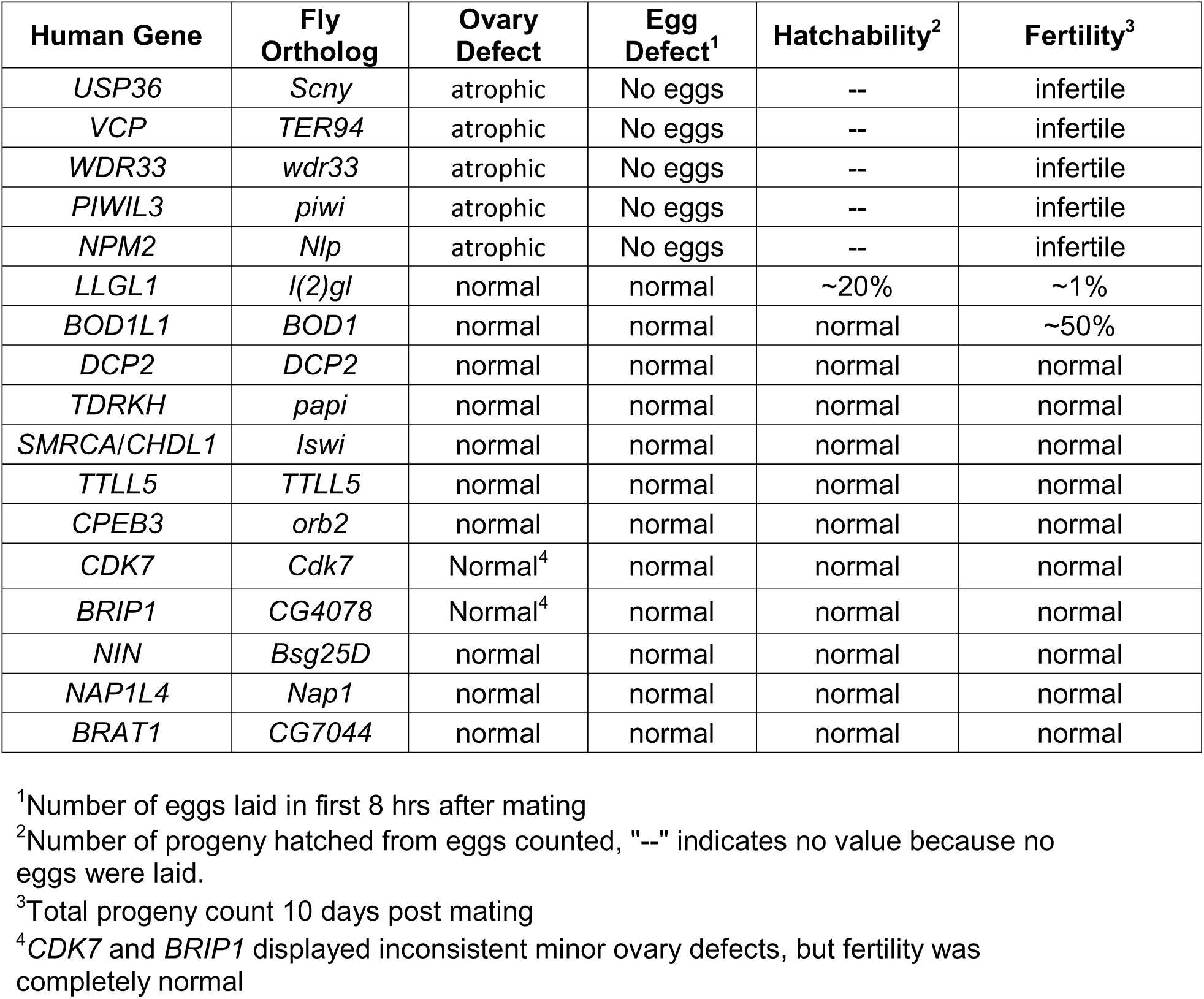
Ovary and fertility phenotypes in *D. melanogaster* RNAi knockdown in ovaries/germline.

## Discussion

We performed WES in 291 subjects with POI from three cohorts. Using two methods, a broad and unbiased discovery method and a more robust prioritization algorithm (GEM), we identified the most likely pathogenic variants in these women with POI. Our data suggest that the candidate genes for POI in individual women are highly heterogeneous. However, when the most likely candidate genes were categorized into functionally related groups, the genes aligned into 13 clusters that were enriched in cases compared to controls after correcting for multiple testing, gene size and pathogenic specificity for POI. New candidate genes were found in enhanced gene sets that included genes important for transcription/translation, DNA damage and repair, meiosis and cell division. Functional analysis in *D. melanogaster* supported a role in oocyte or ovary development for seven genes not previously associated with POI. Taken together, the data support a categorical approach to understanding the genetic architecture for POI.

After an initial broad search for damaging variants, we used an AI-based eCDSS tool, GEM, that employs variant impact (VAAST and VVP), patient phenotypes (Phevor), known Mendelian and pathogenic variants (OMIM, ClinVar) and ancestry to identify disease-causing genotypes^15,24,25,42,54^. Using GEM, we supplemented our data with previously analyzed WESs and have replicated genetic findings in 11 out of 12 subjects with primary amenorrhea, demonstrating the utility of the new software. The only gene variant that was not identified was in *MARF1*, which has not yet been associated with POI in OMIM. GEM identified additional homozygous or compound heterozygous mutations (*HFM1, DCAF17*) or heterozygous mutations (*NR5A1*) in previously identified genes^78–80^.

GEM also identified heterozygous deleterious variants in known genes, particularly in women with POI and secondary amenorrhea. GEM uncovered variants in 28 genes previously demonstrated to cause POI with recessive inheritance, out of 42 gene variants total (67%). GEM also identified 13 of 20 candidate variants in genes with evidence for ovarian insufficiency in an animal or other experimental species model (65%). The results are not surprising based on the use of HPO POI terms in the algorithm, which emphasizes phenotype in GEM^54^. Further tool development will encompass gene pathways for discovery.

With the exception of subjects with POI and primary amenorrhea, the majority of candidate variants identified in the current study are heterozygous, arguing for a dominant, semi-dominant, or complex inheritance pattern for POI that occurs later in the reproductive years. Genome-wide association studies (GWASs) of age at natural menopause support the concept that menopause has a complex inheritance pattern^68^. Further, the largest GWAS of early menopause, defined as menopause before the age of 45 years, replicated 4 common variants associated with age at natural menopause and demonstrated that menopause risk alleles have an additive contribution to age at menopause^21^. Recent data also suggest that common variants contribute to menopause occurring as early as 34 years^22^. However, the contribution of common variation explains only a small portion of the genetic risk for menopause under age 40 years^22^. The current study using WES was not able to assess common variants, but did demonstrate overlap between common GWA variants and rare, deleterious variants in the same candidate genes. For example, nonsynonymous variants in *BRCA1* are associated with earlier menopause by approximately 6 months^68^. In the current study, we identified a frameshift mutation, expected to result in early protein termination, possibly causative for POI. Additional genes with deleterious variants (*MSH6, CHD7)* and some with rare missense variants (*RAD54L, HELQ, POLG)* also overlap with candidate genes associated with age at natural menopause (Table 2). The apparent overlap of common variants associated with menopause age and deleterious variants in the same candidate genes is consistent with the hypothesis that mutations in these genes play a causative role in POI.

Further support for the causative role of heterozygous gene variants in POI comes from the reproductive history of the mothers of girls with primary amenorrhea. A heterozygous *MND1* gene mutation in a mother resulted in POI at age 35 years^45^. Similarly, a heterozygous mutation in *MCM8* caused POI in a mother at age 29 years^18^. In both families, the daughters with homozygous mutations presented with primary amenorrhea. With the exception of a few reports, age at menopause is rarely mentioned or may not yet have occurred for mothers of girls with POI. However, age at menopause is heritable supporting the segregation of ovarian damaging genes with an effect on age at menopause through the mother^3^. It is also not surprising that heterozygous variants that relatively decrease fertility would be removed from the population through decreased progeny^81^, and might therefore be inherited from the father since reproductive lifespan is not limited in men. Taken together, these cases also support the hypothesis that heterozygous mutations can result in earlier age at menopause.

Although the number of subjects in the current study is not sufficient to replicate the genes individually, we were able to demonstrate significantly enriched gene clusters controlled for multiple testing. Previous work in autism and congenital heart disease has used a similar category-wide association study approach^27,28^. Our approach was unbiased; first examining the most deleterious variants in women with POI to identify known genes and candidates with previous functional models, and subsequently determining whether additional genes were found in the clustered gene sets. Interestingly, a MAGENTA analysis of age at natural menopause variants identified similar enhanced categories for candidate genes inferred from genome-wide associated variants^68^. In addition, known genes causing male azoospermia were enriched in comparable pathways^82^. Taken together, a category enhanced approach identifies consistent gene sets across reproductive studies. Genes falling into gene sets including oogenesis, spermatogenesis, meiosis, DNA damage and repair, transcription and translation, chromatin binding, regulation of gene expression, growth factors, embryo development, cell division, extracellular to cytoplasmic signaling, protein kinase phophorylation, and vasoactivity and hormone regulation were enriched compared to controls in our unbiased candidate gene search for damaging mutations across the genome (Figure 1)^83^. New candidate genes were identified within these gene sets demonstrating that the category approach provides a mechanism for new candidate gene discovery.

Our *D. melanogaster* knockdown model affords a mechanism to determine an oocyte and ovarian phenotype at scale for genes in enhanced pathways. The genes and developmental processes involved in oogenesis in *D. melanogaster* overlap with those in the mouse^84^. We chose quantifiable fertility assays including egg laying rates, hatchability and ovarian morphology^62^. The use of RNAi technology also presumes that the gene is not fully deleted and serves as an excellent model for heterozygous gene variants. Using our *D. melanogaster* model, we identified five genes that are critical for ovarian or oocyte development and that fall into the enriched pathways we defined: transcription/translation, meiosis, DNA repair and DNA damage. RNAi knockdown resulted in atrophic ovaries with no eggs or progeny (Table 6 and Supplementary Figures 18 and 19).

*USP36* is a deubiquitinase demonstrated to promote RNA polymerase I stability for the ribosomal RNA processing and translation^85^. Previous studies found that the scny *D. melanogaster* homologue also acts as a histone H2B ubiquitin protease^86^. The atrophic ovary in the knockdown shows that the ribosomal RNA translation and/or the chromatin modification function may affect oocyte or ovarian development in addition to its role in embryogenesis^85^.

*WDR33* plays a role as one of 4 proteins that recognize the polyadenylation signal in the 3’-end processing of mRNA precursors^87^. The gene is highly expressed in testes and we have now demonstrated that it is also highly expressed in mouse oocytes (Supplementary Table 8). RNAi knockdown results in an atrophic ovary. Thus, *WDR33* may also play critical role in ovarian or oocyte development.

*PIWIL3* is a P-element induced wimpy testis protein short RNA found in human, nonhuman primate and bovine oocytes. It is specifically expressed in maturing human oocytes during oogenesis^88^ and in bovine oocytes from the GV stage onward^89^. It is critical for germline integrity from DNA transposable element activity^90^. The affected subject carries two *PIWIL3* variants; a frameshift mutation and a stop gain mutation that both remove the PIWI domain from the protein^90^. We also identified a stop gain mutation in *PIWIL2,* a family member that is expressed in fetal human germ cells^89^. Although knockouts of the mouse *PIWIL2* homologue *Mili* were described as fertile, there were no details provided across the reproductive lifespan^91^. These data demonstrate the importance of the *PIWIL* genes in the ovary in addition to the testes.

*NPM2* is found in oocytes before germinal vesicle breakdown^92^. The *Npm2* knockout females are infertile, with normal sized pronuclei that lack nucleoli^92,93^. Although previous studies suggest that infertility is caused by failure of zygote development, our data suggest that *NPM2* is critical for oocyte and ovary development.

*VCP,* or valosin-containing protein, is an ATPase associated with a variety of activities^94^. It is expressed in GV oocytes and preimplantation embryos in the mouse and controls germinal vesicle breakdown. *Vcp* knockout mice demonstrate no homozygotes because they have a defect in early embryonic development. Our model demonstrates atrophic ovaries. A missense variant in a highly conserved threonine in the N terminal was found in two sisters and a mother with POI (Supplementary Figure 2E). The N terminal is the portion that interacts with other proteins. Therefore, VCP should be considered a new candidate for POI.

In contrast to genes described above, there were only subtle phenotypes identified using our model in genes associated with DNA damage and repair pathways. Two genes in the pathway, *LLGL1* and *BOD1L1,* were highly expressed in the oocyte and demonstrated decreased hatchability and decreased fertility. The *LLGL1* cytoskeletal network is involved in maintaining cell polarity and epithelial integrity^95^. Mutations including the gene region on chr 17 cause Smith Magenis syndrome, a disorder of developmental delay, behavioral abnormalities, sleep disturbance and abdominal obesity. An indel upstream of *LLGL1* in Shaanbei White Cashmere goats is associated with change in litter size^96^. *BOD1L1* stabilizes *RAD51* at the site of DNA replication forks^97^. The frameshift variant in our subject would remove all ATM phosphorylation sites, along with the majority of the protein. Other genes in the DNA damage and repair pathway with no previous functional models or human mutations had no phenotype in our *D. melanogaster* model and more sensitive functional models may be needed. Nevertheless, a number of previously well validated variants involved in the homologous recombination steps in meiosis were discovered in our cohort (Figure 2)^83^. Many of these gene mutations may result in meiotic failure and oocyte loss. Given the large number of gene mutations falling into the DNA damage and repair pathway, intervening to rescue meiosis for development of normal gametes may be a treatment opportunity in POI.

**Figure 2.**
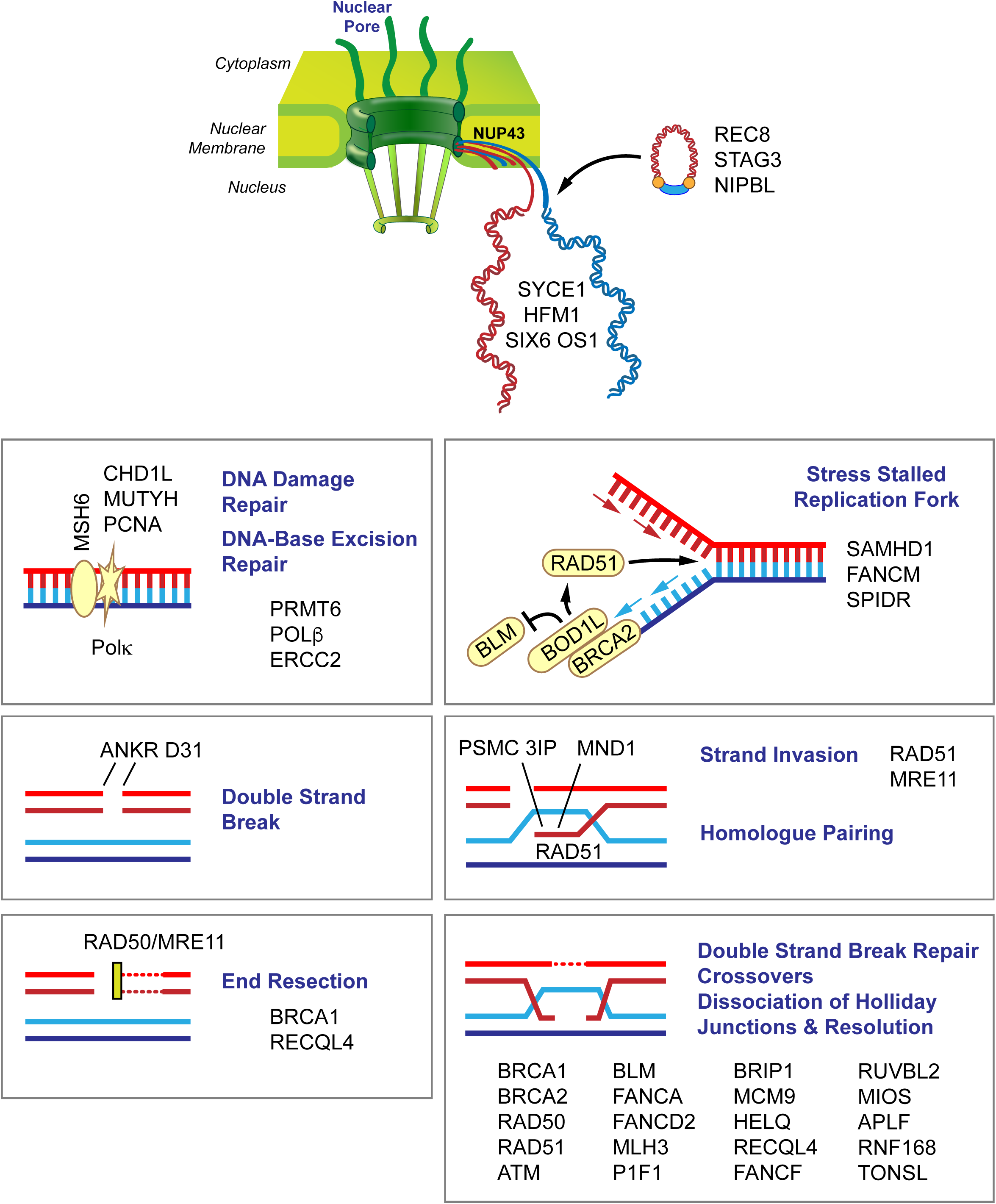
Candidate genes in women with POI. Variants in a number of genes involved in chromosome pairing and DNA damage and repair are involved in meiosis. The figure depicts candidate genes that are involved in chromosome movement, double strand breaks, end resection, double strand break repair, crossovers and dissociation and resolution of Holliday junctions. Members of the nuclear pore complex (NUP43) play a role in chromosome movement and organization. After DNA replication (*ORC6*), the synaptonemal complex pairs homologous chromosomes (*PSMC3IP*) loaded with condensin and cohesion complex proteins (*STAG3, REC8, NIPBL*) and connects the synaptonemal complex to DNA repair proteins (*SYCE1*). During recombination, double strand breaks form (*ATM*, *ANKRD3, PIF1*), ends are resected (*BRCA1, SAMHD1, BOD1L1*), and crossovers occur (*HFM1*) through strand invasion (*PSMC3IP, MND1, RAD51*). Subsequently, DNA double strand break repair (*CHD1L, POLG, POLK, MSH6, PCNA, NUPR1, APLF, NBN, RAD50, RUVBL2, MRE11*), DNA repair (*CDK7, MLH3, PRMT6, HELQ, TONSL*), strand annealing (*RECQL4*) and repair via homologous recombination (*BRCA2, BRIP1, FANCD2, HELQ, FANCM, FANCF, BLM, MCM9, USP36*) take place. Kinetochore/chromosome assembly, orientation and segregation (*HAUS6*, *CENPF, NUP43, NCAPG2, LLGL1, NINL, ATRX*) follow recombination.

An association between autoimmune oophoritis, with POI as the end-stage, has been demonstrated only with adrenal autoimmunity^98^. We identified a novel *PTPN22* variant, a gene associated with adrenal insufficiency, and in*TARBP1,* a gene associated with autoimmune syndromes^99,100^. A final subject carried a variant in IL1B, which has been associated with ovarian inflammation. Further delineation of the associated autoimmune risk genes and diseases will clarify the relationship between autoimmunity, genetics and POI.

Our study is limited by whole exome sequencing. We were not able to evaluate common variation, some promoter regions and could not evaluate copy number variants. We did not have trios for the majority of subjects and did not recruit family members to clarify segregation or *de novo* mutations. Future studies will also be needed to more carefully analyze the mitochondrial genome.

The current cohort forms one of the largest WES datasets analyzed for POI. We used an unbiased approach and a new AI-based algorithm to identify the most likely pathogenic variants. We also demonstrated new genes important for oogenesis and ovarian development using a model *D. melanogaster* system. Our more global approach contrasts to previous studies that examined individual consanguineous families and/or were restricted to candidate gene lists. Collectively, our results identify not only disease-causing variants, but also gene categories involved in POI. These results should prove useful for precision medicine efforts aimed at early identification of gene variants increasing a woman’s likelihood to experience infertility or a shortened reproductive lifespan. The early identification of women at risk for POI may enable fertility preserving measures. More broadly, better understanding of the genetic architecture of POI might also aid in identifying additional comorbid risks in a subset of the subjects.

## Data Availability

The data from NIH is available in dbGAP (Project #11971).

https://www.ncbi.nlm.nih.gov/gap/

## Acknowledgements

The work in this publication was supported by the Center for Genomic Medicine and its Functional Analysis Service. The work was also supported by R56HD090159 and R01HD099487 (CKW) and R35GM124780 (CYC and EC). The content is solely the responsibility of the authors and does not necessarily represent the official views of the National Institutes of Health.

---

Supplementary Table 1. Annotation clusters identified using the Database for Annotation, Visualization and Integrated Discovery (DAVID). Data were organized into 47 clusters with enrichment scores of >2.

Supplementary Table 2. Housekeeping genes. Genes that are constantly and uniformly expressed over many developmental and adult time points in 16 tissue types were chosen as housekeeping genes to examine enrichment.

Supplementary Table 3. PCR primers used to analyze gene expression in super ovulated mouse oocytes using RTPCR.

Supplementary Table 4. Samples removed by Peddy for very low heterozygosity and low coverage.

Supplementary Table 5. Related subjects identified by Peddy.

Supplementary Table 6. Ancestry identified by PCA plot and projection onto 1000 genomes data.

Supplementary Table 7. GEM results for all subjects. The GEM results for all genes with a GEM score greater than 0 are presented.

Supplementary Table 8. Oocyte expression. RTPCR was performed in superovulated mouse oocytes for gene targets with no previous functional studies.

Supplementary Table 9. Genes chosen for RNAi knockdown in a *D. melanogaster* model with Bloomington *Drosophila* Stock Center Number (BDSC#).

Supplementary Figure 1. Quality control metrics for 283 POI cases.

Box whisker plots of the alignment statistics and vcf statistics for 283 cram and vcf files that passed QC metrics: A) Total number of reads per sample, B) Percentage of Aligned reads, C) Percentage of duplicate reads, D) Mean coverage per sample, E) Median coverage per sample, F) Percentage of Coverage over 20 bases, G) Number of SNPs per sample extracted from the Bcftools statistics, H) Number of Indels found per sample, and I) Average Depth per sample.

Supplementary Figure 2. Peddy analysis of 283 samples from the final VCF files of women with POI. A) The predicted sex was female for all cases. B) The proportion of heterozygous calls ranged from 0.12 to 0.18 at a median depth of 30 to 65. C) PCA projection of the 283 cases onto ancestry of 1000 Genomes data. The majority of subjects were of European ancestry as expected. D) Coefficient of relatedness between two samples plotted by sampling 25K sites in the genome and comparing the relatedness reported in the ped file to the relatedness inferred from the genotypes. Thus, five sib pairs were confirmed, along with grandparent-parent and parent-child relationships. E) Five pedigrees of relationships confirmed by Peddy and investigators.

Supplementary Figure 3-16. Enriched pathways in the POI data set as determined by the permutation tests in cases (upper left panels) and controls (lower left panels) and compared to data from the same pathways for the root phenotypic abnormality (upper right and lower right panels). The number of damaged genes from the pathways of interest (red arrow) is compared to the distribution of damaged genes in random gene lists of equal number to the lists of interest (gray bars), burden-matched control genes (pink arrows), and housekeeping genes (green arrows). The burden-matched genes and 16A) housekeeping genes are not significantly enriched for any gene set.

Supplementary Figure 17. PA-1 cells were transfected using PolyJet transfection reagent (SignaGen Laboratories, Rockville, MD) with WT *eIF4ENIF1* or *eIF4ENIF1* containing the c.603T>G variant created using the QuikChange II Site-Directed Mutagenesis kit (Agilent Technologies, Santa Clara, CA) into a pcDNA3.1(-) expression vector (Invitrogen, Carlsbad, CA) using the NEBuilder HiFi DNA Assembly Cloning Kit (New England Biolabs, Ipswich, MA). Stable cell lines were generated by selection of colonies resistant to 750 μg/ml G418 (Life Technologies, Carlsbad, CA). Cells were seeded at 3 × 10^4^ cells/well in an 8-well chamber slide. After 48 hours, cells were fixed with ice-cold 100% methanol for 5 minutes at room temperature (RT), followed by washing with PBST comprising 0.1% Tween-20 in 1× PBS (Fisher Scientific, Waltham, MA). Cells were blocked with 1% BSA and 22.52 mg/ml glycine (Fisher Scientific, Waltham, MA) in PBST for 30 minutes at RT and then incubated with an N-terminal antibody (Novus Biologicals, Centennial, CO) diluted in 1% BSA in PBST at 4℃ overnight. After another wash with PBST, cells were labeled with an anti-rabbit Alexa Fluor 594 (Invitrogen, Carlsbad, CA) for 1 hour at RT, washed with PBST as before, counterstained with DAPI (Southern Biotech, Birmingham, AL), and mounted with glycerol mounting medium with DABCO (Electron Microscopy Sciences, Hatfield, PA). The Nikon fluorescent microscope was used for image acquisition. The c.603T>G variant, p.S201R, is located in the nuclear import signal of *eIF4ENIF1*. The top panels show the N terminal eIF4ENIF1 images with DAPI staining of the nucleus, while the bottom panels show the N terminal eIF4ENIF1 images. Compared to the A) wild type *eIF4ENIF1*, the B) S201R variant transfected cells demonstrated disorganized localization of *eIF4ENIF1* with increased intranuclear protein.

Supplementary Figure 18. *Drosophila melanogaster* phenotypes.

Hatchability and total fertility values are plotted for all genes tested by RNAi in *Drosophila*. P values for genes with significantly different phenotypic values are highlighted in red. N= 8-10 for all measurements. C= control; KD= RNAi knockdown

Supplementary Figure 19. *Drosophila melanogaster* ovarian phenotype.

Representative images of ovaries from RNAi knockdowns that produced atrophic ovaries and a control. All other RNAi knockdowns that produced normal ovaries appear identical to the control and are not shown.

## Notes

### Competing Interest Statement

Disclosure: MY is a stock holder or has received stock option awards from Fabric Genomics Inc. BM and MY have received consulting fees from Fabric Genomics Inc. ARC is a consultant for Ferring and a scientific advisory board member for INVObioscience and Celmatix.

### Funding Statement

The work was supported by R56HD090159 and R01HD099487 (CKW) and R35GM124780 (CYC and EC) from the NIH.

### Author Declarations

University of Utah, Washington University, University of Pittsburgh and the Sorbonne Universite IRB.

