## Supplemental Data for "Causal and Candidate Gene Variants in a Large Cohort of Women with Primary Ovarian Insufficiency"

Supplementary Information

Meiosis

1. Nucleoporin-Related Genes

*NUP43* is a component of the human *NUP107* nuclear pore complex^1,2^. A homozygous missense mutation in NUP107 was demonstrated to disrupt protein structure and cause POI, with no effect on male fertility or other cellular processes^3^. The nuclear envelope breaks down during mitosis and meiosis and is reformed, largely driven by the *NUP107* complex. *NUP43* appears to be essential for the nucleoporin complex formation. The *NUP43* gene is moderately expressed in oocytes (Supplementary Table 9). The deletion mutation identified would result in loss of six of the seven WD40 repeats^1^. Of note, *NUP43* interaction modeling suggests that it sits in the *NUP107* complex and binds to *SEH1*^2^. The deletion mutation identified would abolish *SEH1* binding. *SEH1* mutations have been demonstrated to result in defective oogenesis in the Drosophila^4^. Therefore, a mechanism is plausible for defective oogenesis through shortened *NUP43*.

2. *HFM1*

Initially described in a Han Chinese woman with compound heterozygous mutations in *HFM1*^5^, mouse models demonstrate that *HFM1* is required to form normal numbers of crossovers during meiosis and to complete synapsis^6^. One subject in the current study carried a homozygous mutation, p.Ile709Asn, predicted to be deleterious and found in the helicase C-terminal. One additional subject, who was diagnosed with Noonan syndrome and carried a *PTPN11* mutation, also carried two *HFM1* variants, one predicted to be deleterious (Table 2) and one benign (Supplementary Table 8). As female patients with Noonan syndrome do not typically suffer from POI, it is possible that one or both *HFM1* mutations are causing the phenotype in this patient.

3. *SYCE1*

We identified a frameshift mutation in *SYCE1*. One previous study demonstrated a microdeletion resulting in haploinsufficiency of *SYCE1* in a woman with POI at age 21 yrs^7^. Therefore, it is possible that haploinsufficiency is causal for POI. *SYCE1* disrupts repair of DNA double strand breaks during meiosis^8^. The *SYCE1* protein connects the synaptonemal complex to DNA repair proteins^8^. The frameshift mutation removes the last 3 exons in *SYCE1* and may disrupt ability to complex with these DNA repair proteins.

4. *NIPBL*

*NIPBL* mutations cause Cornelia de Lange syndrome. The subjects with POI do not have Cornelia de Lange syndrome but carry missense variants that are novel and in highly conserved amino acids. They are not found in critical protein domains, although the Asn533Tyr is found in exon 10, which has the greatest number of missense variants and the His1128Gln variant is located immediately adjacent to the nuclear localization signal^9^. There is a spectrum of phenotypic abnormalities in Cornelia de Lange syndrome and irregular menses appears to be a common feature, although the cause is not well defined. The NIPBL cohesin regulatory protein is critical for meiosis and studies in *Drosophila* demonstrate that oocytes lacking the *Drosophila* homologue *Nipped-B* do not progress beyond zygotene and are sterile^10^. Therefore, it is possible that mutations in *NIPBL* result in POI. It is interesting that the variants in the current study have not been described in the full syndrome and appear to be found in regions that are not considered functional domains, perhaps indicating less severe protein dysfunction.

5. *STAG3*

One subject carried a stop gain mutation in *STAG3,* which forms a cohesin ring that surrounds sister chromatids during meiosis^11^. Previous studies have not described age at menopause in the mothers of women with *STAG3* mutations and POI. Another subject carried a frameshift variant in the antisense transcript pseudogene *GATS* (*CASTOR3*). It is not clear if that antisense transcript plays a role in the oocyte or in meiosis. A homologue is not found in mouse for examination.

6. *PSMC3IP*

One subject carried a splice acceptor variant in *PSMC3IP*. We and others demonstrated that homozygous mutations in *PSMC3IP* cause POI^12,13^. Although the splice site is rare, it sits at a NAGNAG site, which is often used as an alternate splice site and removes one codon in a frame preserving manner^14^. The splice site is located on the final exon, which interacts with RAD51/DMC1^15^. In the heterozygous state, it is not yet clear if it affects age at menopause, but fertility appears normal in carrier mothers during the reproductive years^13^.

7. *MND1*

One subject demonstrated a frameshift in *MND1*, a gene recently identified as a cause of POI with a homozygous deletion of exon 6 in a consanguineous family^16^. The frameshift mutation deletes the final three exons 6 through 8 in *MND1*. *MND1* forms a complex with *PSMC3IP* for proper single to double strand DNA pairing^15^.

8. *ATM*

Mice with *ATM* deletions have oocyte loss through failure to repair DNA double strand breaks^17^. One subject had a frameshift mutation. The rest carried *ATM* missense variants in conserved amino acids. Although the missense variants were of uncertain significance, there were no ATM damaging variants found in any controls^18^.

9. *ANKRD31*

*ANKRD31* acts as a scaffold regulating double strand break formation. *ANKRD31* deletions in a mouse model result in POI^19^. The frameshift mutation would be expected to delete the last 5 exons of the protein.

10. *C11orf80*

*C11orf80,* also known as *TopoVIbl* is expressed in the testis and is critical for the formation of double strand breaks^20^. Male mice knockouts had smaller testes. However, there was no discussion of the effect on ovarian function. Expression was not high in the oocyte (Supplementary Table 9). Therefore, additional studies are needed and the frameshift mutation identified was not reported in the final results.

11. *PIF1*

*PIF1* mutations in *Drosophila* result in failure of DNA replication after double strand breaks, with failed embryogenesis and hatching^21^. Homozygous missense variants have been described a woman with recurrent pregnancy loss and failed embryogenesis^22^. The subject in the current study has a frameshift mutation, which will need additional functional studies.

12. *CHD1L*

*CHD1L* localizes to areas of DNA damage^23^. It sustains activity through recruitment of DNA repair proteins such as *PARP1*. The frameshift mutations remove the Macro domain that binds PARP and one also removes the C terminal helicase domain^23^. The gene is expressed in mouse oocytes (Supplementary Table 9). However, we were not able to confirm a phenotype in our *D. melanogaster model* (Table 6).

13. *PCNA*

*PCNA* encircles damaged double strand DNA and recruits necessary polymerases^24,25^. It is highly expressed in primordial follicles and regulates physiological apoptosis^26^.

14. *FANC* and *BRCA* Genes

The genes coding for Fanconi anemia are associated with ovarian insufficiency in association with hematologic malignancies^27^. In general, severe mutations in the Fanconi anemia genes predispose to cancer risk in a heterozygous state^28^. There is one report of familial POI with secondary amenorrhea associated with a homozygous stop gain mutation in *FANCM* at the C-terminal end^29^. In the current study we identified 2 subjects with heterozygous missense variants in *FANCM* and *FANCD2,* along with BRCA1 and BRCA2, discussed below. The *FANCM* missense variant changes alanine to valine at amino acid position 121, located in the ATP binding helicase domain, and the variant is predicted to affect protein function. One maternal carrier of a stop gain mutation had regular menses at age 47 years, therefore any effect on age at menopause for heterozygous *FANCM* variants may be small if carried alone^29^. The *FANCD2* frameshift results in premature termination of protein coding, before a critical ubiquitination site^30^. We also identified a stop mutation in *BRIP1 (FANCJ*), which has not previously been described as a causal gene for POI. A mutation in *FANCF* was also identified in one subject. The *FANC* genes (A-C, E-G and L) monoubiquitinate the *FANCD2* and *FANCI* heterodimer, which regulates the DNA repair/replication rescue activities of the third group of *FANC* proteins, which includes *BRCA1* and *BRCA2*.

Two subjects had deleterious mutations in *BRCA2*. In many previously identified families with homozygous mutations in POI genes, it has been difficult to determine whether the heterozygous mother has early menopause or POI. In one case, a *BRCA2* gene mutation carrier suffered from ovarian cancer and menopause age was not determined^3^. Studies have provided conflicting information regarding the reproductive potential in women with *BRCA2* mutations. Some suggest a longer reproductive lifespan in *BRCA* mutation carriers^31,32^, whereas others suggest earlier menopause, lower AMH levels and poor response to ovarian hyperstimulation, which support the concept of decreased follicle number^33-43^. A recent study demonstrates compound heterozygous mutations causing primary amenorrhea and ovarian insufficiency^12^. One study delineated the *BRCA2* mutations in women with low AMH levels, but only one mutation per person was found and it would be of interest to characterize the ovarian follicle complement based on specific mutations^42^. Both the frameshift and stop gain mutation found in our subjects are pathogenic and were not listed among subjects studied previously^42^. We also identified a missense variant (c.8585T>C; p.Leu2862Pro) in a highly conserved base pair that is predicted to be deleterious. One previous study did not find severe mutations in *BRCA1* or *2* in a group of less than 100 patients^44^. It has been suggested that the more severely *BRCA* compromising mutations may not have been studied if those women had early cancer and were not included in studies of menopause age. Since our termination mutations should severely affect protein function, it is possible that they are also more likely to cause severe ovarian dysfunction. We cannot rule out the possibility that a mutation in a second gene enhanced the severity of the effect on ovarian function in these women, but we have not been able to identify a distinct candidate in the current analysis.

15. *RNF168*

Recessive mutations in *RNF168* cause RIDDLE syndrome, a disorder of radiosensitivity, dysmorphic features, immunodeficiency, intellectual impairment^45^. A mouse knockout model also demonstrated decreased fertility in mouse heterozygotes at 8 weeks and male infertility at 12 months, although females were not studied. A stop gain mutation in one patient with POI was identified, and was documented to be pathogenic in ClinVar^18^. The gene is involved in DNA damage repair and is highly expressed in mouse oocytes (Supplementary Table 9).

16. *MSH6*

We identified a frameshift insertion in *MSH6* in a patient and her mother. The insertion has been classified as uncertain significance in ClinVar^18^. Expression is high in the ovary and a common exonic variant is associated with natural age at menopause^46,47^. Thus, it remains a candidate gene variant.

17. *MCM9*

*MCM9* mutations have previously been determined to cause POI^48^. In the current study, the results from two previously reported subjects were replicated, one with compound heterozygous mutations and one with a homozygous stop-gain mutation^49^. Heterozygous, rare variants were identified in 4 additional women. One of these variants was a stop-gain variant in the AAA+ core domain of the protein^50^. Previous data demonstrated heterozygous stop gain *MCM9* variants in women with POI and secondary amenorrhea^48^. One additional missense variant was found in a conserved amino acid in the AAA+ core domain, one in the DNA binding domain and one in the linker region.

18. *BLM*

*BLM* mutations cause a cancer syndrome with POI as a feature. One subject carried a novel heterozygous frameshift mutation. The identified *BLM* variant has overlap with other pathogenic mutations in the gene^51^. Previous reports of POI in Bloom Syndrome result from recessive inheritance^52^.

19. *BRAT1*

*BRAT1* codes for a *BRCA1*-associated and *ATM*-activator protein that stabilizes the complex for DNA repair^53^. Recessive mutations cause a neonatal seizure disorder resulting in early death. Heterozygous mutations may predispose to breast and ovarian cancer^53^. The gene is moderately expressed in oocytes (Supplementary Table 9). However, we were not able to confirm a phenotype in our *D. melanogaster model* (Table 6).

20. *SAMHD1*

Although *SAMHD1* has been studied in viral replication, there is evidence for its role in DNA replication at stalled replication forks^54^. When deleted, single strand DNA accumulates. The gene is highly expressed in mouse oocytes (Supplementary Table 9).

21. *HELQ*

The *HELQ* missense variant is located in a conserved base pair in the helicase region^55^. Ovarian expression is high^47^. A mouse *HELQ* deletion causes decreased follicle number in the ovaries and decreased litter number^55^. Heterozygotes have a milder infertility phenotype. Variants in *HELQ* are associated with age at natural menopause^56^.

22. *POLG*

Both dominant and recessive *POLG* mutations cause disease. In a woman with progressive muscle weakness and atrophy along with primary amenorrhea, a heterozygous missense mutation in *POLG* was implicated^57^.

23. *POLK*

*POLK* interacts with *PCNA* to replicate DNA after damage, maintaining DNA replicative ability while incorporating the mutation^24^. The stop gain mutation deletes the ubiquitin zinc finger binding domains at the C-terminal of the protein. The gene is expressed in mouse oocytes (Supplementary Table 9).

24. *NUPR1*

One subject carried a variant in *NUPR1*, a gene previously identified in a duplication region on a CGH array^58^. *NUPR1* is expressed at high levels in the ovary, but the mouse *NUPR1* model does not have a POI phenotype^59^. Rather, it appears to be important for LH receptor expression and deletion results in decreased corpora lutei. The stop gain variant is identified in the only region in which other stop gain variants are found in the gene.

25. *APLF*

*APLF* stabilizes factors involved in non-homologous end joining DNA repair after DNA damage^60^. The frameshift mutations identified would prevent *APLF* from binding *XRCC4* and other critical DNA repair factors. The missense variants at the N-terminal are found in the forkhead-associated domain, replacing highly conserved amino acids^60^. The gene is highly expressed in oocytes (Supplementary Table 9).

26. *NBN*

One subject had a splice site variant in *NBN*. The base pair is highly conserved^61^. Heterozygous deletion of *NBN* results in predisposition to cancers after exposure to ionizing radiation^62^, suggesting that a heterozygous genotype does carry risk. Although the heterozygous mice were determined to be fertile, there was no formal study of pup or litter number.

27. *RAD50*

*RAD50* is a component of the *RAD50/MRE11* complex that repairs DNA double strand breaks^63^. Both the identified frameshift and stop gain variants cause a shortened protein that would fail to form the expected complexes and DNA binding regions.

28. *MRE11*

Hypomorphic *Mre11* resulted in oocyte loss in mice at 12 weeks^64^. The variant we identified resulted in Arg572Ter, possibly resulting in failure of binding to *RAD50*^64^. We also identified a missense variant that replaces a highly conserved leucine.

29. *RUVBL2*

The RUVBL1/RUVBL2 complex associates with NEIL3 in the pathway of DNA repair, independent of the Fanconi anemia/*BRCA* pathway^65^. The missense variant is less common in females and is characterized as possibly damaging by Polyphen and deleterious by SIFT. Expression is very high in the oocyte (Supplementary Table 9). The siRNA knockdown resulted in lethality and its ovarian phenotype was not able to be tested.

30. *CDK7*

*CDK7* has a critical role for oocyte mRNA transcription through phosphorylating RNA polymerase II subunits and it acts as the catalytic subunit of transcription factor IIH in porcine oocytes^66^. Knockdown of *CDK7* results in arrest of GVBD in a majority of oocytes. Loss of *CDK7* prevents the premature activation of anaphase-promoting complex/cyclosome until the kinetochores are attached to the spindles in mice to control the meiosis I to meiosis II transition^67^. The frameshift mutation we identified might decrease the overall quantities of CDK7 in the oocyte and may interfere with complete GVBD. However, we did not identify a phenotype in our *D. melanogaster* model.

31. *MLH3*

We identified a frameshift and missense variant in *MLH3*. *Mlh3* deletion results in defective chromosome segregation in a mouse model^68^. Only 50% of *Mlh3* null mouse oocytes completed meiosis I after fertilization. The missense variant p.Ile239Ser was more common in men and was found probably damaging by Polyphen and deleterious by SIFT.

32. *PRMT6*

We identified a homozygous frameshift mutation in *PRMT6,* a single exon gene that is constrained for loss of function mutations. *PRMT6* demethylates histones to antagonize cofactor chromatin binding^69^. It also plays a role in DNA base excision repair^70^. Knockdown of *PRMT6* resulted in germ cell apoptosis in the male^71^. A deleted chromosome region containing *PRMT6* was associated with non-obstructive azoospermia in Han Chinese men^72^.

33. *TONSL*

*TONSL* recessive mutations cause SPONASTRIME dysplasia, a disorder of spondyloepimetaphyseal dysplasia^73^. *TONSL* plays a role in DNA replication and homologous recombination dependent repair. The frameshift mutation would be expected to remove the PB1 ubiquitin-like domain and the leucine-rich repeat region. There has not yet been a report of infertility in this disorder. There is low expression in mouse oocytes (Supplementary Table 9).

34. *RECQL4*

*RECQL4* recessive mutations cause growth retardation, radial defects, dermatologic changes and increase risk for osteosarcoma^74^. We did identify one subject with two missense mutations, but do not have parent DNA and cannot determine whether the variants are on separate alleles. Two additional subjects had a frameshift mutation and a conserved splice site mutation previously determined to be causal. Mutations cause a defect in sister-chromatid adhesion and chromosome segregation, along with sterility^75^.

35. *SMARCA1*

We identified a *SMARCA1* variant at a splice site in a highly conserved amino acid. *SMARCA1* is highly expressed in the oocyte and plays a role in ATP dependent chromatin remodeling and possibly DNA damage^76^. However, we were not able to confirm a phenotype in our *D. melanogaster model* (Table 6).

36. *HAUS6*

*HAUS6* is highly expressed in the testis and has a role in recruiting γtubulin and branching microtubules to the mitotic spindle^77^. The frameshift mutation removes part of the N terminal domain required for *HAUS6* to bind to the augmin complex in centrosome independent spindle assembly during meiosis^78^. The gene is highly expressed in mouse oocytes (Supplementary Table 9).

37. *CENPF*

Centromere protein F is located in the kinetochore where chromosomes and spindle microtubules interact for segregation in meiosis and mitosis^79^. One subject had a homozygous missense variant in a non-conserved amino acid that was predicted to be possibly damaging^80^. Recessive mutations cause Stromme syndrome, a ciliopathy disorder. The gene is expressed in mouse oocytes (Supplementary Table 9).

38. *NCAPG2*

*NCAPG2* forms one of the subunits of the condensin II complex, important for structural maintenance of chromosomes and condensation of chromosomes before cell division^81^. Mutations cause syndromic neurodevelopment abnormalities and can play a role in renal tubule defects. The gene is expressed in oocytes (Supplementary Table 9).

39. *NIN* and *NINL*

*NIN* and *NINL* are centrosomal proteins important for microtubule assembly, with mutations causing Seckel syndrome; primordial dwarfism, cognitive deficiencies and increased sensitivity to DNA toxic stress^82^. Although a *Drosophila* model did not identify fertility defects with *NIN* mutations (Table 6)^82^, expression was high in oocytes and additional models may be needed (Supplementary Table 9).

40. *ATRX*

Deletion of *ATRX* results in loss of premeiotic germ cells in males and females^83^. It is involved in chromatin remodeling and chromosome alignment in mitosis^84^. The gene is constrained for missense mutations^61^.

41. *CASP3*

*CASP3* is a pro-apoptotic signal in the pathway of cell demise. Deletion in a mouse model causes defective granulosa cell apoptosis but is not critical for germ cell apoptosis and does not cause follicle loss^85^. The subject carried a frameshift mutation in *CASP3*.

42. *SKP2*

*SKP2* regulates of p27, which acts to restrain the cell cycle. Deletion results in loss of oocytes in a mouse model and the heterozygotes also have decreased fertility^86^. *SKP2* is highly constrained^61^. Therefore, *SKP2* is an excellent candidate gene for POI.

43. *BCL2*

*BCL2* represses cell death through apoptosis. *Bcl2* deletion in a mouse model results in empty follicles with no oocyte^87^. Therefore, *BCL2* is another excellent candidate gene.

44. *MUTYH*

*MUTYH* is a base excision DNA repair enzyme. Recessive mutations cause adenomatous polyps and colorectal cancer. It also plays a role in other cancers and neurologic disease. The missense variant changing p.Arg106Trp is located in an amino acid that is highly conserved^88^. The splice site sits next to an exon that codes for the initial portion of the nudix hydrolase domain^88^. Both are determined likely pathogenic in ClinVar^18^. There is low expression in mouse oocytes (Supplementary Table 9).

45. *SIX6OS1*

*SIX6OS1,* or *C14orf39*, deletion in mice resulted in lower follicle number by day 6 postpartum and at 4 months^89^. The protein is essential for mouse fertility and interacts with SYCE1 at the N terminus^89^. The frameshift deletion results in deletion of the N terminal region.

46. *C10orf90*

*C10orf90*, also known as FATS, exhibits high expression in testis, brain and some in ovary, which we replicated in our mouse oocytes (Supplementary Table 9)^47^. The gene is a ubiquitin ligase that activates p53 in response to DNA damage^90^.

47. *ERCC2*

*ERCC2* is critical for nucleotide excision repair. Mutations cause Xeroderma Pigmentosa and mice with a missense mutation in *ERCC2* exhibit premature aging and infertility^91^. ERCC2 levels decline in aging rat oocytes, one of the DNA damage repair proteins found in oocytes^92^.

48. *ORC6*

*ORC6* codes for a mutation in the gene causing Meier Gorlin syndrome, a disorder of growth, microcephaly and patellar aplasia, with some evidence for menstrual cycle dysfunction, although it is poorly characterized^93^. The gene is expressed in the ovary (Supplementary Table 9). The frameshift mutation identified would terminate the protein in the second exon.

49. *SETX*

*SETX* is an RNA/DNA helicase involved in transcription, resolution of R-loops and DNA repair ^94^. Homozygous mutations cause ataxia with oculomoter apraxia type 2. Mice with heterozygous loss of Setx have smaller litters and early follicle loss at all stages ^95^.The variant identified in the current study has is of undetermined pathogenic significance but is found in a very conserved amino acid.

Ovarian Development and Function and Hormone Stimulation

1. *FSHR*

Homozygous and compound heterozygous mutations were found in the *FSHR*, also previously published^96,97^. Two sisters demonstrated the same frameshift mutation in *FSHR* in the final exon. Although *FSHR* is not constrained for loss of function mutations, the intracellular domain is shortened. Heterozygous *FSHR* carriers do not have early menopause. However, these subjects also carry other variants of interest.

2. *PMM2*

In addition to those previously diagnosed^49^, one subject carried two pathogenic mutations in *PMM2*^18^. A congenital disorder of glycosylation, *PMM2* mutations typically manifest with psychomotor retardation, ataxia, failure to thrive, dysmorphic features, and coagulopathy, but asymptomatic cases have been described^98^. Although the DNA was not available from the parents, the variants were close enough to demonstrate that they were on opposite alleles in IGV.

3. *TDRKH*

*TDRKH* is a conserved tudor domain protein localized on mitochondria that is critical for pre-piRNA 30 end trimming in diverse animal models^99^. *TDRKH* binds *PIWIL3* and is expressed in GV oocytes, suggesting that it works in concert with *PIWIL3* for germ cell maintenance. However, we were not able to confirm a phenotype in our *D. melanogaster model* (Table 6).

4. *USP9X*

*USP9X* was identified as a gene critical for eye and oocyte development in Drosophila (faf)^100^. The missense variant identified is found in a highly conserved amino acid.

5. *MARF1*

One subject had a homozygous missense variant in a highly conserved amino acid in *MARF1*, which was previously reported^49^. Another subject had a heterozygous variant in the same exon (exon 8), and a third subject had a heterozygous variant in a highly conserved amino acid in exon 3. *Marf1* deletions cause female infertility in mice^101^. In addition, manipulation of the C terminal of the gene by adding a GFP tag caused failure of meiosis^101^. Function of *MARF1* in the control of oocyte meiotic resumption is probably elicited by its N terminal RNase-like NYN domain through maintaining the RNA homeostasis within the oocytes (aa 352-500, exon 5-7). The RRM (RNA recognition motif) 1 that binds to ssRNA found at aa 513-581; with the current variants found at amino acids 552 and 563. Of note, the subjects with the p.Glu182Lys variant also carried a frameshift mutation in *RAD50*.

6. *MCM3AP*

Recessive *MCM3AP* mutations cause Charcot-Marie-Tooth neuropathy with intellectual disability, which has also been associated with POI^102^. *MCM3AP* encodes germinal center nuclear protein, which works as a scaffolding protein to move mRNA from the nucleus to the cytoplasm. The deleterious mutations are heterozygotes. The gene is expressed in mouse oocytes (Supplementary Table 9).

7. *MIOS*

We identified a heterozygous missense mutation in *MIOS*. The homologous gene in *Drosophila* localizes to the nucleus at the start of prophase I of meiosis and mutants fail to develop into oocytes^4^. The mutation is found before the WD40 domain number 6 at aa 395-437. *MIOS* associates with *SEH1*, which is part of the nucleoporin complex^4^.

8. *NAP1L4*

*NAP1L4* expression is high in testis, and high in the oocyte (Supplementary Table 9). The protein is known to replace histones with transition proteins during spermatogenesis in the creation of nucleosomes^37^. A role in the oocyte has not been described, but the high expression suggests it is an interesting candidate gene (Supplementary Table 9). However, we were not able to confirm a phenotype in our *D. melanogaster model* (Table 6).

9. *GJA1*

*GJA1* is a connexin that forms gap junctions. Deletion in a murine model demonstrates loss of germ cells starting in fetal life^103^. The novel missense variant identified in one subject is a possible candidate in women.

10. *HSD17B4*

One subject had a heterozygous frameshift mutation in *HSD17B4*, which is a constrained gene for loss of function variants. Although the subject also carried a missense variant (c.392G>A, p.Arg131His), the variant is found in over 40% of the population and is unlikely to be playing a role to cause POI. There is no phenotype described for heterozygotes with loss of function mutations in *HSD17B4* but the finding is compelling for POI.

11. *ZFR2*

Some of the previous candidate genes have been identified in women with POI, such as *ZFR2*. The gene was found as a heterozygous deletion on chromosome 19 in a region that contained 3 additional genes. Although a reasonable candidate based only on expression in oocytes, there is no functional data supporting the gene^104,105^

12. *AFF2*

*AFF2*, previously known as *FMR2*, demonstrated an increase in the repeat number in women with POI^106^ The missense variant was identified at a highly conserved amino acid, with no microdeletions noted.

13. *SMAD3*

*SMAD3* protein is a downstream signaling protein for TGFb. *SMAD3* deletions demonstrate decreased follicle growth, high FSH and increased apoptosis^107^. All of these features are consistent with decreased FSH signaling.

14. *MRPS22*

One subject had a heterozygous splice donor variant at a highly conserved site in the known gene mitochondrial ribosomal protein s22, *MRPS22*^108^. The gene encodes a component of the small 28S ribosomal subunit. The splice variant might affect the final two exons in the protein, which are distal to the homozygous missense variants previously identified^108^. Knockdown in a *Drosophila* model demonstrated absence of germ cells^108^. We also identified the same variant previously found in a homozygous state. The c.605G>A, p.Arg202His mutation, demonstrated no defects in OXPHOS activity or mitochondrial rRNA levels^108^. The absence of effect is consistent with the relatively milder phenotype of POI and the absence of lactic acidosis. The same subject carried an additional variant in *POLRMT*, responsible for protein translation in mitochondria and may thus affect mitochondrial protein synthesis via two mechanisms.

15. *AMH*

We identified one frameshift variant in the AMH gene that truncates the protein in exon 1 of 5, and would therefore be expected to result in nonsense mediated decay. Heterozygous *AMH* missense variants have been associated with POI in previous studies^109^, and a mouse model demonstrates that deletion of *AMH* results in follicle loss^110^. The *AMH* gene is not highly constrained^61^, however, loss of one allele and haploinsufficiency might be expected to affect AMH protein action.

16. *ZP1*

One subject demonstrated a frameshift in *ZP1*. The frameshift mutation removes the *ZP* and transmembrane domain, in a similar region found in a previous subject identified with a homozygous frameshift variant p. I390fs404Ter^111^.

17. *FRZB*

*FRZB* inhibits Wnt signaling and is upregulated in the theca interna in bovine follicles^112-114^. *FRZB* is also expressed in the epicardium and pericardium of the heart^115^, which may have implications based on Tetralogy of Fallot in the subject. Interestingly, the subject also carries a missense mutation in *LEO1*, which has been identified as essential for cardiac development at the atrioventricular border in zebrafish^116^. It is possible that both gene mutations were responsible for the cardiac defect, although the characterization is beyond the scope of the current manuscript. *LEO1* has no previously identified role in ovarian or follicle development.

18. *NANOGNB*

*NANOGNB* is found in 8 cell and morula stage embryos^117^. It regulates downstream gene expression and appears to be critical for embryo development.

19. *TSG101*

*TSG101* is a component of the endosomal sorting complex and interacts with TEX14, a component of germ cell intercellular bridges in both male and female germ cells. It participates in the final stages of cell separation after mitosis^118^. The gene is highly expressed in mouse oocytes (Supplementary Table 9).

20. *HNRNPK*

Heterogeneous nuclear ribonucleoprotein K, *HNRNPK*, is involved in transcription, RNA splicing and translation^119^. Knockdown in rat primordial follicles demonstrate that it is required for normal primordial follicle assembly and ovarian development^119^. We identified a missense variant in a conserved amino acid in the highly constrained gene. The gene is highly expressed in mouse oocytes (Supplementary Table 9).

21. *NR2F2*

*NR2F2* is a nuclear transcription factor that The missense mutation identified is novel, in a protein that is not tolerant to missense mutations. The mutation is found in the hinge region. Frameshift mutations in the DNA binding domain cause eyelid abnormalities and cardiac anomalies and XX ovotestes^120^.

22. *CPZ*

*CPZ* is carboxypeptidase Z, which is a secreted metallocarboxypeptidase with an N-terminal cysteine-rich region that targets binds Frizzled-like domains in WNT proteins^121^. Expression is very high in the ovary ^47^. Although the CPZ is not constrained for missense variants, the coverage was low in earlier version so gnomAD, and the variant of interest was confirmed in the more recent version^80^. It appears to be a part of the extracellular matrix and may be important for trophoblast invasion in the placenta^122^. Its role in the ovary has not been studied.

Translation

1. *eIF4ENIF1*

The Pro855Leu homozygous *eIF4ENIF1* variant substitutes a cyclic amino acid with an aliphatic acid with no side chain in a coil region between alpha helices and would be expected to disrupt the coil structure^123^. The p.Ser201Arg variant is located in the nuclear import signal of eIF4ENIF1. Functional studies were performed to determine the effect of *eIF4ENIF1* variants using a PA-1 cell line^124^. The c.603T>G (p.Ser201Arg) variant transfected into PA-1 cells demonstrated decreased nuclear localization (Supplementary Figure 17).

2. *eIF2B2*

A heterozygous frameshift deletion was confirmed in *eIF2B2.* The patient with the heterozygous mutation had menopause at age 39 years. She had no known neurologic disease. Of note, one previous subject with ovarioleukodystrophy also carried a frameshift mutation in *eIF2B2*, although that patient carried an additional missense variant, and had menopause at age 26 years^125^. Although previous studies have not identified *eIF2B* mutations in women with isolated POI (n=93)^126^, the current data suggests it may play a role in some POI.

3. *DCP2*

*DCP2* decaps mRNA to stop translation, which is critical for regulatory control in the oocyte and embryo. There is high expression in testes and ovary^47,127^. Therefore, it remains an interesting candidate variant. However, we were not able to confirm a phenotype in our *D. melanogaster model* (Table 6).

4. *RPL5*

*RPL5* codes for a ribosome biogenesis protein. Haploinsufficiency causes Diamond Blackfan Anemia, which is manifested by anemia, congenital malformations and growth retardation^128^. The disorder is associated with urogenitial anomalies, although POI has not been associated with the disease at this time. Expression of *RPL5* was high in eel previtelline oocytes and is highly expressed in mouse oocytes (Supplementary Table 9)^129^.

5. *BDP1*

*BDP1* is a component of TFIIIB complex that recruits RNA Pol III to begin transcription^130^. The C terminal, which is lost by the frameshift, is not critical for transcription but is important for regulation. Recessive frameshift mutations at a similar amino acid result in deafness^131^. The gene is highly expressed in mouse oocytes (Supplementary Table 9).

6. *ZNF572*

*ZNF572* is a zinc finger protein with no known function. It is highly expressed in the ovary, and the frameshift variant is found in males, only, making its frameshift mutation of interest^47^.

7. *EEF2*

*EEF2* is a eukaryotic elongation factor important for protein translation, specifically for the translocation of peptidyl-tRNA in the ribosome. It is highly expressed in the ovary and in mouse oocytes (Supplementary Table 9)^47^. A missense mutation has been identified in a family presenting with spinocerebellar ataxia^132^. The missense variant identified in the current subject is in a highly conserved amino acid.

8. *CPEB3*

Cytoplasmic polyadenylation binding protein 3, *CPEB3*, promotes polyadenylation induced translation^133^. CPEB3 appears to be expressed primarily in the brain and testis, but is also found in the ovary, and is expressed in mouse oocytes (Supplementary Table 9)^47^.

The frameshift mutation is found early in the first exon and would be expected to delete one copy of the protein. However, we were not able to confirm a phenotype in our *D. melanogaster model* (Table 6).

Autoimmune

1. *PTPN22*

We have identified a rare variant in *PTPN22* in a woman with adrenal insufficiency and POI. *PTPN22* can dephosphorylate kinases that activate T cell receptors to limit T cell signaling and particularly enhances T memory cells and raises the threshold for autoantigen activation^134^. A common *PTPN22* variant, Arg620Trp, which limits *PTPN22* ability to dephosphorylate has previously been identified as a susceptibility locus in many autoimmune diseases, including adrenal insufficiency^135^. Knock out mouse models generally demonstrate an increased propensity to autoimmune disease^134^. The variant in the current subject was identified in only one female subject and is classified as probably damaging and deleterious^80^.

2. *TARBP1*

We identified a frameshift mutation in *TARBP1*. The gene is mainly expressed in T cells and we found only minimal expression in oocytes (Supplementary Table 9). Variants in the *TARBP1* locus are associated with Sjogren’s, multiple sclerosis and lupus^136,137^. The protein plays a role in cell proliferation and transcription elongation. It is also suggested to play a role in DNA repair.
