## Supplemental Figures for "Causal and Candidate Gene Variants in a Large Cohort of Women with Primary Ovarian Insufficiency"

### Supplemental Figure 1

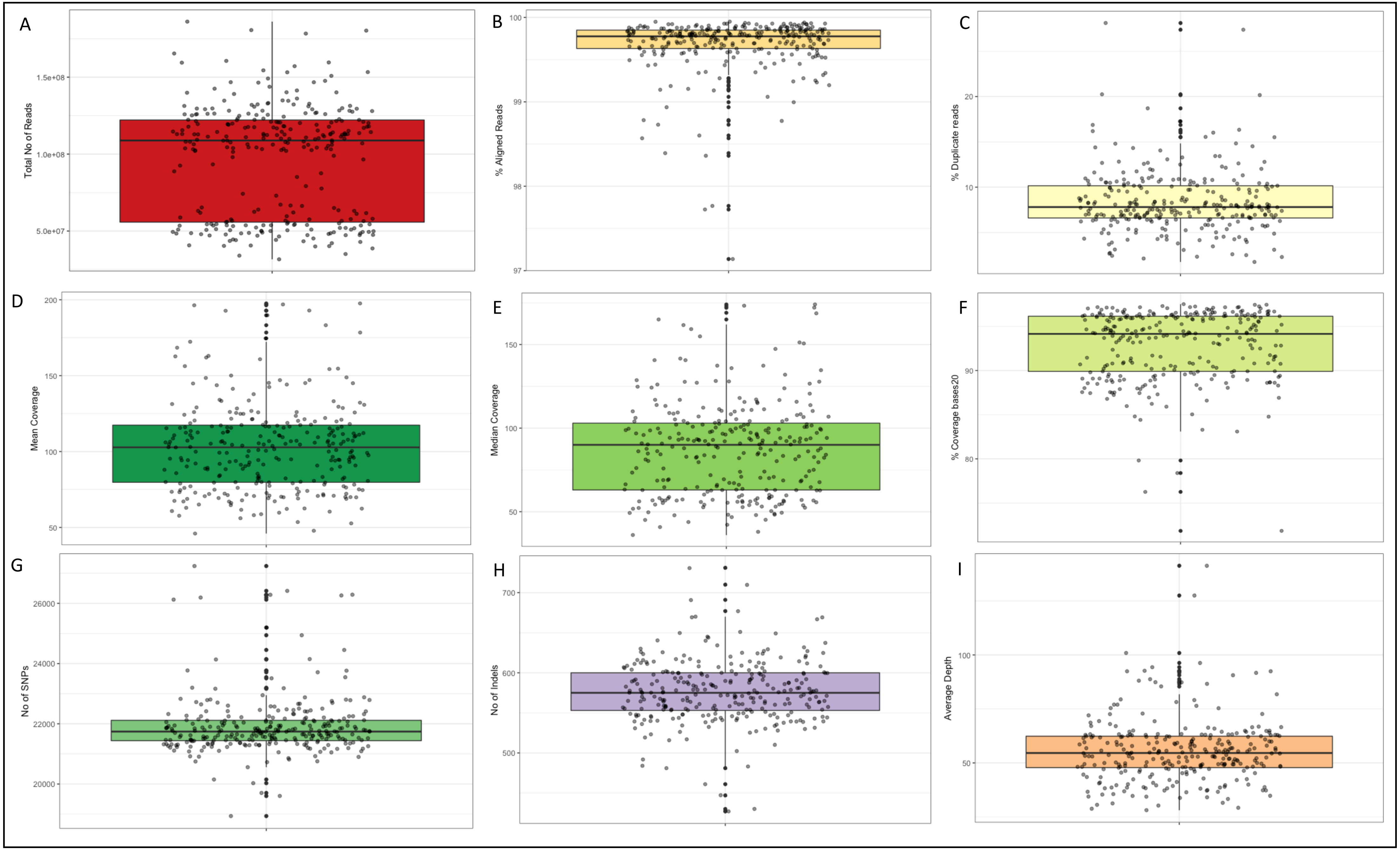

### Supplemental Figure 2

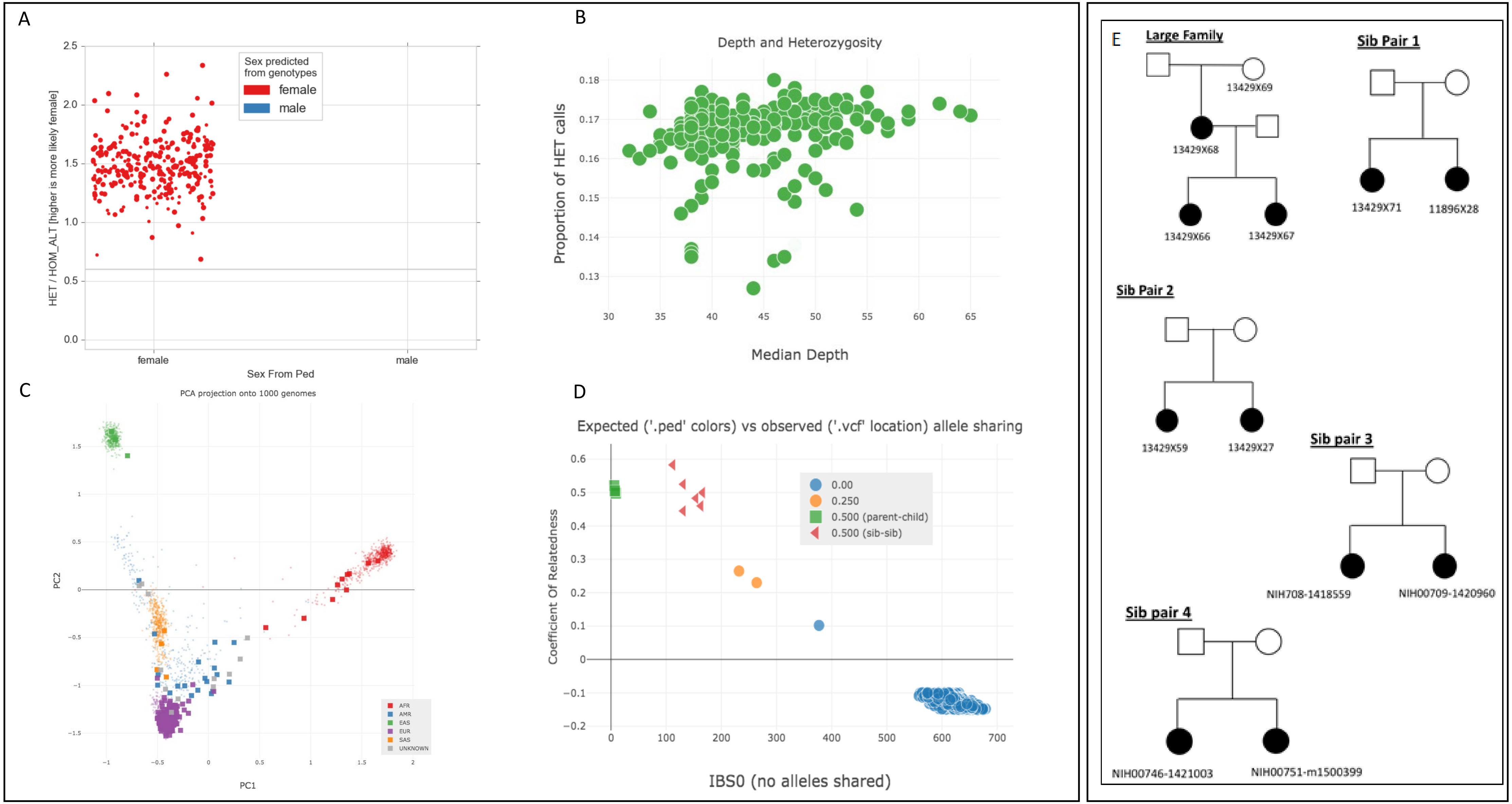

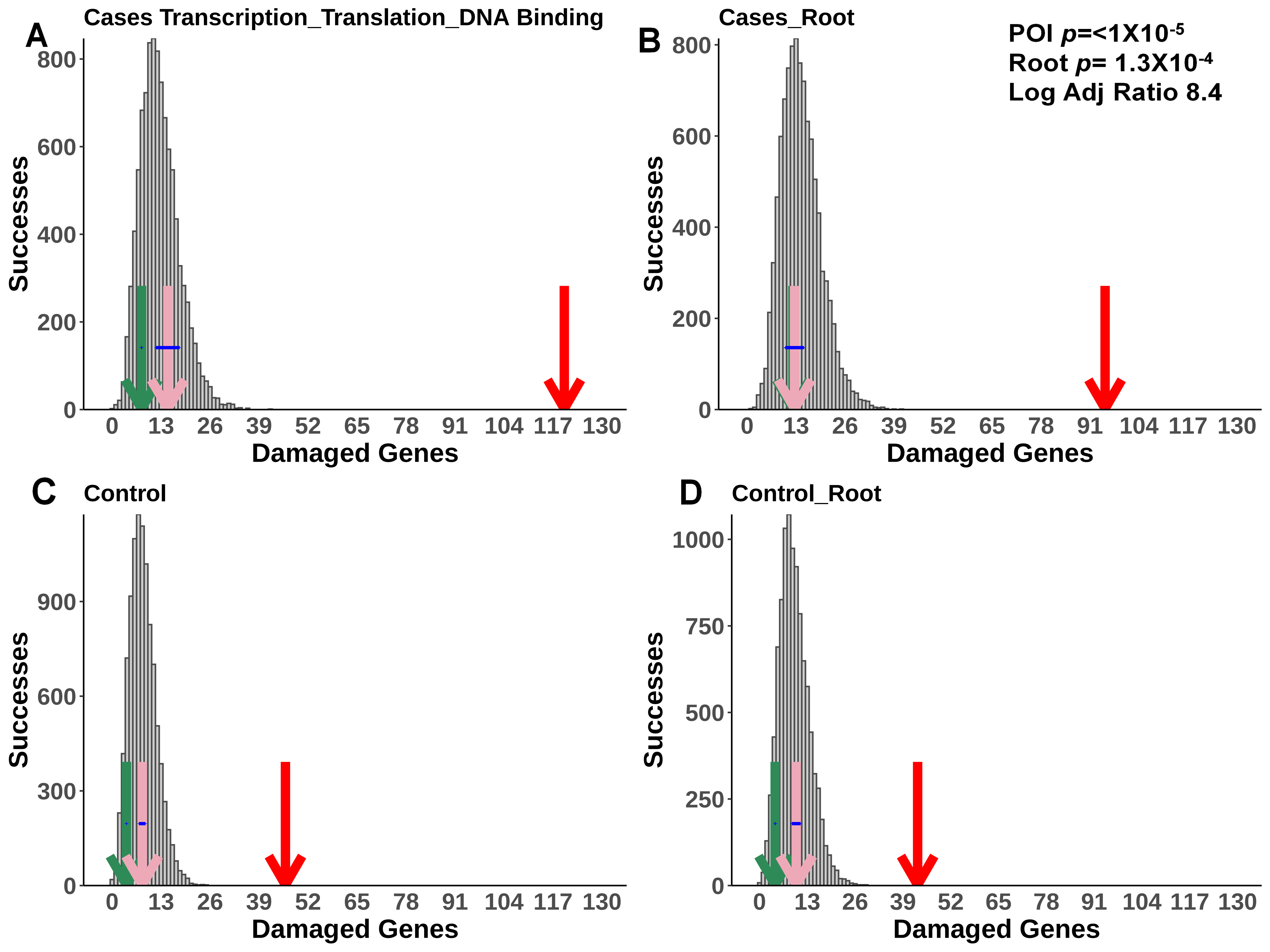

A
Cases Transcription_Translation_DNA Binding
Successes
800
600
400
200
0
0 13 26 39 52 65 78 91 104 117 130
Damaged Genes
B
Cases_Root
POI p=<1X10-5
Root p= 1.3X10-4
Log Adj Ratio 8.4
Successes
800
600
400
200
0
0 13 26 39 52 65 78 91 104 117 130
Damaged Genes
C
Control
Successes
900
600
300
0
0 13 26 39 52 65 78 91 104 117 130
Damaged Genes
D
Control_Root
Successes
1000
750
500
250
0
0 13 26 39 52 65 78 91 104 117 130
Damaged Genes

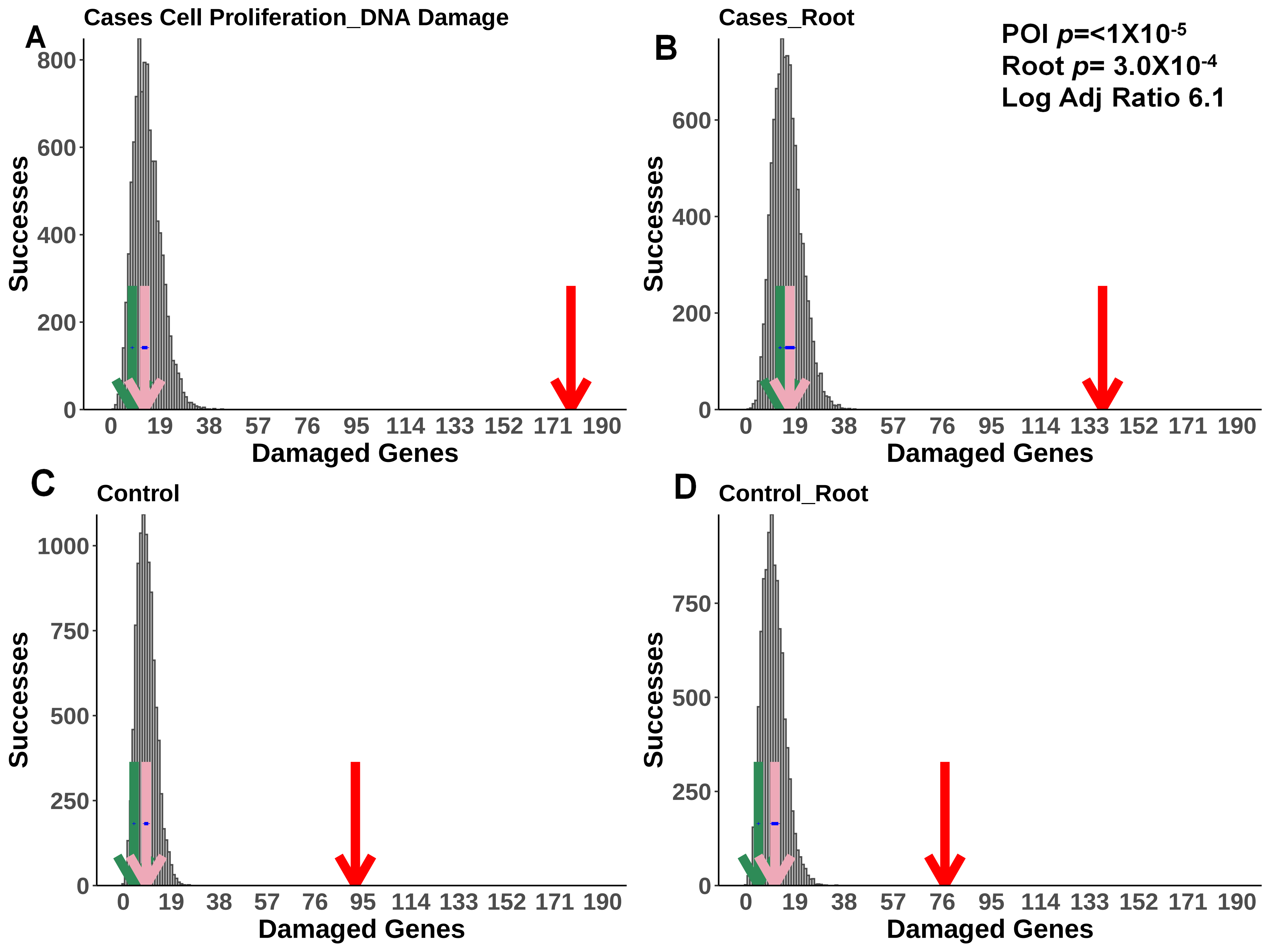
A
Cases Cell Proliferation_DNA Damage
Successes
Damaged Genes
B
Cases_Root
POI p=<1X10-5
Root p= 3.0X10-4
Log Adj Ratio 6.1
Successes
Damaged Genes
C
Control
Successes
Damaged Genes
D
Control_Root
Successes
Damaged Genes

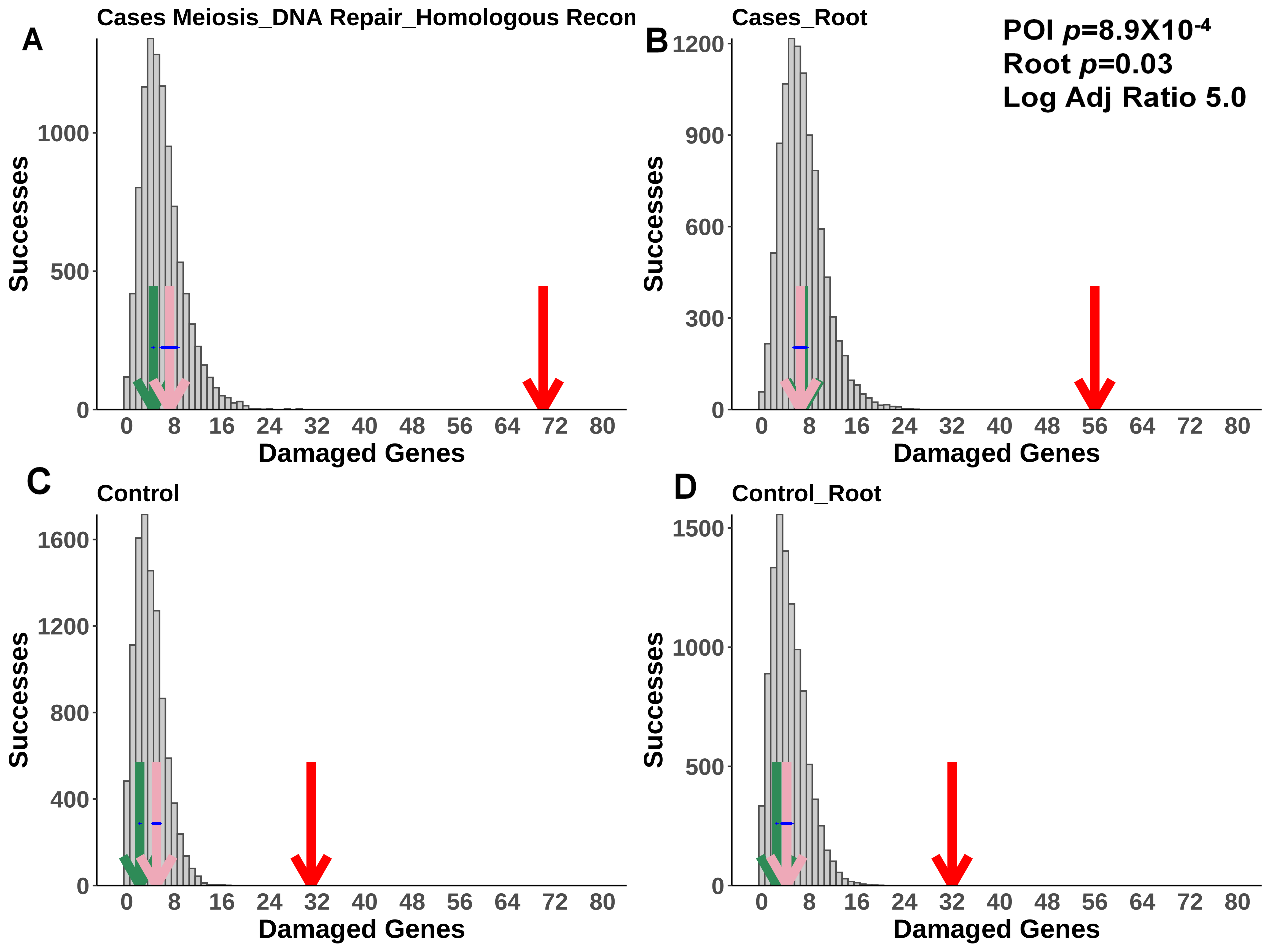
A
Cases Meiosis_DNA Repair_Homologous Recon
Successes
Damaged Genes
B
Cases_Root
POI p=8.9X10-4
Root p=0.03
Log Adj Ratio 5.0
Successes
Damaged Genes
C
Control
Successes
Damaged Genes
D
Control_Root
Successes
Damaged Genes

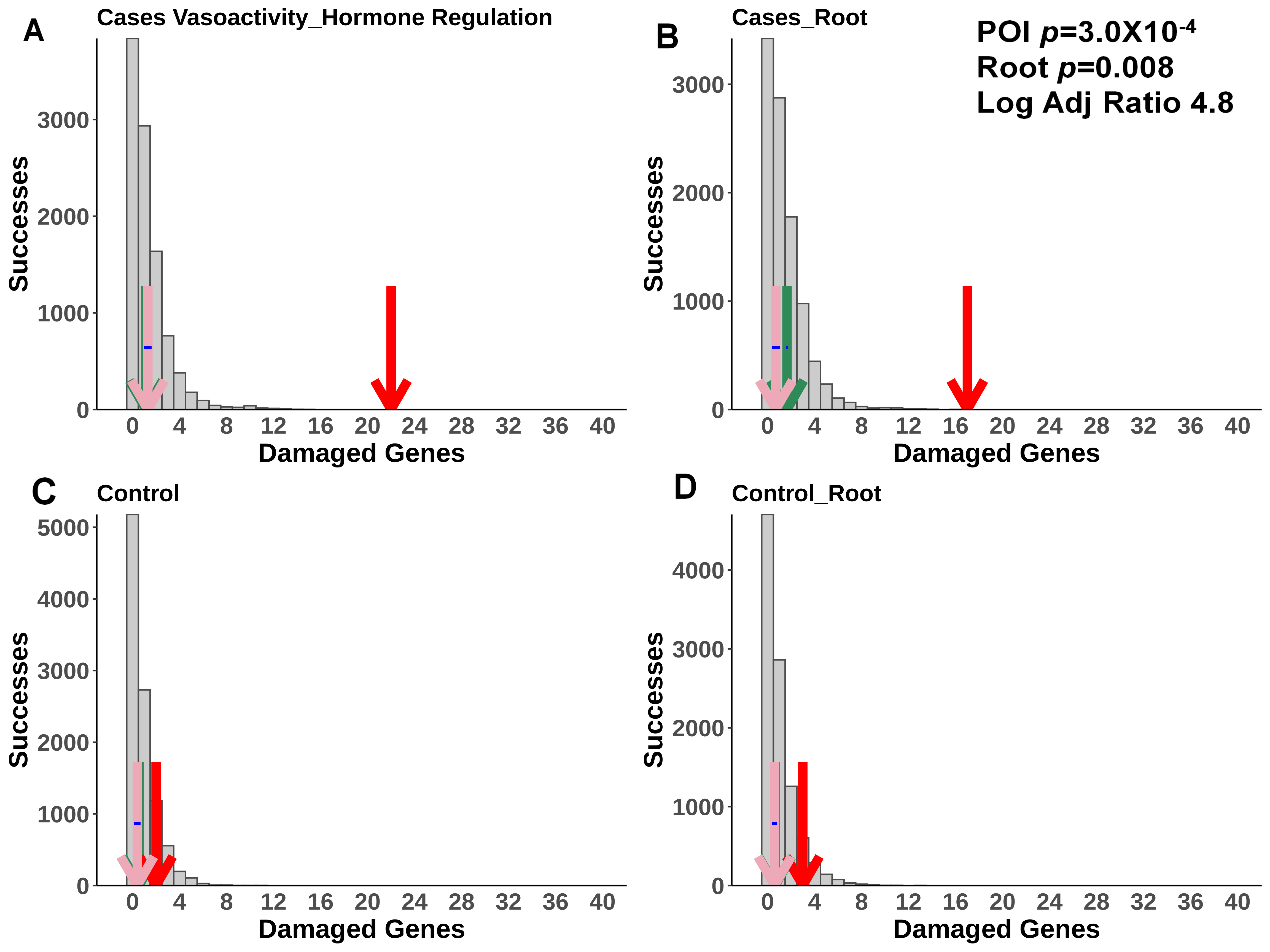
A
Cases Vasoactivity_Hormone Regulation
B
Cases_Root
POI $p$=3.0X10$^{-4}$
Root $p$=0.008
Log Adj Ratio 4.8
C
Control
D
Control_Root
Successes
Damaged Genes
0 4 8 12 16 20 24 28 32 36 40
0 1000 2000 3000
0 1000 2000 3000 4000 5000

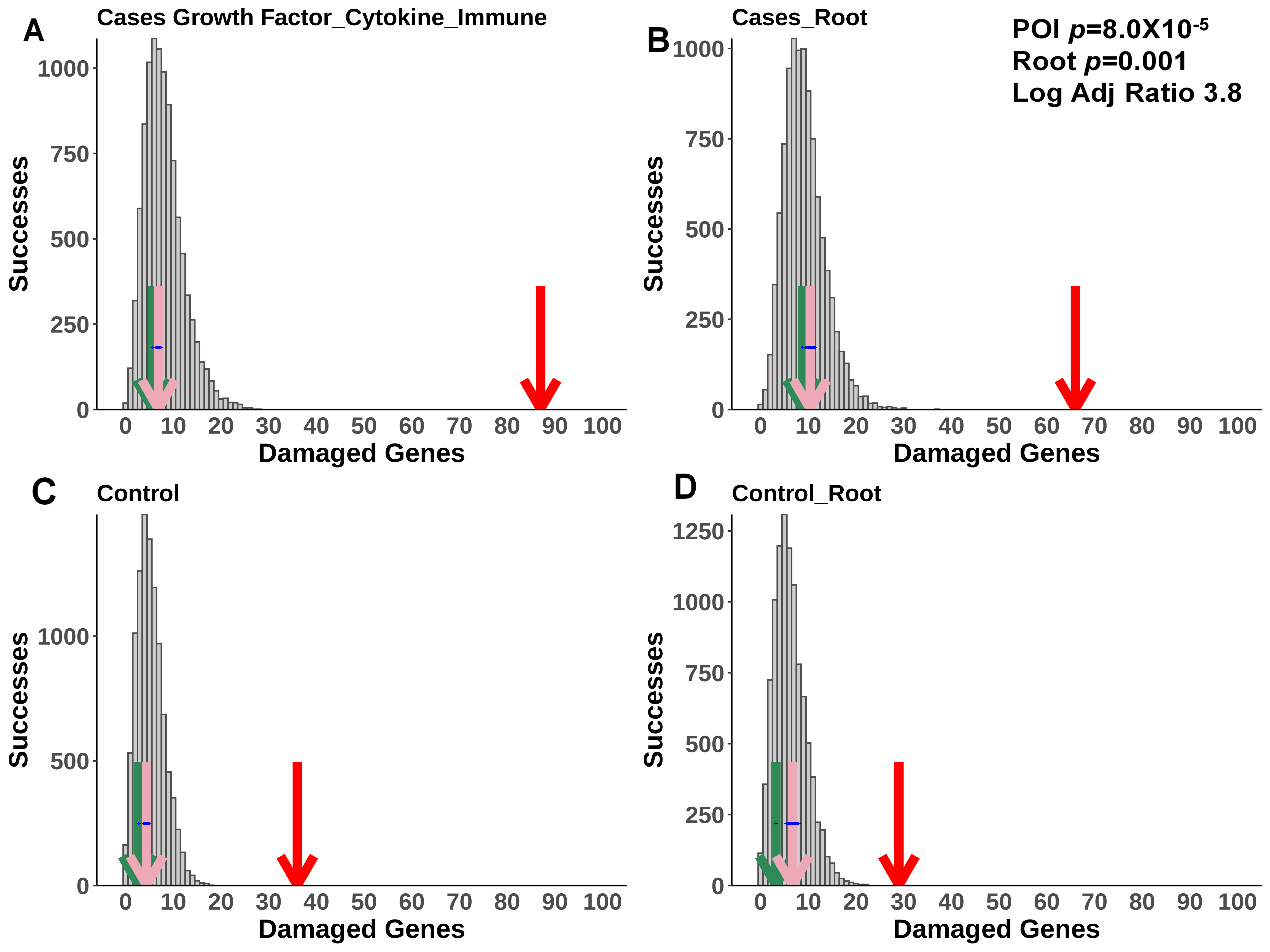
A
Cases Growth Factor_Cytokine_Immune
B
Cases_Root
POI p=8.0X10-5
Root p=0.001
Log Adj Ratio 3.8
C
Control
D
Control_Root
Successes
Damaged Genes
0
250
500
750
1000
1250
0 10 20 30 40 50 60 70 80 90 100

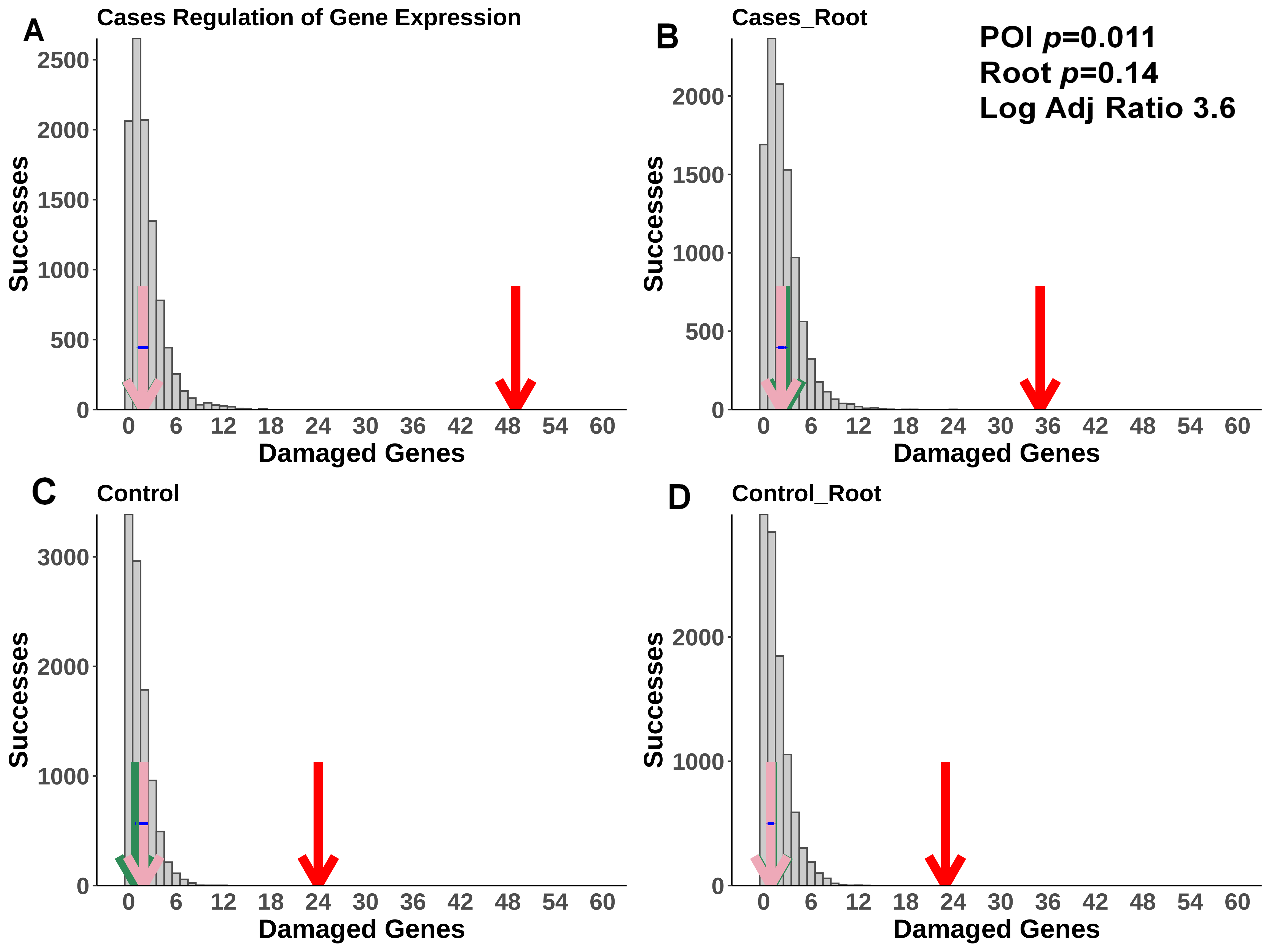
A
Cases Regulation of Gene Expression
B
Cases_Root
POI p=0.011
Root p=0.14
Log Adj Ratio 3.6
C
Control
D
Control_Root
Successes
Damaged Genes
0
6
12
18
24
30
36
42
48
54
60
0
500
1000
1500
2000
2500
3000

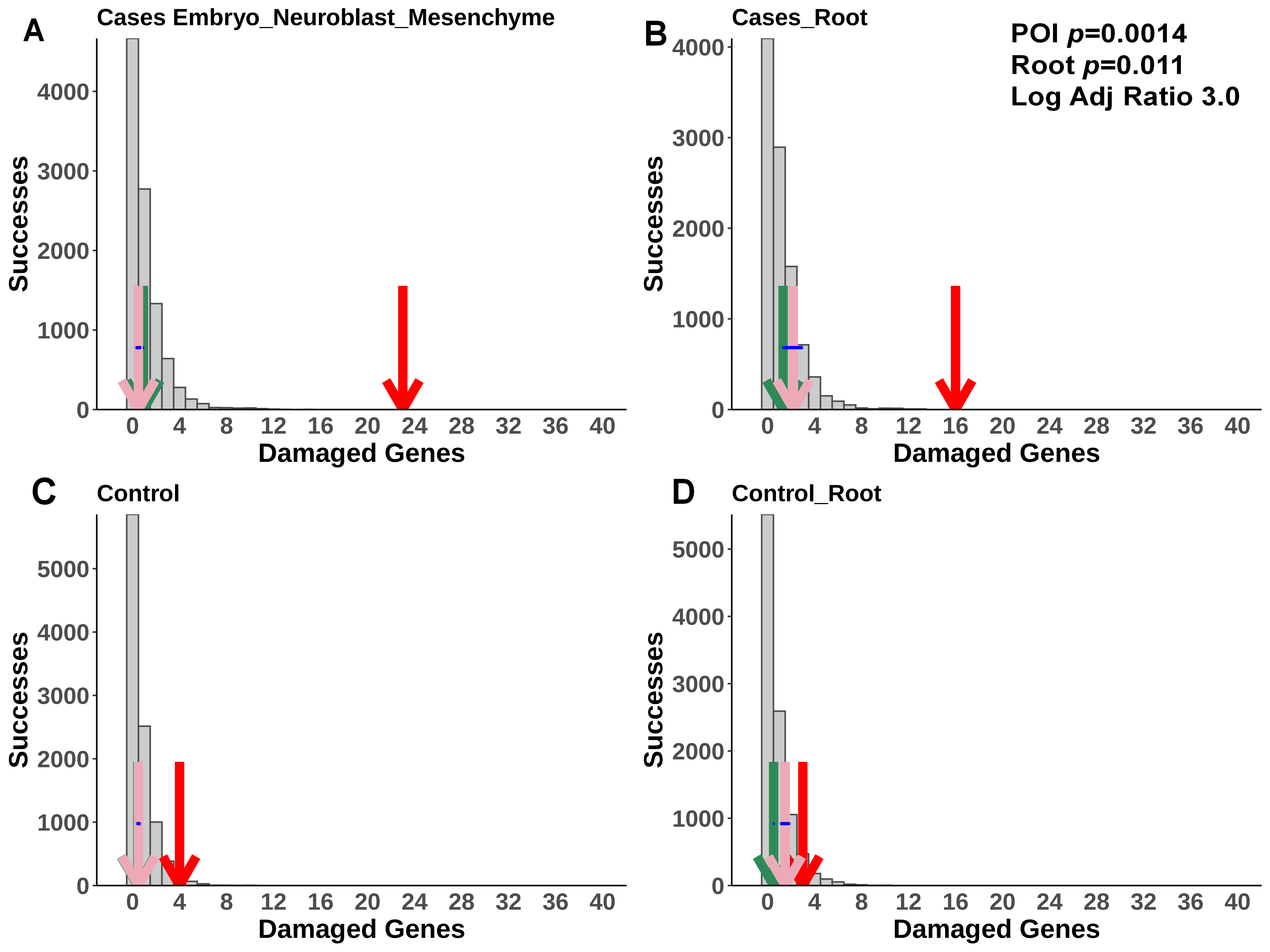

A
Cases Embryo_Neuroblast_Mesenchyme
B
Cases_Root
POI *p*=0.0014
Root *p*=0.011
Log Adj Ratio 3.0
C
Control
D
Control_Root
Successes
Damaged Genes
0
1000
2000
3000
4000
5000
0 4 8 12 16 20 24 28 32 36 40

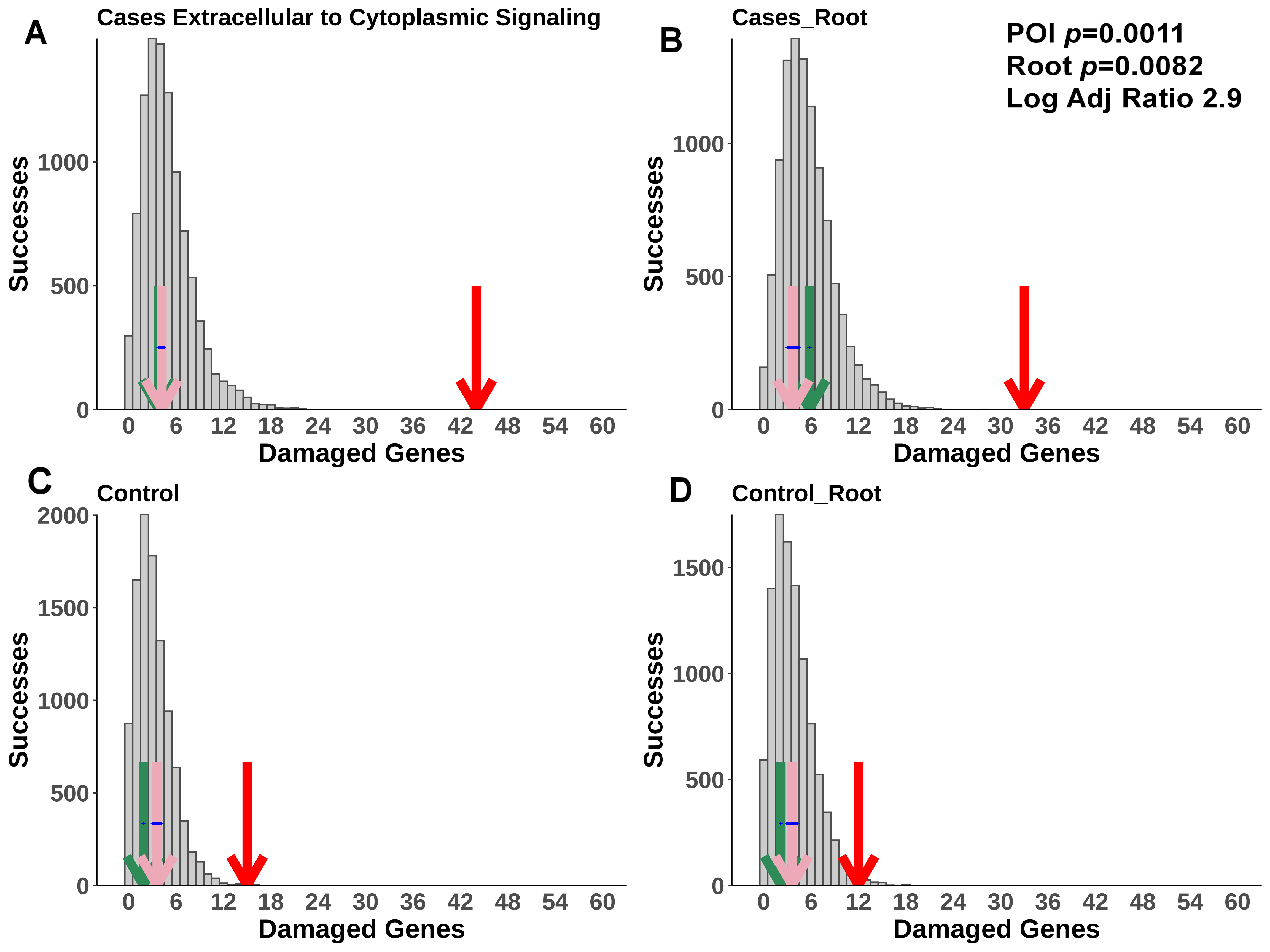
A
Cases Extracellular to Cytoplasmic Signaling
B
Cases_Root
POI p=0.0011
Root p=0.0082
Log Adj Ratio 2.9
C
Control
D
Control_Root
Successes
Damaged Genes
0
6
12
18
24
30
36
42
48
54
60
500
1000
1500
2000

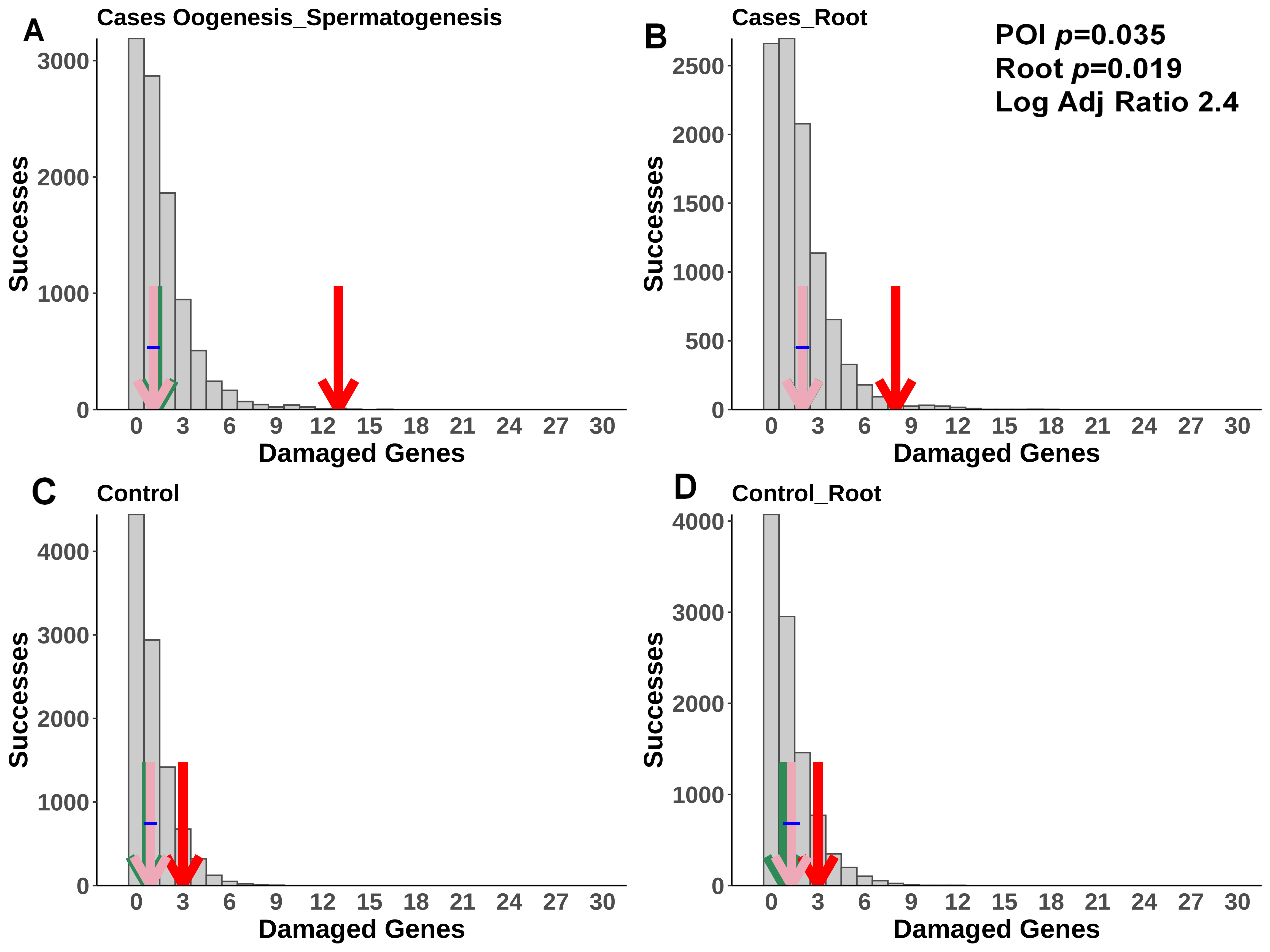
A
Cases Oogenesis_Spermatogenesis
Successes
Damaged Genes
B
Cases_Root
POI p=0.035
Root p=0.019
Log Adj Ratio 2.4
Successes
Damaged Genes
C
Control
Successes
Damaged Genes
D
Control_Root
Successes
Damaged Genes

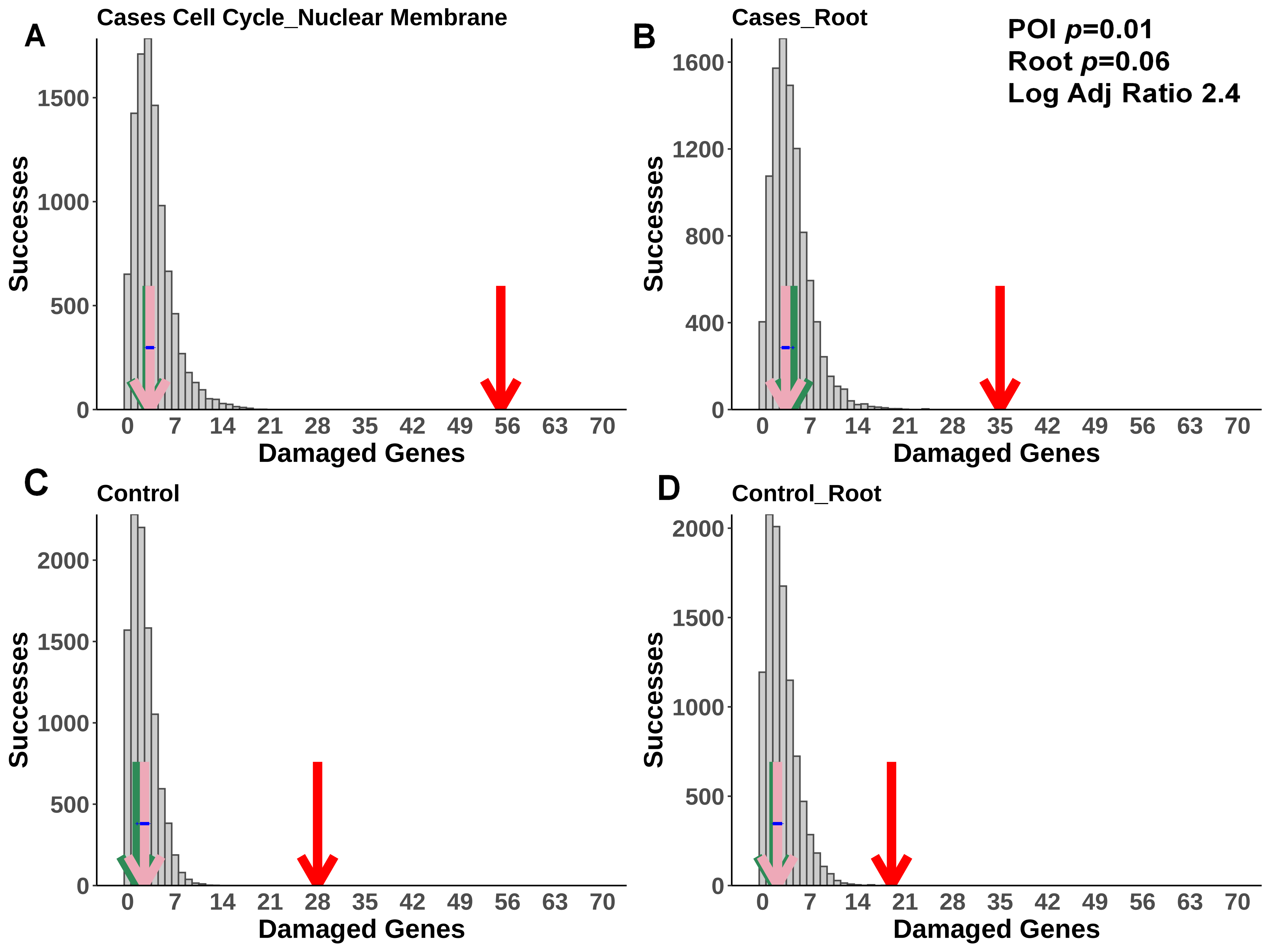
A
Cases Cell Cycle_Nuclear Membrane
Successes
Damaged Genes
B
Cases_Root
POI p=0.01
Root p=0.06
Log Adj Ratio 2.4
Successes
Damaged Genes
C
Control
Successes
Damaged Genes
D
Control_Root
Successes
Damaged Genes

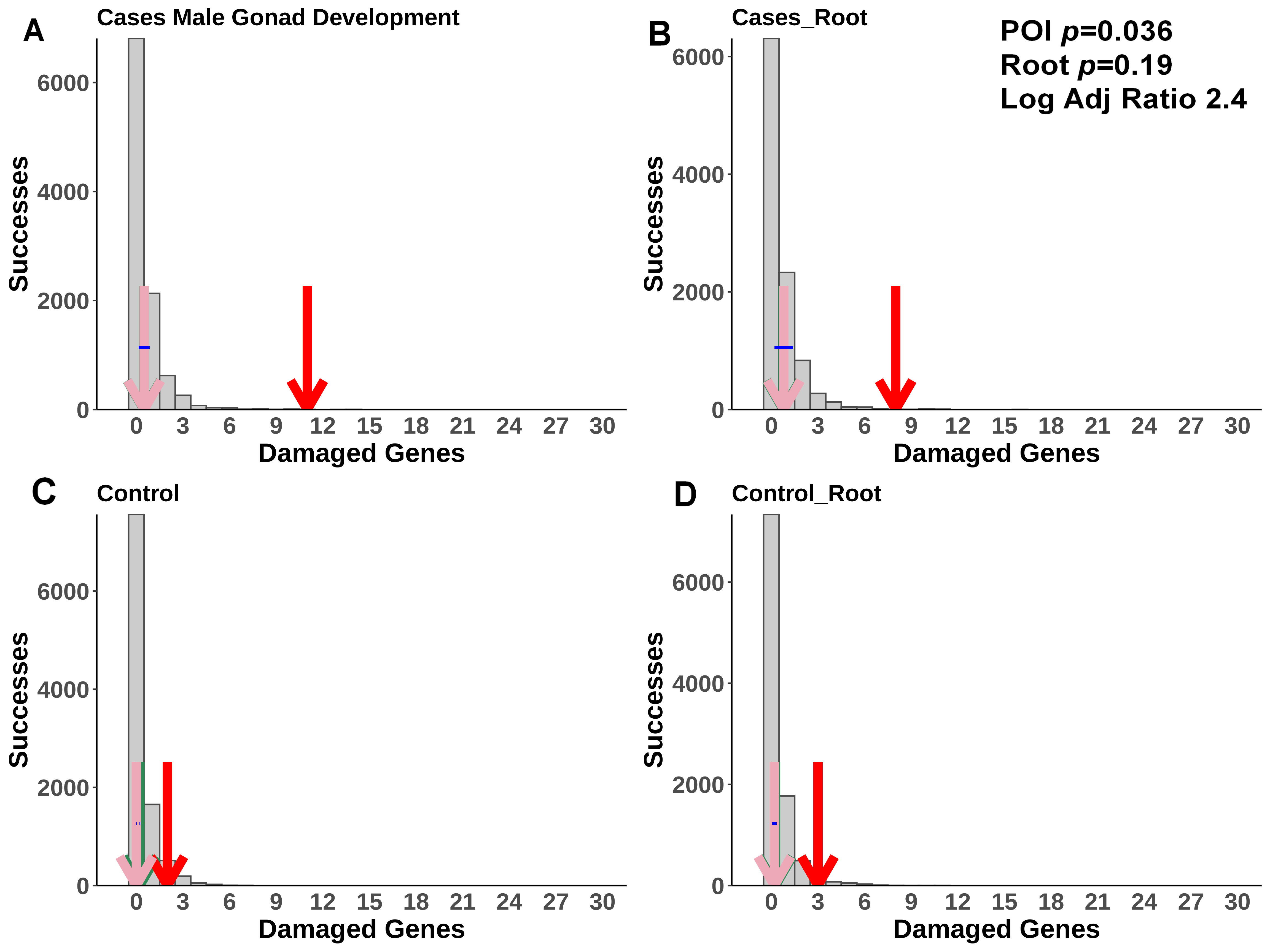

A
Cases Male Gonad Development
Successes
Damaged Genes
B
Cases_Root
POI p=0.036
Root p=0.19
Log Adj Ratio 2.4
Successes
Damaged Genes
C
Control
Successes
Damaged Genes
D
Control_Root
Successes
Damaged Genes

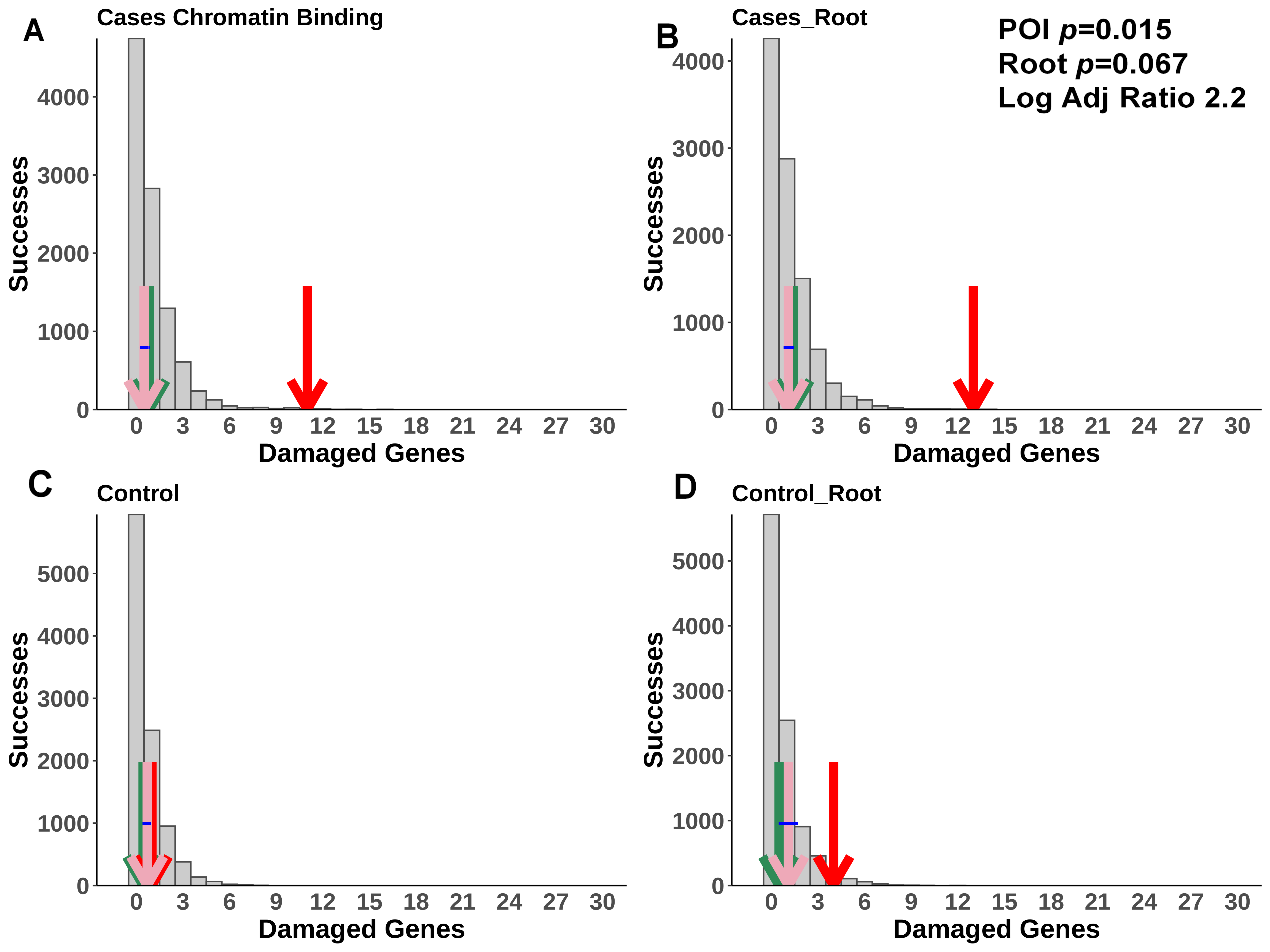
A
Cases Chromatin Binding
B
Cases_Root
POI p=0.015
Root p=0.067
Log Adj Ratio 2.2
C
Control
D
Control_Root
Successes
Damaged Genes
0
1000
2000
3000
4000
5000
0 3 6 9 12 15 18 21 24 27 30

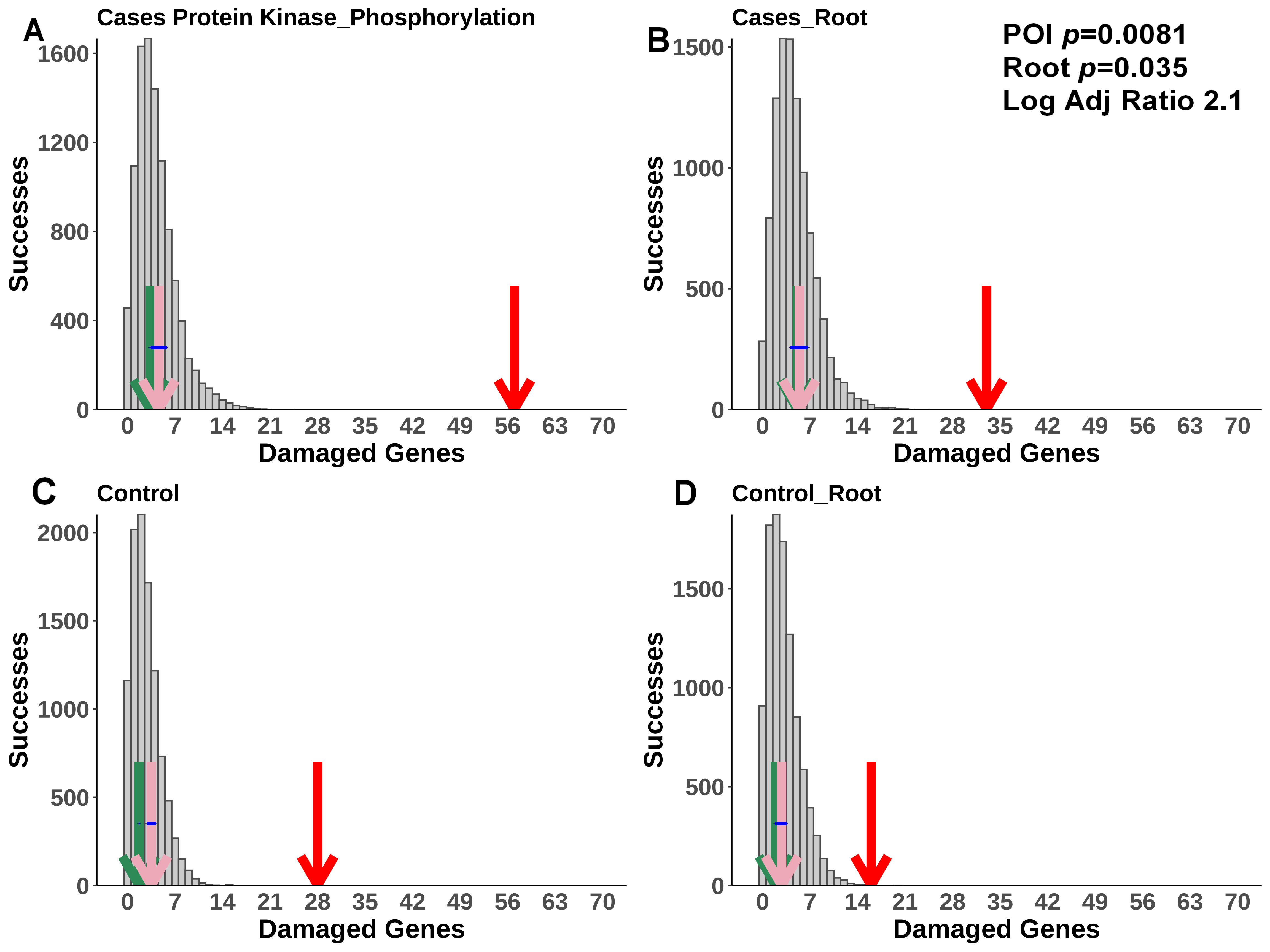

A
Cases Protein Kinase_Phosphorylation
Successes
0
400
800
1200
1600
0 7 14 21 28 35 42 49 56 63 70
Damaged Genes
B
Cases_Root
POI *p*=0.0081
Root *p*=0.035
Log Adj Ratio 2.1
Successes
0
500
1000
1500
0 7 14 21 28 35 42 49 56 63 70
Damaged Genes
C
Control
Successes
0
500
1000
1500
2000
0 7 14 21 28 35 42 49 56 63 70
Damaged Genes
D
Control_Root
Successes
0
500
1000
1500
0 7 14 21 28 35 42 49 56 63 70
Damaged Genes

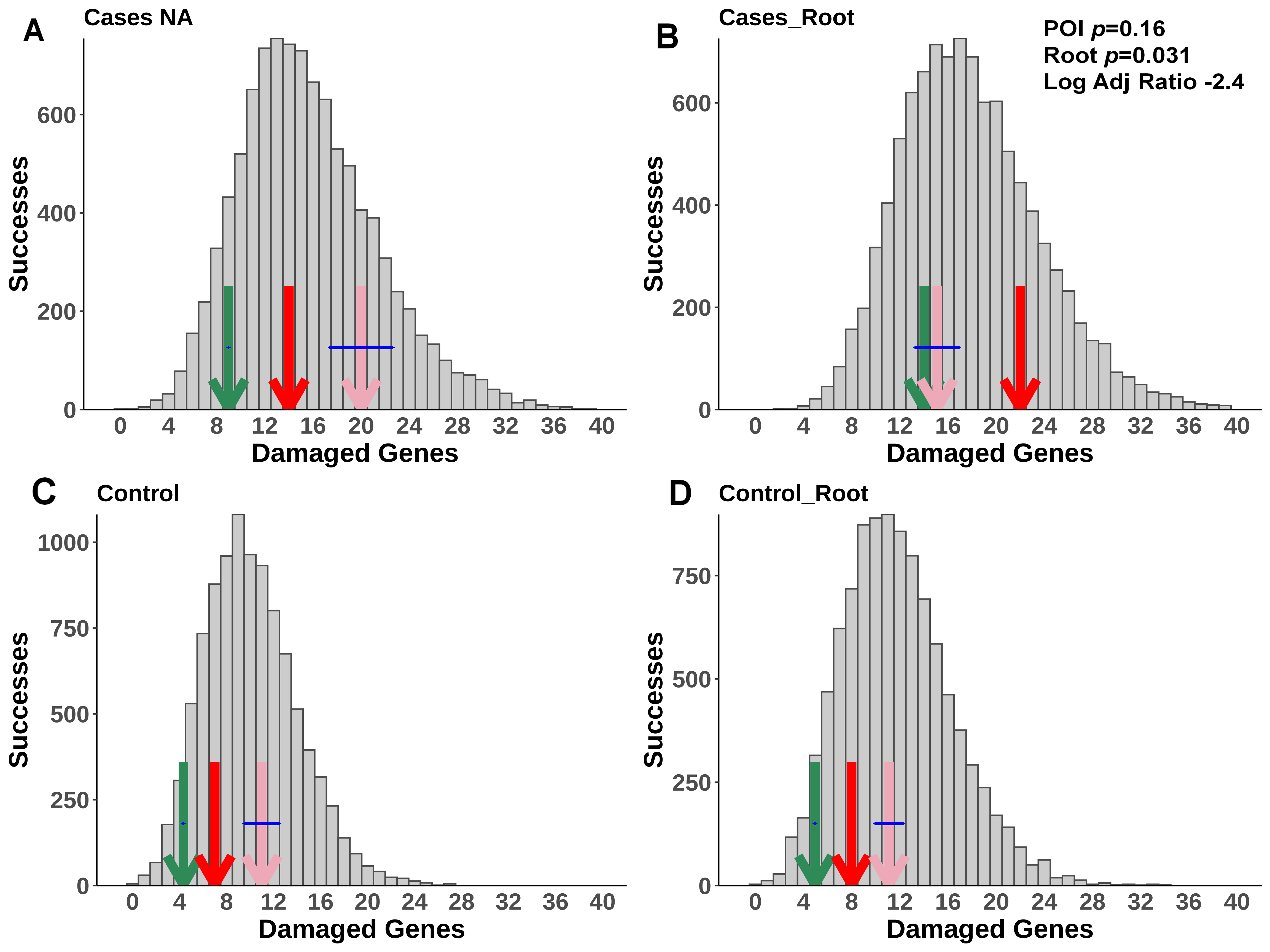

A
Cases NA
Successes
0
200
400
600
0 4 8 12 16 20 24 28 32 36 40
Damaged Genes
B
Cases_Root
POI *p*=0.16
Root *p*=0.031
Log Adj Ratio -2.4
Successes
0
200
400
600
0 4 8 12 16 20 24 28 32 36 40
Damaged Genes
C
Control
Successes
0
250
500
750
1000
0 4 8 12 16 20 24 28 32 36 40
Damaged Genes
D
Control_Root
Successes
0
250
500
750
0 4 8 12 16 20 24 28 32 36 40
Damaged Genes

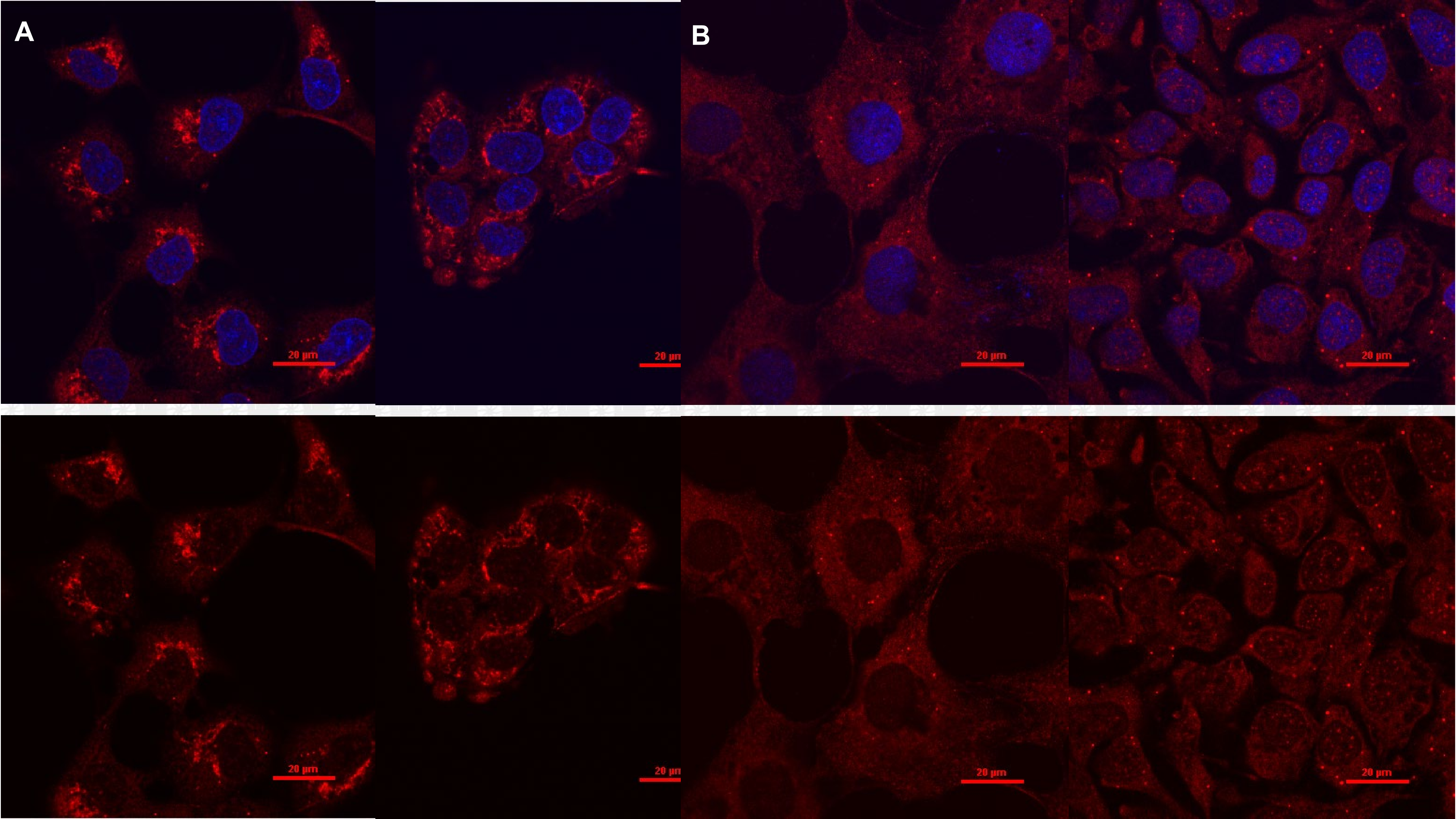
A
B
20 μm
20 μm
20 μm
20 μm
20 μm
20 μm
20 μm
20 μm

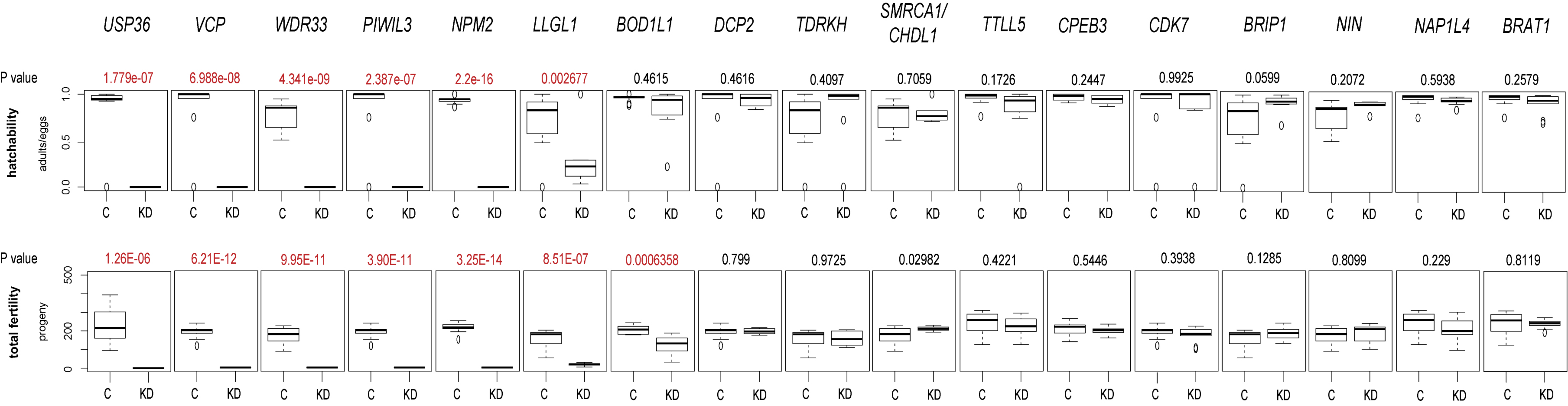

USP36
VCP
WDR33
PIWIL3
NPM2
LLGL1
BOD1L1
DCP2
TDRKH
SMRCA1/ CHDL1
TTLL5
CPEB3
CDK7
BRIP1
NIN
NAP1L4
BRAT1
P value
1.779e-07
6.988e-08
4.341e-09
2.387e-07
2.2e-16
0.002677
0.4615
0.4616
0.4097
0.7059
0.1726
0.2447
0.9925
0.0599
0.2072
0.5938
0.2579
hatchability
adults/eggs
1.0
0.5
0.0
C
KD
P value
1.26E-06
6.21E-12
9.95E-11
3.90E-11
3.25E-14
8.51E-07
0.0006358
0.799
0.9725
0.02982
0.4221
0.5446
0.3938
0.1285
0.8099
0.229
0.8119
total fertility
progeny
500
200
0
C
KD

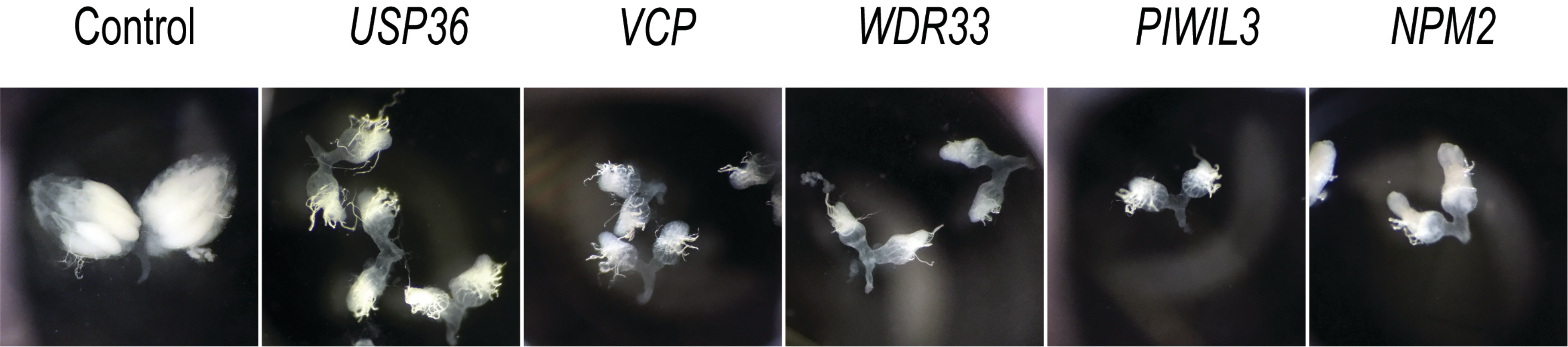
Control
*USP36*
*VCP*
*WDR33*
*PIWIL3*
*NPM2*
